# Prevalence of maternal substance use during pregnancy and first two years of life: A whole-population cohort of 970,470 Australian children born 2008-2017

**DOI:** 10.1101/2024.12.09.24318518

**Authors:** Madeline Powell, Rhiannon Pilkington, Tasnia Ahmed, Mark Hanly, BJ Newton, John W. Lynch, Timothy Dobbins, Jess Stewart, Michelle Cretikos, Alys Havard, Kathleen Falster

**Affiliations:** National Drug and Alcohol Research Centre (NDARC), University of New South Wales, Sydney, Australia; School of Population Health, Faculty of Medicine and Health, University of New South Wales, Sydney, Australia; School of Public Health, University of Adelaide, Adelaide, South Australia, Australia; Centre for Big Data Research in Health, Faculty of Medicine and Health, University of New South Wales, Sydney, Australia; Social Policy Research Centre, University of New South Wales, Sydney, Australia; Bristol Medical School, Population Health Sciences, University of Bristol, Bristol, United Kingdom; Family and Community Services Insights, Analysis and Research (FACSIAR), NSW Department of Communities and Justice, Sydney, Australia; Centre for Epidemiology and Evidence, NSW Ministry of Health, Sydney, Australia

**Keywords:** Epidemiology, maternal substance use, prevalence, child health, prevention, data linkage, child protection

## Abstract

**Objectives:** To estimate the prevalence of maternal substance use during the first 1000 days of children’s lives, to inform planning and resourcing of antenatal screening and substance use in pregnancy services, alongside antenatal and postnatal health, parenting and social support services for pregnant women/new mothers and their babies.

**Method:** This whole-population cohort was assembled from birth registration, perinatal, and hospital data for children born 2008-2017, and their mothers, using data linked for the New South Wales (NSW) Child E-Cohort Project. The primary outcome was maternal substance use and treatment recorded in six health, death, and child protection data sources from the child’s conception to age 2-years (the first 1000 days), including illicit substances, alcohol, opioid-agonist treatment, and misuse of psychoactive medicines or substances.

**Results:** Of 970,470 children born to 625,856 mothers, 3.4% (N=32,647) had ≥1 maternal substance use records in the first 1000 days, including alcohol use (N=13,647; 1.4%) and other drug use (N=23,485; 2.4%). Maternal substance use was recorded during the pregnancy period for 1.2% of children, and from 28-1000 days post-birth for 2.4% of children. Outcome ascertainment was highest from child protection records (N=26,045), followed by mother’s (N=12,956) then children’s hospital records (N=3,826). Child protection records more than doubled the prevalence from health and death records alone (1.4%). Social and health disadvantage was more common among children with maternal substance use.

**Conclusion:** During the first 1000 days of life, 3.4% of NSW children had ≥1 maternal substance use record in health, child protection and death data sources. Child protection data enhances public health intelligence on the burden of maternal substance use among whole-populations of children. Near universal health system contact during pregnancy and birth is an opportunity to initiate early support for maternal substance use and co-occurring health and social disadvantage, to promote child health and development.

**What is already known on this topic:**

- Population-level evidence of maternal substance use during the first 1000 days is limited by heterogenous study designs.
- Cross-sectional surveys and self-reports illustrate low-level use from 10-18%; studies linking one-four whole-population health datasets provide estimates of 0.2-3% for more harmful use.

**What this study adds:**

- Of 970,000 children born 2008-2017 in NSW, Australia, 3.4% had a maternal substance use record during their first 1000 days of life in six linked administrative datasets.
- Adding child protection to health and death data more than doubled prevalence estimates.

**How this study might affect research, practice or policy:**

- Public health intelligence to inform screening and support services for pregnant women, new mothers and children affected by maternal substance use can be enhanced using child protection data, in addition to health and death data sources.
- Near universal health system contact during pregnancy and birth is an opportunity to initiate early support for maternal substance use and co-occurring health and social disadvantage, to promote child health and development.

## INTRODUCTION

Maternal substance use may harm child health and development, both through exposure during gestation^1^ and impacts on parenting, family functioning, and the home environment during early life.^2-4^ Screening and support for pregnant women and new mothers using substances is important for the health of mothers, and to promote children’s health and development from the earliest opportunity. To plan prevention responses for mothers and children, contemporary data on the scale and type of maternal substance use at the population-level are needed.

Until recently, general population cross-sectional surveys have been the main source of data on maternal substance use prevalence during pregnancy. For example, alcohol use among pregnant women was reported to be 10% in the USA in 2021,^5^ 15% in Australia in 2019,^6^ and 18% in Canada in 2019.^7^ Maternal substance use recorded in administrative data also offers public health intelligence to inform prevention. To date, most studies using administrative data have reported the prevalence of single substance use during pregnancy/birth or neonatal abstinence syndrome (NAS). Some studies have used one data source, often the mother’s or child’s hospital records, to ascertain maternal substance use indicators with estimates such as 1.1% for prenatal illicit substance use;^8^ and 0.1-1% for NAS/in utero substance exposure.^9-15^ Higher prevalence estimates have been reported in studies using two to four data sources to ascertain maternal substance use from health service contacts, including hospital and mental health outpatient data (substance use, 3.2%),^16^ or prenatal screening and primary care data (alcohol use, 3.0-11.7%; illicit drug use, 3.6-6.0%).^17, 18^

In this study, we aimed to quantify the prevalence and type of maternal substance use from conception to the child’s second birthday (first 1000 days of life) to inform screening and support services that may reduce maternal substance use associated harm or risk for children. We ascertained indicators of maternal substance use and treatment from mother and child records in six health, death and child protection administrative data sources for all children born in New South Wales (NSW), the most populous state in Australia, from 2008-2017.

## METHODS

This birth cohort study used population-level administrative data linked for the NSW Child E-Cohort Project. Reporting follows the RECORD Guidelines (Supplement 1).^19^

### Data sources and data linkage

Data sources included: perinatal data (includes live births ≥20 weeks gestation or >400 grams birth weight); births and deaths registrations; hospital inpatient; opioid treatment register; cause of death; emergency department presentations; publicly funded mental health outpatients, public housing, and child protection data (Supplement 2-3). Data were linked by the NSW Centre for Health Record Linkage (false positive linkages, 0.4-0.5%).^20^

### Study population

The study population included live and still births recorded in the perinatal, birth registration and/or hospital data in NSW from July 2001 to December 2019 (N=1,895,850) (Supplement 4). Mother-child dyads were linked using the perinatal and birth registration data. We restricted the study population to children born January 2008, to December2017 (N= 990,631), so the availability of all data sources aligned with the gestation to 2^nd^ birthday period for all children (Supplement 3, 6).

### Outcomes

The primary outcome was maternal substance use, including diagnoses and treatment for maternal substance use conditions (henceforth *maternal substance use*), during the first 1000 days recorded in ≥1 of the six data sources with substance use information. *Substance use* was defined as the use of, or conditions related to the use of illicit substances or alcohol, misuse of prescription medicine, use of opioid-agonist treatment, and misuse of organic compounds, solvents or substances that produce psychoactive effects. *Substance use* was categorised into: *alcohol use* and *other drug use,* and further broken down by drug type where possible. In child and mother hospital, emergency department, mental health outpatients, and cause of death records maternal drug and/or alcohol use diagnosis codes were recorded using the International Classification of Diseases and Related Health Problems, Tenth Revision, Australian Modification (ICD-10-AM) or the Systemized Nomenclature of Medicine Clinical Terminology (SNOMED) system (Supplement 5-5a). We selected ICD-10AM and SNOMED codes from published literature^8, 21-29^ government reports,^30, 31^ and a syndicated terminology server (Ontoserver) for SNOMED (Supplement 5-5a).^32^ We ascertained maternal opioid agonist treatment (OAT) records from the opioid treatment registry.

Measures from child protection data comprised child concern reports and field assessments of suspected or actual risk of harm due to drug and/or alcohol use, that were reported by health professionals, police, educators and others in the community. This data source does not differentiate between maternal and other carers, however, there is evidence the majority of reports relate to mothers.^33, 34^ Henceforth, we include this in the maternal substance use estimates indicators from all six data sources, whilst also stratifying results by child protection records, and maternal and child health and death data sources.

#### Outcome ascertainment periods

We ascertained outcomes during the first 1000 days of children’s lives, defined as the conception date (child’s birth date minus gestational age, plus 14 days)^35^ until the child’s second birthday. We also ascertained outcomes for: 1) the *prenatal period,* commencing from the conception date until <28 days of age, as diagnoses at birth (e.g. neonatal abstinence syndrome) indicate substance use during pregnancy; and 2) the *early-life period,* commencing 28 days of age until the child’s second birthday (Supplement 6).

### Sociodemographic and health characteristics of mothers and children at birth

We describe the sociodemographic and health characteristics of children and mothers (at birth) who did and did not have substance use recorded in any data source, any health and death data source, or child protections records alone (Table 1, Supplement 7).

**Table 1.**
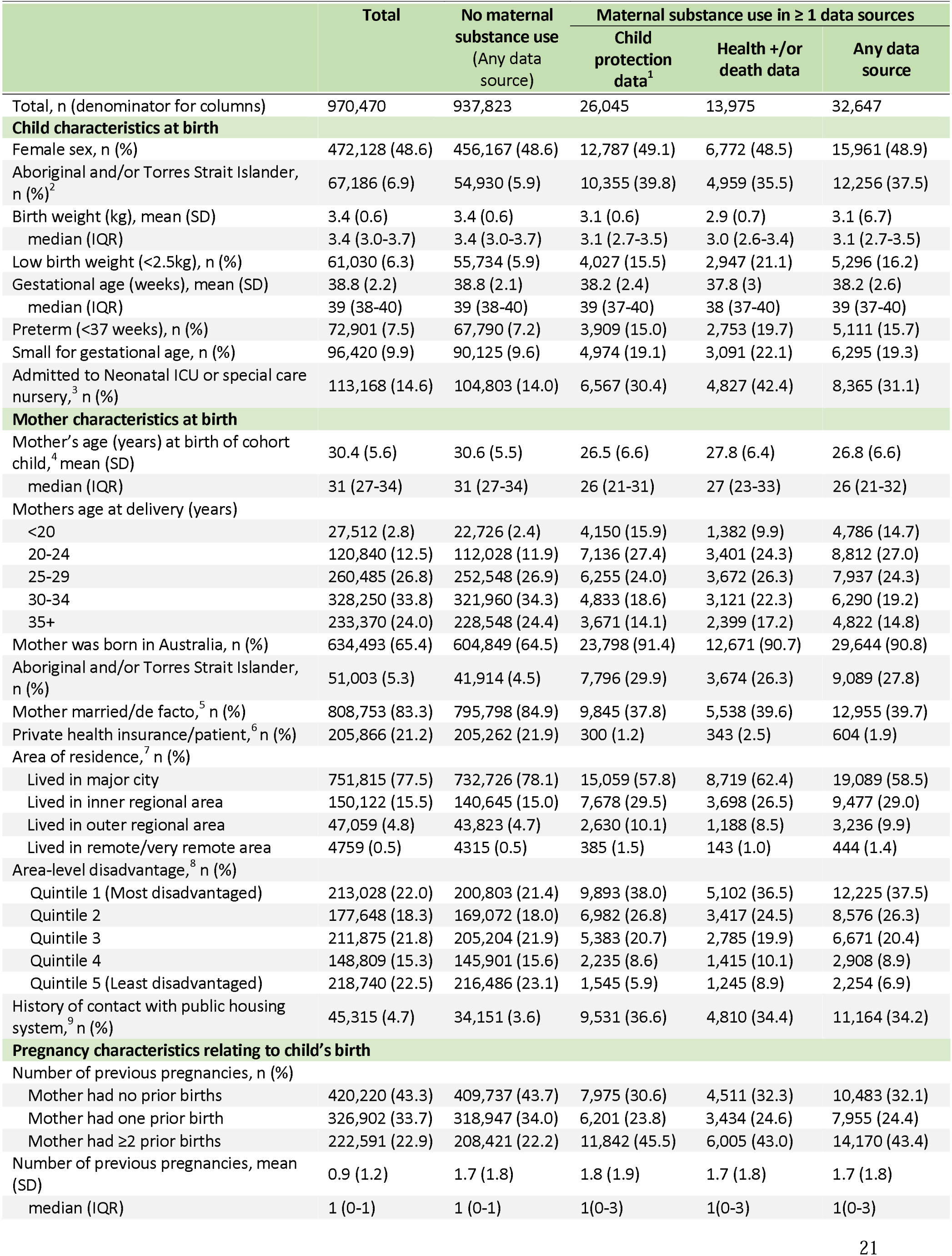
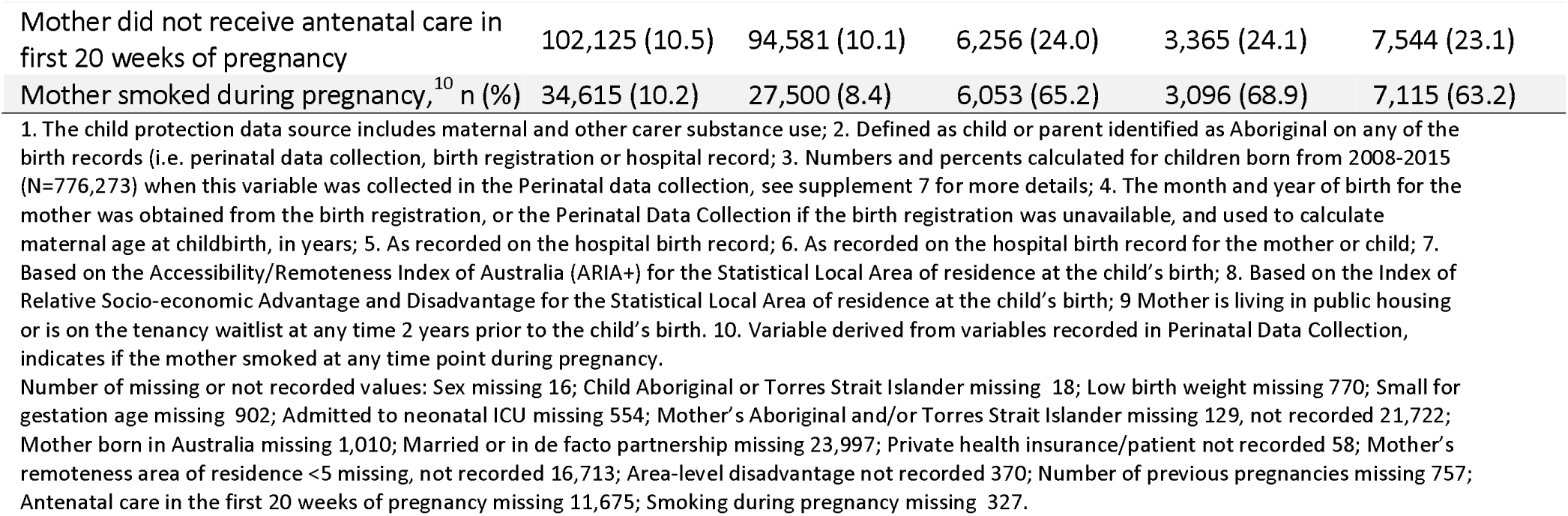
Maternal, pregnancy and child characteristics at birth of children with and without substance use recorded during the first 1000 days.

|  | Total | No maternal substance use<br>(Any data source) | Maternal substance use in ≥ 1 data sources |  |  |
| --- | --- | --- | --- | --- | --- |
|  |  |  | Child protection data<sup>1</sup> | Health +/or death data | Any data source |
| Total, n (denominator for columns) | 970,470 | 937,823 | 26,045 | 13,975 | 32,647 |
| **Child characteristics at birth** |  |  |  |  |  |
| Female sex, n (%) | 472,128 (48.6) | 456,167 (48.6) | 12,787 (49.1) | 6,772 (48.5) | 15,961 (48.9) |
| Aboriginal and/or Torres Strait Islander, n (%)<sup>2</sup> | 67,186 (6.9) | 54,930 (5.9) | 10,355 (39.8) | 4,959 (35.5) | 12,256 (37.5) |
| Birth weight (kg), mean (SD) | 3.4 (0.6) | 3.4 (0.6) | 3.1 (0.6) | 2.9 (0.7) | 3.1 (0.7) |
| median (IQR) | 3.4 (3.0-3.7) | 3.4 (3.0-3.7) | 3.1 (2.7-3.5) | 3.0 (2.6-3.4) | 3.1 (2.7-3.5) |
| Low birth weight (<2.5kg), n (%) | 61,030 (6.3) | 55,734 (5.9) | 4,027 (15.5) | 2,947 (21.1) | 5,296 (16.2) |
| Gestational age (weeks), mean (SD) | 38.8 (2.2) | 38.8 (2.1) | 38.2 (2.4) | 37.8 (3) | 38.2 (2.6) |
| median (IQR) | 39 (38-40) | 39 (38-40) | 39 (37-40) | 38 (37-40) | 39 (37-40) |
| Preterm (<37 weeks), n (%) | 72,901 (7.5) | 67,790 (7.2) | 3,909 (15.0) | 2,753 (19.7) | 5,111 (15.7) |
| Small for gestational age, n (%) | 96,420 (9.9) | 90,125 (9.6) | 4,974 (19.1) | 3,091 (22.1) | 6,295 (19.3) |
| Admitted to Neonatal ICU or special care nursery,<sup>3</sup> n (%) | 113,168 (14.6) | 104,803 (14.0) | 6,567 (30.4) | 4,827 (42.4) | 8,365 (31.1) |
| **Mother characteristics at birth** |  |  |  |  |  |
| Mother's age (years) at birth of cohort child,<sup>4</sup> mean (SD) | 30.4 (5.6) | 30.6 (5.5) | 26.5 (6.6) | 27.8 (6.4) | 26.8 (6.6) |
| median (IQR) | 31 (27-34) | 31 (27-34) | 26 (21-31) | 27 (23-33) | 26 (21-32) |
| Mothers age at delivery (years) |  |  |  |  |  |
| <20 | 27,512 (2.8) | 22,726 (2.4) | 4,150 (15.9) | 1,382 (9.9) | 4,786 (14.7) |
| 20-24 | 120,840 (12.5) | 112,028 (11.9) | 7,136 (27.4) | 3,401 (24.3) | 8,812 (27.0) |
| 25-29 | 260,485 (26.8) | 252,548 (26.9) | 6,255 (24.0) | 3,672 (26.3) | 7,937 (24.3) |
| 30-34 | 328,250 (33.8) | 321,960 (34.3) | 4,833 (18.6) | 3,121 (22.3) | 6,290 (19.2) |
| 35+ | 233,370 (24.0) | 228,548 (24.4) | 3,671 (14.1) | 2,399 (17.2) | 4,822 (14.8) |
| Mother was born in Australia, n (%) | 634,493 (65.4) | 604,849 (64.5) | 23,798 (91.4) | 12,671 (90.7) | 29,644 (90.8) |
| Aboriginal and/or Torres Strait Islander, n (%) | 51,003 (5.3) | 41,914 (4.5) | 7,796 (29.9) | 3,674 (26.3) | 9,089 (27.8) |
| Mother married/de facto,<sup>5</sup> n (%) | 808,753 (83.3) | 795,798 (84.9) | 9,845 (37.8) | 5,538 (39.6) | 12,955 (39.7) |
| Private health insurance/patient,<sup>6</sup> n (%) | 205,866 (21.2) | 205,262 (21.9) | 300 (1.2) | 343 (2.5) | 604 (1.9) |
| Area of residence,<sup>7</sup> n (%) |  |  |  |  |  |
| Lived in major city | 751,815 (77.5) | 732,726 (78.1) | 15,059 (57.8) | 8,719 (62.4) | 19,089 (58.5) |
| Lived in inner regional area | 150,122 (15.5) | 140,645 (15.0) | 7,678 (29.5) | 3,698 (26.5) | 9,477 (29.0) |
| Lived in outer regional area | 47,059 (4.8) | 43,823 (4.7) | 2,630 (10.1) | 1,188 (8.5) | 3,236 (9.9) |
| Lived in remote/very remote area | 4759 (0.5) | 4315 (0.5) | 385 (1.5) | 143 (1.0) | 444 (1.4) |
| Area-level disadvantage,<sup>8</sup> n (%) |  |  |  |  |  |
| Quintile 1 (Most disadvantaged) | 213,028 (22.0) | 200,803 (21.4) | 9,893 (38.0) | 5,102 (36.5) | 12,225 (37.5) |
| Quintile 2 | 177,648 (18.3) | 169,072 (18.0) | 6,982 (26.8) | 3,417 (24.5) | 8,576 (26.3) |
| Quintile 3 | 211,875 (21.8) | 205,204 (21.9) | 5,383 (20.7) | 2,785 (19.9) | 6,671 (20.4) |
| Quintile 4 | 148,809 (15.3) | 145,901 (15.6) | 2,235 (8.6) | 1,415 (10.1) | 2,908 (8.9) |
| Quintile 5 (Least disadvantaged) | 218,740 (22.5) | 216,486 (23.1) | 1,545 (5.9) | 1,245 (8.9) | 2,254 (6.9) |
| History of contact with public housing system,<sup>9</sup> n (%) | 45,315 (4.7) | 34,151 (3.6) | 9,531 (36.6) | 4,810 (34.4) | 11,164 (34.2) |
| **Pregnancy characteristics relating to child's birth** |  |  |  |  |  |
| Number of previous pregnancies, n (%) |  |  |  |  |  |
| Mother had no prior births | 420,220 (43.3) | 409,737 (43.7) | 7,975 (30.6) | 4,511 (32.3) | 10,483 (32.1) |
| Mother had one prior birth | 326,902 (33.7) | 318,947 (34.0) | 6,201 (23.8) | 3,434 (24.6) | 7,955 (24.4) |
| Mother had ≥2 prior births | 222,591 (22.9) | 208,421 (22.2) | 11,842 (45.5) | 6,005 (43.0) | 14,170 (43.4) |
| Number of previous pregnancies, mean (SD) | 0.9 (1.2) | 1.7 (1.8) | 1.8 (1.9) | 1.7 (1.8) | 1.7 (1.8) |
| median (IQR) | 1 (0-1) | 1 (0-1) | 1(0-3) | 1(0-3) | 1(0-3) |
| Mother did not receive antenatal care in first 20 weeks of pregnancy | 102,125 (10.5) | 94,581 (10.1) | 6,256 (24.0) | 3,365 (24.1) | 7,544 (23.1) |
| Mother smoked during pregnancy,<sup>10</sup> n (%) | 34,615 (10.2) | 27,500 (8.4) | 6,053 (65.2) | 3,096 (68.9) | 7,115 (63.2) |

1. The child protection data source includes maternal and other carer substance use; 2. Defined as child or parent identified as Aboriginal on any of the birth records (i.e. perinatal data collection, birth registration or hospital record); 3. Numbers and percents calculated for children born from 2008-2015 (N=776,273) when this variable was collected in the Perinatal data collection, see supplement 7 for more details; 4. The month and year of birth for the mother was obtained from the birth registration, or the Perinatal Data Collection if the birth registration was unavailable, and used to calculate maternal age at childbirth, in years; 5. As recorded on the hospital birth record; 6. As recorded on the hospital birth record for the mother or child; 7. Based on the Accessibility/Remoteness Index of Australia (ARIA+) for the Statistical Local Area of residence at the child's birth; 8. Based on the Index of Relative Socio-economic Advantage and Disadvantage for the Statistical Local Area of residence at the child's birth; 9. Mother is living in public housing or is on the tenancy waitlist at any time 2 years prior to the child's birth. 10. Variable derived from variables recorded in Perinatal Data Collection, indicates if the mother smoked at any time point during pregnancy.
Number of missing or not recorded values: Sex missing 16; Child Aboriginal or Torres Strait Islander missing 18; Low birth weight missing 770; Small for gestation age missing 902; Admitted to neonatal ICU missing 554; Mother's Aboriginal and/or Torres Strait Islander missing 129, not recorded 21,722; Mother born in Australia missing 1,010; Married or in de facto partnership missing 23,997; Private health insurance/patient not recorded 58; Mother's remoteness area of residence <5 missing, not recorded 16,713; Area-level disadvantage not recorded 370; Number of previous pregnancies missing 757; Antenatal care in the first 20 weeks of pregnancy missing 11,675; Smoking during pregnancy missing 327.

### Analysis methods

We calculated the number and percent of children born 2008-2017 with outcomes recorded in: each data source; any data source; any health/death data source (Tables 1, 2); and the 34 most common data source combinations (Figure 1). We calculated the number and percent of children with 1) records of different substance use types (where recorded) and 2) maternal substance use among birth-year cohorts.

**Figure 1.**
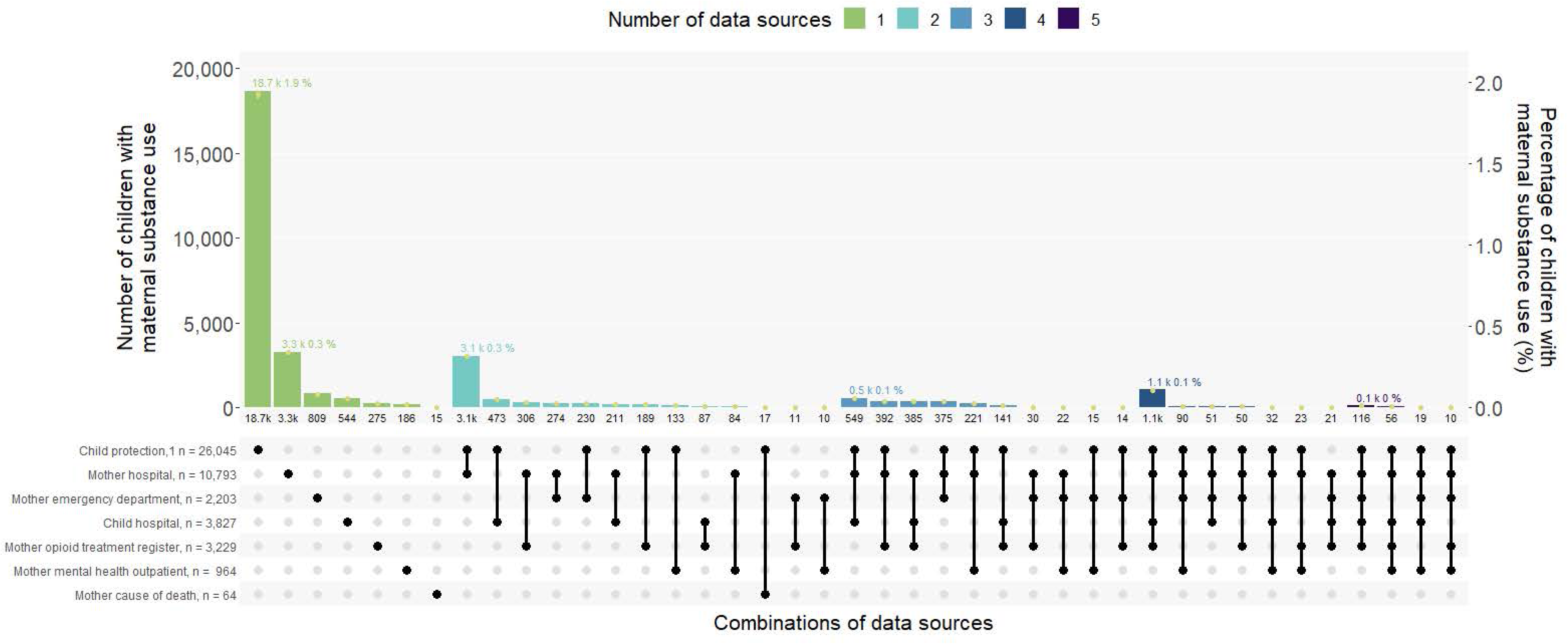
Most common combinations of data sources with maternal substance use recorded, for children born in NSW from 2008-2017 (n=970,470) Footnotes. 122 children were not represented on the upset plot: 93 children were not included as we excluded combinations of data sources with less than 10 children, and 19 children were not included as they had a record of maternal substance use in 6 data sources. All values for numerators, denominators, and 95% confidence intervals provided in supplement 8. 1. The child protection data source includes maternal and other carer substance use.

#### Sensitivity analyses

We conducted sensitivity analyses where the maternal substance use outcome: (i) included additional ICD-10-AM and SNOMED codes that may indicate either the misuse of, or accidental overdose or poisoning from prescription medicines (Supplement 5-5a), or (ii) excluded the receipt of OAT. Whilst OAT is a harm minimisation strategy protective for pregnant women and the developing fetus/infant, exposure to OAT *in utero* may impact the child’s health (e.g., NAS).

## RESULTS

There were 990,631 children born in NSW between January 1, 2008, and December 31, 2017. After excluding 20,150 children who were <20 weeks gestation or <400 grams birth weight, and 21 children with a death record prior to birth, 970,470 children and 625,856 mothers were included (Supplement 4). The number of children born each year was relatively constant, with 96,288 in 2008 and 95,725 in 2017 (mean: 97,000 per year). In total, 48.6% of children were female, 6.9% were Aboriginal and/or Torres Strait Islander, 77.5% were born to mothers residing in major cities, and the mean maternal age at birth was 30.4 years (standard deviation 5.6 years) (Table 1).

### Prevalence of maternal substance use ascertained from mother/child records in u1 data sources

We found 32,647 (3.4%) children had ≥1 mother and/or child records of maternal substance use during the first 1,000 days in ≥1 of the six data sources, and no substance use records for 937,823 children (96.6%) (Table 2). Figure one shows us that, 23,490 children (2.4%) had maternal substance use recorded in only one data source and 10,254 children (1.1%) in ≥2 data sources (Figure 1). There were child protection records of maternal and other carer substance use for 26,045 children (2.7%), including 18,672 (1.9%) children with substance use only recorded in the child protection data. 13,975 children (1.4%) had maternal substance use ascertained from child and/or mother hospital records (Table 2), including 3,283 children with substance use only in maternal hospital records and 544 only in child hospital records (Figure 1). An additional 1,310 children had maternal substance use recorded in data sources other than hospital or child protection (Supplement 9).

**Table 2.** Number and percentage of children with maternal substance use in the first 1000 days, ascertained from six data sources, for children born in NSW from 2008-2017 (N=970,470)

| Data source |  |  |  |  | Single data sources |  |  |  |  |  |  |  | Combined data sources |  |  |  |
| --- | --- | --- | --- | --- | --- | --- | --- | --- | --- | --- | --- | --- | --- | --- | --- | --- |
|  | Cause of death<sup>1</sup> |  | Mental health outpatients |  | Opioid treatment register |  | Emergency department |  | Hospital |  | Child protection<sup>2</sup> |  | Health +/or cause of death<sup>3</sup> |  | Any data source<sup>3</sup> |  |
| Substance type | n | %<sup>4</sup> | n | %<sup>4</sup> | n | %<sup>4</sup> | n | %<sup>4</sup> | n | %<sup>4</sup> | n | %<sup>4</sup> | n | %<sup>4</sup> | n | %<sup>4</sup> |
| **Maternal data sources (5 data sources)** |  |  |  |  |  |  |  |  |  |  |  |  |  |  |  |  |
| Any substance<sup>5</sup> | <64 | <0.1 | 964 | 0.1 | 3,229 | 0.3 | 2,206 | 0.2 | 10,793 | 1.1 | - | - | - | - | 12,956 | 1.3 |
| Alcohol | - | - | 511 | <0.1 | - | - | 1,431 | 0.1 | 3,046 | 0.3 | - | - | - | - | 4,246 | 0.4 |
| Other drug<sup>6</sup> | - | - | 473 | <0.1 | 3,229 | 0.3 | 817 | 0.1 | 8,985 | 0.9 | - | - | - | - | 10,126 | 1.0 |
| Cannabis | - | - | 107 | <0.1 | - | - | 43 | <0.1 | 4,435 | 0.5 | - | - | - | - | 4,505 | 0.5 |
| Opioid or opioid treatment | - | - | 30 | <0.1 | 3,229 | 0.3 | 144 | <0.1 | 3,063 | 0.3 | - | - | - | - | 4,011 | 0.4 |
| Stimulant | - | - | 102 | <0.1 | - | - | 56 | <0.1 | 2,117 | 0.2 | - | - | - | - | 2,182 | 0.2 |
| Sedatives or other<sup>7</sup> | - | - | 6 | <0.1 | - | - | 10 | <0.1 | 549 | 0.1 | - | - | - | - | 572 | 0.1 |
| Unspecified or multiple<sup>8</sup> | - | - | 342 | <0.1 | - | - | 618 | 0.1 | 1,858 | 0.2 | - | - | - | - | 2,576 | 0.3 |
| **Child data sources (3 data sources)** |  |  |  |  |  |  |  |  |  |  |  |  |  |  |  |  |
| Any substance<sup>5</sup> | <5 | <0.1 | - | - | - | - | - | - | 3,827 | 0.4 | 26,045 | 2.7 | - | - | 27,327 | 2.8 |
| Alcohol | - | - | - | - | - | - | - | - | 62 | <0.1 | 10,702 | 1.1 | - | - | 10,726 | 1.1 |
| Other drug<sup>6</sup> | - | - | - | - | - | - | - | - | 3,787 | 0.4 | 18,152 | 1.9 | - | - | 19,559 | 2.0 |
| **Child and maternal data sources combined (6 data sources)<sup>3</sup>** |  |  |  |  |  |  |  |  |  |  |  |  |  |  |  |  |
| Any substance<sup>5</sup> | 64 | <0.1 | 964 | 0.1 | 3,229 | 0.3 | 2,203 | 0.2 | 12,067 | 1.2 | 26,045 | 2.7 | 13,975 | 1.4 | 32,647 | 3.4 |
| Alcohol | - | - | 511 | <0.1 | - | - | 1,431 | 0.1 | 3,074 | 0.3 | 10,702 | 1.1 | 11,175 | 1.2 | 13,637 | 1.4 |
| Other drug<sup>6</sup> | - | - | 473 | <0.1 | 3,229 | 0.3 | 814 | 0.1 | 10,282 | 1.1 | 18,152 | 1.9 | 4,272 | 0.4 | 23,485 | 2.4 |

1. For cause of death data, reporting is limited to the outcome "any substance use" due to low numbers in specific substance types; 2. The child protection data source includes maternal and other carer substance use; 3. The outcomes are not mutually exclusive across data sources or substance types; therefore, numbers in rows/columns do not sum to numbers reported in "All records and data sources" (rows) or the "Any substance use" outcome (columns); 4. 95% confidence intervals provided in supplement 8; 5. Any substance use includes the use of alcohol and or any drug use; 6. Other drug use includes the use of drugs other than alcohol and tobacco, including illicit drugs, misuse of prescription drugs, and use of opioid agonist treatment; 7. "Sedatives or other" includes the use of sedatives, hallucinogens, sedatives, solvents, or organic compounds, which were aggregated due to small cell sizes in individual substance categories; 8. "Unspecified or multiple substance use" includes ICD10AM codes that indicate multiple drug use or do not specify the type of drug or psychoactive substance.

### Prevalence of maternal substance use types

Using child and/or mother data sources, there were records indicating maternal alcohol use for 13,637 children (1.4%) and other drug use for 23,485 children (2.4%) (Table 2). Maternal data sources provided additional details on drug use types. From mother health/death records, there were records indicating maternal alcohol use for 4,246 children (0.4%) and other drug use for 10,126 children (1.0%). Cannabis was the most common type of other drug use ascertained (4,505 children, 0.5%), followed by opioids and OAT (4,011 children, 0.4%). “*Unspecified or multiple drug use”* was recorded in mothers’ health records for 2,117 (0.2%) children. Single substance use was recorded more frequently than multiple use: 8,734 children had 1 and 3,087 children ≥2 substance/s recorded in their mother’s records (Supplement 10-11).

### Annual maternal substance use prevalence between 2008 and 2017

The prevalence of maternal substance use ascertained from ≥1 health/death data sources in the first 1,000 days was relatively stable, ranging from 1.7% in 2008 to 1.3% in 2017 (Figure 2). Consistent with the substantial decrease in all child protection reports following system-wide changes to reporting and screening in 2010,^36^ there was a corresponding decrease in reports related to maternal and other carer substance use from 4.7% in 2008 to 3.1% in 2010, and 3.0% in 2017. On average, each year during the study period, approximately 3,400 children had ≥1 record of maternal substance use, 1,400 alcohol use, and 2,400 other drug use.

**Figure 2.**
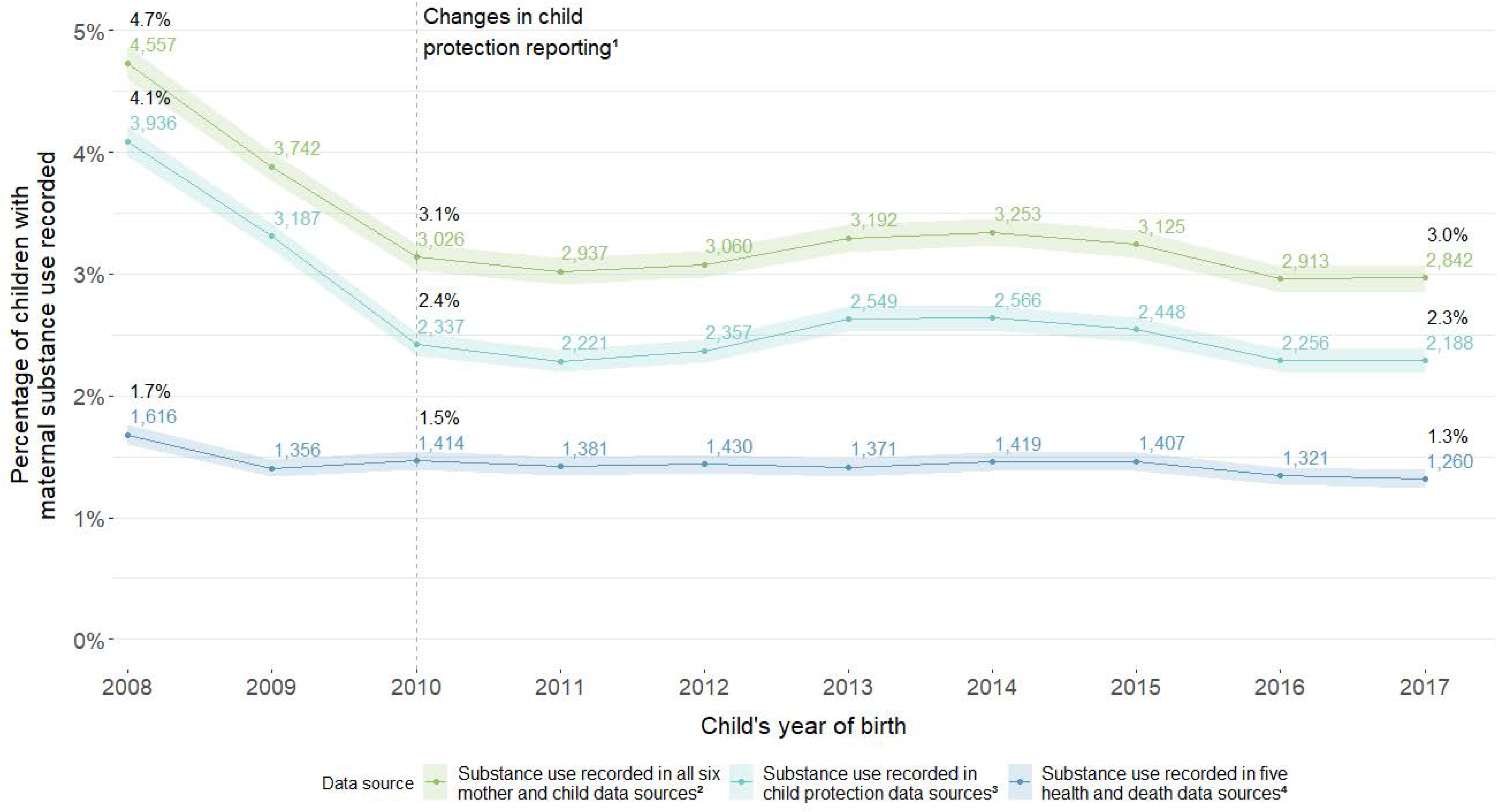
Children with maternal substance use in the first 1000 days of life, ascertained from different combinations of administrative data sources, by child’s year of birth. Footnotes: Total denominator for each birth year ranged from 95,725 to 99,458, mean 97,047 (SD 1134), all denominators, numerators, percentages and 95% confidence intervals provided in supplementary table 12; 1. The Keep Them Safe Initiative and related changes to mandatory reporting guidelines in 2010 in NSW reduced the numbers of child protection reports across all age groups from 2010. 2. Substance use ascertained from maternal hospital, cause of death, mental health outpatients, opioid treatment registry and emergency department records, and child hospital, cause of death and child protection records; 3. The child protection data source includes maternal and other carer substance use; 4. Substance use ascertained from maternal hospital, cause of death, mental health outpatients, opioid treatment registry, and emergency department records, and child hospital and cause of death records.

### Maternal substance use prevalence ascertained during the prenatal and early-life periods

In the prenatal period, 13,987 (1.4%) children had records of maternal substance use, including 3,776 (0.4%) with maternal alcohol use and 11,605 (1.2%) other drug use (Figure 3). In the early-life period (i.e. 28 days to 2^nd^ birthday), 26,198 (2.7%) children had records of maternal substance use, including 11,262 (1.2%) with alcohol use and 17,479 (1.8%) other drug use.

**Figure 3.**
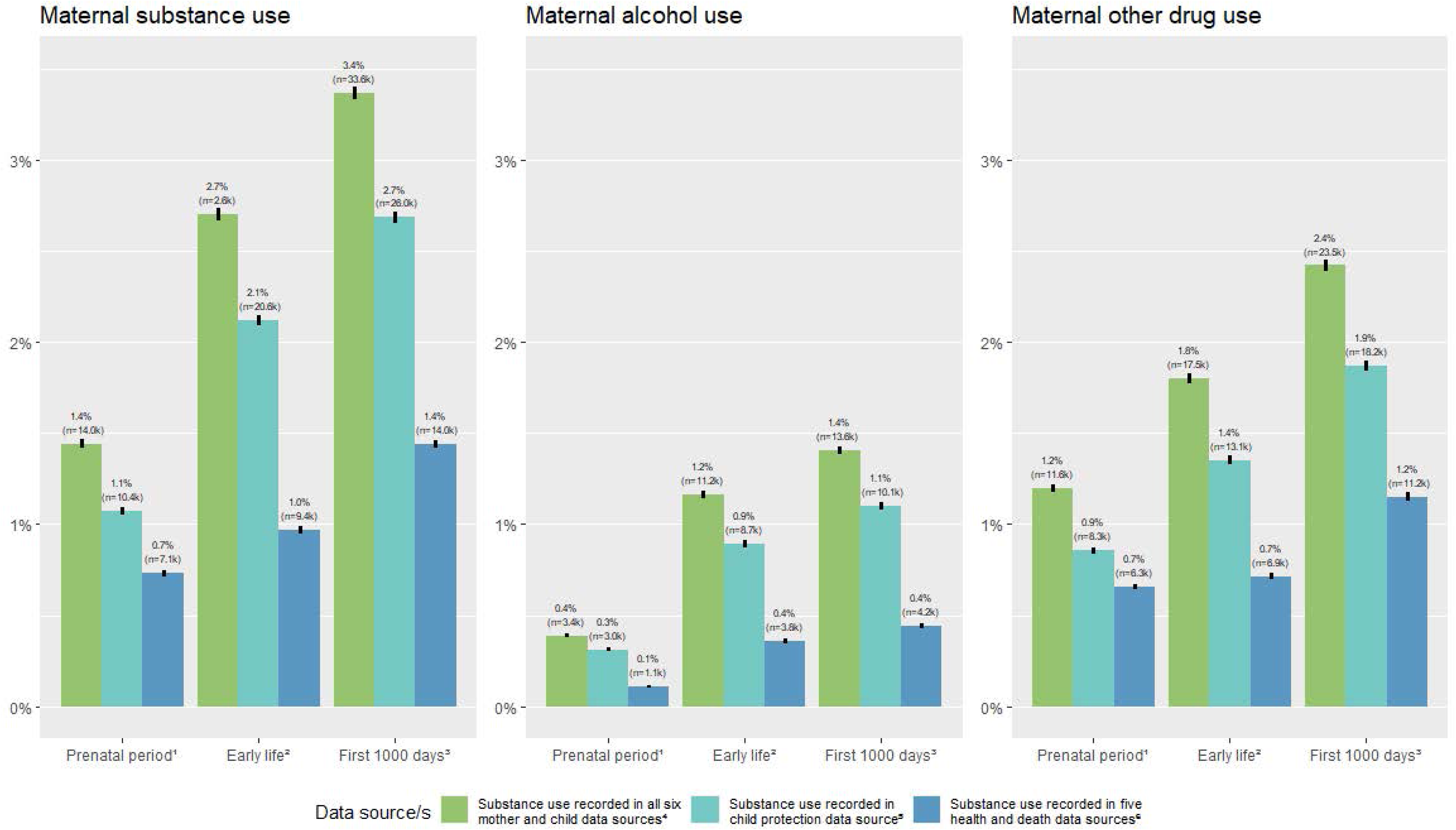
Maternal substance use during the prenatal period, early life, and first 1000 days, among 970,470 children born in New South Wales, from 2008-2017. Footnotes: 1. Prenatal period defined as conception until 27 days post-birth; 2. Early life defined as 28 days post-birth until two years post-birth; 3. Frist 1000 days defined as conception until two years post-birth; 4. Substance use ascertained from maternal hospital, cause of death, mental health outpatients, opioid treatment registry and emergency department records, and child hospital, cause of death and child protection records; 5. The child protection data source includes maternal and other carer substance use; 6. Substance use ascertained from maternal hospital, cause of death, mental health outpatients, opioid treatment registry, and emergency department records, and child hospital and cause of death records.

### Sociodemographic and health profile of children and mothers, according to maternal substance use

Adverse pregnancy and birth characteristics were more common among children with maternal substance use recorded than without, including maternal smoking during pregnancy, low birth weight, preterm birth, and admission to neonatal intensive/special care units (Table 1). Indicators of socioeconomic disadvantage were more common among children with than without records of maternal substance use, including younger maternal age at birth (e.g., <20 years: 14.7% vs. 2.4%), living in more disadvantaged areas (e.g., most disadvantaged: 37.4% vs. 21.4%), and single mother at birth (60.3% vs. 15.1%). Aboriginal and Torres Strait Islander children and mothers were overrepresented in the maternal substance use group (37.5% versus 5.9%).

### Sensitivity analyses

When we included diagnosis codes that potentially indicated misuse of, or accidental overdose/poisoning from, prescription medicines, 236 children were added to the maternal substance use group, with the prevalence remaining 3.4% (Supplement 13).

Almost all children (91.5%) with a mother on the opioid treatment register also had a record of maternal substance use in another data source (Figure 1). When we excluded OAT from the outcome, the prevalence of maternal substance use reduced from 3.36% to 3.34%, representing 275 less children (Supplement 14).

## DISCUSSION

Our study of almost one million Australian children showed that more than 3 in every 100 children had a maternal substance use record during their first 1,000 days of life. This included >1 in 100 children with prenatal substance use exposure and almost 3 in 100 children with maternal substance use records from 28-days to 2-years of age. More than 1 in 100 children had a record of maternal alcohol use and >2 in 100 children a record of other drug use. Health and social disadvantage at birth was more common among children with records of maternal substance use, compared with other children.

Adding child protection data to health and death data more than doubled our prevalence estimates, from 1.4% to 3.4%. While we consider these child protection measures provide a useful additional indicator of maternal substance use, in some cases, they may represent suspected harm relating to the substance use of a non-maternal carer. Nevertheless, children with exposure to family substance use are also at potential risk of harm during early life and important targets of support services. After child protection data, the next highest numbers of children with maternal substance use were ascertained from mother, then child hospital records, which includes near-universal health system contacts at birth. Our prevalence estimates from mother/child hospital records during pregnancy and birth were comparable to previous studies using whole-of-population mother/child hospital data conducted in Australia^8, 10, 11, 24, 27, 29, 37-39^ and other high-income countries over the last two decades.^11-14, 28, 40^ We followed children beyond birth, revealing that almost 3 in 100 had maternal substance use records between 28 days and their second birthday.

Whilst we found fewer children had records of maternal alcohol use than other drug use, another whole-of-population cohort study in Canada reported the opposite (during-pregnancy alcohol use 11.7% versus illicit drug use 3.6%) when using prenatal screening data in addition to hospital data.^18^ This difference likely reflects the different levels of substance use measured in different healthcare contexts and data. Similar to general population surveys, universal prenatal screening uses self-report and thus is likely to capture lower-level and/or intermittent alcohol consumption, whereas child protection and hospital data are likely to capture more visible and harmful alcohol consumption. It is also documented that substance use disorders are underreported in hospital records, with alcohol use disorders underreported more than other drug disorders.^38^

A strength of our study was the use of comprehensive health, death, and child protection data sources spanning 10 years. Using administrative data enabled us to assemble birth cohorts highly representative of our target population, avoiding selection issues common in primary data collection studies. However, using administrative data limited the ascertainment of substance use to what was observed, reported, treated and/or recorded in health system or child protection contacts. In our study, we ascertained a broad spectrum of substance use that resulted in tertiary health care contact; from records potentially indicating harmful use (e.g., diagnoses of drug dependence), one off, infrequent use (e.g., emergency presentations for intoxication), as well as the receipt of substance use treatment (mental health appointments and OAT). We also cannot understand how well mothers are supported by their health care professionals and other services from our results. If mothers are supported, can function as a parent, and are not breastfeeding or around their child while using/affected, the risks to the child may be low. However, we have likely captured more harmful substance use from most data sources, as substance use resulted in death or contact with the child protection or tertiary health systems. Furthermore, the association of maternal substance use with socioeconomic and health disadvantage, indicate that substance use remains a signal that these pregnant women and their children may benefit from support services.

Other data, such as data from antenatal screening and primary care settings, may capture information on lower levels of substance use, as well as more universal health system contacts. In our study only birth admissions in hospital data, represented universal service contacts for all mothers and children. All data sources were collected for statewide services, but most were limited to those mothers and/or children in contact with a service (e.g., emergency department or publicly funded mental health services). However, the additional children ascertained from child protection data may represent children who were visible to and reported from other health and human services. We describe other considerations related to data source-specific coding and ascertainment in supplement 16.

Another important consideration when using administrative data, is that different population groups may be screened or surveilled differently for maternal substance use. Studies suggest women who present late to antenatal care, are younger,^41, 42^ or among racial and ethnic minority populations^42, 43^ are more likely to be screened for substance use. We also found a higher proportion of younger, single mothers, who did not receive antenatal care <20 weeks had records of maternal substance use. Aboriginal mothers and children were also overrepresented, reflecting the historical and ongoing systemic racism, over-surveillance and intervention in the lives of Aboriginal peoples in Australia across sectors, including health, child protection and social services.^44, 45^ Given substance use can be a response and coping mechanism to adversity, interpersonal and state violence - including intergenerational trauma and child removals-^46^ it is critical to consider culturally safe and trauma-informed responses to maternal substance use, alongside other unmet health and social needs.^44, 47^

More than 3 in 100 children had maternal substance use or related conditions recorded in administrative data during the critical first 1,000 days of development. This period of near universal contact with the health system provides the opportune time to identify and respond to maternal and family substance use early in a child’s life to prevent adverse outcomes. By combining child protection records with health data more traditionally used for public health surveillance, we demonstrated the potential to build a more comprehensive view of maternal substance use at the population-level to inform prevention responses for pregnant women, new mothers, and children at the earliest opportunity. However, as substance use was ascertained from health and child protection records, our estimate (3.4%) likely represents the minimum number of families that may need support for potentially harmful substance use. Children and families with evidence of maternal substance use experienced a disproportionate burden of health and social disadvantage, for example 4 in 10 children with a record of maternal substance use live in the most disadvantaged areas. Pregnant women and new mothers using substances may need support from antenatal, birth and postnatal health services, that is coordinated with other mainstream and Aboriginal community-controlled services, including alcohol and other drug services, mental health services, early childhood health services, and other social services such as housing and child protection.

## CONCLUSION

More than 3 in every 100 Australian children had a record of maternal substance use or related conditions in administrative data sources during the first 1000 days of life in this decade-long study. In addition to health and death data sources, child protection data offers public health insights into the scale of maternal substance use among whole-population cohorts of children. The higher burden of health and social disadvantage among children and families affected by maternal substance use during pregnancy and early life highlights the need for coordinated health and social supports from the earliest opportunity.

## Acknowledgement

We thank the children and families of New South Wales (NSW) whose data are included in this study. We thank the NSW Centre for Health Record Linkage for managing and conducting the data linkage for the NSW Child E-Cohort Project (led by KF). We also thank the NSW Ministry of Health, NSW Register of Births, Deaths and Marriages, NSW Department of Education, NSW Department of Communities and Justice for use of the other data sources in this study. We thank the Families and Community Services Insights, Analysis and Research (FACSIAR), Department of Communities and Justice, for their review and feedback on the manuscript. We thank staff at the Aboriginal Health and Medical Research Council (AH&MRC) of New South Wales and the NSW Child, Family and Community Peak Aboriginal Corporation (AbSec) for their input on the study and manuscript, including Sally Cowling, Annaliese Gielingh and Shiny Varghese. We thank staff at the Centre for Alcohol and Other Drugs, NSW Ministry of Health for input and discussion of study findings. The findings and views reported in this study are those of the authors and should not be attributed to any agency or government department.

## Study Registration

Not applicable.

## Contribution Statements

Madeleine Powell conceptualised and designed the study, built the analytic datasets, conducted the data analysis, drafted and revised the manuscript.

Dr Falster conceptualised and designed the study, led data acquisition, supervised data analysis, had input into the initial manuscript, and critically reviewed and revised the manuscript for important intellectual content.

Dr Havard conceptualised and designed the study, supervised data analysis, had input into the initial manuscript, and critically reviewed and revised the manuscript for important intellectual content.

Dr Pilkington contributed to the designed the study, supervised data analysis, and interpretation of the results, and critically reviewed and revised the manuscript for important intellectual content.

Tasnia Ahmed and Dr Hanly supervised and assisted with the build of the analytic datasets, contributed coding and statistical expertise to the analysis, interpretation and reporting of results, and critically reviewed and revised the manuscript for important intellectual content.

Dr Dobbins contributed to the designed the study, supervised data analysis, and interpretation of the results, and critically reviewed the manuscript for important intellectual content.

Professor Lynch. contributed epidemiological and content expertise to the interpretation and reporting of results, and critically reviewed and revised the manuscript for important intellectual content.

Dr Newton contributed content expertise to the interpretation and reporting of results, and critically reviewed and revised the manuscript for important intellectual content.

Dr Michelle Cretikos contributed epidemiological and policy expertise to the interpretation and reporting of results, and critically reviewed and revised the manuscript for important intellectual content.

Dr Jess Stewart contributed policy expertise to the interpretation and reporting of results, and critically reviewed and revised the manuscript for important intellectual content.

All authors approved the final manuscript as submitted and agree to be accountable for all aspects of the work.

## Ethics approval

This study was approved by the NSW Population and Health Services Research Ethics Committee (2020/ETH01265), the University of NSW HREC (2020/ETH01265), the Aboriginal Health and Medical Research Council (AH&MRC) of NSW Ethics Committee (1688/20), the NSW Corrective Services Ethics Committee (D20/0886760). The CHeReL operate under strict data security protocols and implements high level physical security measures. Their security protocols are in accordance with the Australian Government Protective Security Policy Framework, the Population Health Research Network Information Governance Framework, and the NHMRC Code for Responsible Conduct of Research.

## Data availability statement

No data are available because Australian privacy laws prohibit us from making the individual-level de-identified data publicly available. The data used for this study were provided by several Australian State and Commonwealth government agencies under agreements with the researchers led by KF (in NSW) and JWL (in SA), the NSW Centre for Health Record Linkage (CHeReL) and SANT Datalink, following approval from multiple ethics committees and data custodians. Data are only able to be accessed by researchers who are approved users by the relevant ethics committees and data custodians. Data can be accessed through an application and approval process administered by the independent data linkage authorities, NSW CHeReL or SANT Datalink.

## Supplementary data

Supplementary data are available at *Addiction* online

