## Supplement for "Prevalence of maternal substance use during pregnancy and first two years of life: A whole-population cohort of 970,470 Australian children born 2008-2017"

**Supplement 1. RECORD statement – checklist of items that should be reported in observational studies using routinely collected health data.**

| | Item No. | STROBE items | RECORD items | Location in manuscript |
|---|---|---|---|---|
| **Title and abstract** | | | | |
| | 1 | (a) Indicate the study's design with a commonly used term in the title or the abstract (b) Provide in the abstract an informative and balanced summary of what was done and what was found | RECORD 1.1: The type of data used should be specified in the title or abstract. When possible, the name of the databases used should be included.<br><br>RECORD 1.2: If applicable, the geographic region and timeframe within which the study took place should be reported in the title or abstract.<br><br>RECORD 1.3: If linkage between databases was conducted for the study, this should be clearly stated in the title or abstract. | Title page 1. Abstract page 3. |
| **Introduction** | | | | |
| Background rationale | 2 | Explain the scientific background and rationale for the investigation being reported | | Introduction Paragraph 1&2 |
| Objectives | 3 | State specific objectives, including any prespecified hypotheses | | Introduction paragraph 3 |
| **Methods** | | | | |
| Study Design | 4 | Present key elements of study design early in the paper | | Methods |
| Setting | 5 | Describe the setting, locations, and relevant dates, including periods of recruitment, exposure, follow-up, and data collection | | Study population paragraph, Supplement 4 |
| Participants | 6 | *(a) Cohort study* - Give the | RECORD 6.1: The methods of study | |
| | | eligibility criteria, and the sources and methods of selection of participants. Describe methods of follow-up<br>*Case-control study* - Give the eligibility criteria, and the sources and methods of case ascertainment and control selection. Give the rationale for the choice of cases and controls *Cross-sectional study* - Give the eligibility criteria, and the sources and methods of selection of participants<br><br>*(b) Cohort study* - For matched studies, give matching criteria and number of exposed and unexposed<br>*Case-control study* - For matched studies, give matching criteria and the number of controls per case | population selection (such as codes or algorithms used to identify subjects) should be listed in detail. If this is not possible, an explanation should be provided.<br><br>RECORD 6.2: Any validation studies of the codes or algorithms used to select the population should be referenced. If validation was conducted for this study and not published elsewhere, detailed methods and results should be provided.<br><br>RECORD 6.3: If the study involved linkage of databases, consider use of a flow diagram or other graphical display to | Study population, Supplement 3 & 4 |

| | Item No. | STROBE items | RECORD items | Location in manuscript |
|---|---|---|---|---|
| | | | demonstrate the data linkage process, including the number of individuals with linked data at each stage. | |
| Variables | 7 | Clearly define all outcomes, exposures, predictors, potential confounders, and effect modifiers. Give diagnostic criteria, if applicable. | RECORD 7.1: A complete list of codes and algorithms used to classify exposures, outcomes, confounders, and effect modifiers should be provided. If these cannot be reported, an explanation should be provided. | Outcomes, Sociodemographic and health characteristics of mothers and children at birth, Supplement 4, 5, Supplement ICD10-AM and SNOMED CT code definitions |
| Data sources/ measurement | 8 | For each variable of interest, give sources of data and details of methods of assessment (measurement).<br>Describe comparability of assessment methods if there is more than one group | | Data sources and data linkage, Outcomes, Supplement 4, 5, Supplement ICD10-AM and SNOMED CT code definitions |
| Bias | 9 | Describe any efforts to address potential sources of bias | | Outcomes, Sensitivity Analysis |
| Study size | 10 | Explain how the study size was arrived at | | Study population, Supplement 3 & 4 |
| Quantitative variables | 11 | Explain how quantitative variables were handled in the analyses. If applicable, describe which groupings were chosen, and why | | Analysis methods, Outcomes |
| Statistical methods | 12 | (a) Describe all statistical methods, including those used to control for confounding (b) Describe any methods used to examine subgroups and interactions<br>(c) Explain how missing data were addressed<br>(d) *Cohort study* - If applicable, explain how loss to follow-up<br>was addressed<br>*Case-control study* - If applicable, explain how matching of cases and controls<br>was addressed | | Analysis methods |

| | Item No. | STROBE items | RECORD items | Location in manuscript |
| --- | --- | --- | --- | --- |
| | | *Cross-sectional study* - If applicable, describe analytical methods taking account of sampling strategy<br>(e) Describe any sensitivity analyses | | |
| Data access and cleaning methods | | .. | RECORD 12.1: Authors should describe the extent to which the investigators had access to the database population used to create the study population.<br><br>RECORD 12.2: Authors should provide information on the data cleaning methods used in the study. | Methods, Data source and data linkage, study population, Supplement 4 |
| Linkage | | .. | RECORD 12.3: State whether the | |
| | | | study included person-level, institutional-level, or other data linkage across two or more databases. The methods of linkage and methods of linkage quality evaluation should be provided. | Data sources and data linkage |
| Participants | 13 | (a) Report the numbers of individuals at each stage of the study (*e.g.*, numbers potentially eligible, examined for eligibility, confirmed eligible, included in the study, completing follow-up, and analysed)<br>(b) Give reasons for nonparticipation at each stage. (c) Consider use of a flow diagram | RECORD 13.1: Describe in detail the selection of the persons included in the study (*i.e.,* study population selection) including filtering based on data quality, data availability and linkage. The selection of included persons can be described in the text and/or by means of the study flow diagram. | Study population, Supplement 3 & 4 |
| Descriptive data | 14 | (a) Give characteristics of study participants (*e.g.*, demographic, clinical, social) and information on exposures and potential confounders<br>(b) Indicate the number of participants with missing data for each variable of interest (c) *Cohort study* - summarise follow-up time (*e.g.*, average and total amount) | | Results paragraph 1, Table 1, Results: Sociodemographic and health profile of children and mothers, according to maternal substance use |
| Outcome data | 15 | *Cohort study* - Report numbers of outcome events or summary measures over time<br>*Case-control study* - Report numbers in each exposure category, or summary measures of exposure<br>*Cross-sectional study* - Report numbers of outcome events or | | Table 2, Figure 1-3, Supplement 7,<br><br>Results paragraph 2-3 |
| | | summary measures | | |

| | Item No. | STROBE items | RECORD items | Location in manuscript |
|---|---|---|---|---|
| Main results | 16 | (a) Give unadjusted estimates and, if applicable, confounder adjusted estimates and their precision (e.g., 95% confidence interval). Make clear which confounders were adjusted for and why they were included (b) Report category boundaries when continuous variables were categorized<br>(c) If relevant, consider translating estimates of relative risk into absolute risk for a meaningful time period | | Table 2, Figure 1-3, Supplement 7-12 Results paragraph 2-5 |
| Other analyses | 17 | Report other analyses done—e.g., analyses of subgroups and interactions, and sensitivity analyses | | Supplement 13-15, Results: Sensitivity analyses |
| Key results | 18 | Summarise key results with reference to study objectives | | Discussion paragraph 1 and 2 |
| Limitations | 19 | Discuss limitations of the study, taking into account sources of potential bias or imprecision. Discuss both direction and magnitude of any potential bias | RECORD 19.1: Discuss the implications of using data that were not created or collected to answer the specific research question(s). Include discussion of misclassification bias, unmeasured confounding, missing data, and changing eligibility over time, as they pertain to the study being reported. | Discussion Paragraph 2-6 |
| Interpretation | 20 | Give a cautious overall interpretation of results considering objectives, limitations, multiplicity of analyses, results from similar studies, and other relevant evidence | | Discussion paragraph 7 |
| Generalisability | 21 | Discuss the generalisability (external validity) of the study results | | Discussion Paragraph 4, 5, 6 |
| Funding | 22 | Give the source of funding and the role of the funders for the present study and, if applicable, for the original study on which the present article is based | | Funding and acknowledgments |
| Accessibility of protocol, raw data, and programming code | | .. | RECORD 22.1: Authors should provide information on how to access any supplemental information such as the study protocol, raw data, or programming code. | NA |

*Reference: Benchimol EI, Smeeth L, Guttmann A, Harron K, Moher D, Petersen I, Sørensen HT, von Elm E, Langan SM, the RECORD Working Committee. The REporting of studies Conducted using Observational Routinely-collected health Data (RECORD) Statement. *PLoS Medicine* 2015; in press.

**Supplement 2. Data sources included in the analysis, with technical names and the years data sources are available**

| Data sources[1] | Pregnancy | Birth | Neonatal period | First 1000 days | Years data are available |
|---|---|---|---|---|---|
| **Mother's records** | | | | | |
| NSW Perinatal Data Collection (formerly known as the Midwives Data Collection) (Perinatal data) | | | | | 2001-2019 |
| NSW Registry of Births, Deaths, and Marriages birth registrations (Birth registrations) | | | | | 2001-2019 |
| NSW Admitted Patient Data Collection (Hospital inpatient data) | | | | | Jul 2001-2019 |
| NSW Emergency Department Data Collection (Emergency department presentations) | | | | | 2005 -2019 |
| NSW Mental Health Ambulatory Data Collection (Mental health outpatient data) | | | | | 2006-2019 |
| Registry of Births, Deaths, and Marriages (RBDM) death registrations | | | | | 2001 -2019 |
| NSW Cause of Death Unit Record File Unit Record File (Cause of death records) | | | | | 2001-2019 |
| NSW Department of Communities and Justice, Social housing applicant file and Tenancy file (Public housing data) | | | | | 2001-2019 |
| NSW Controlled Drugs Data Collection - Opioid Treatment Program (Opioid treatment registry) | | | | | 2001-2019 |
| **Child's records** | | | | | |
| NSW Perinatal Data Collection (formerly known as the Midwives Data Collection) (Perinatal data) | | | | | 2001-2019 |
| NSW Registry of Births, Deaths, and Marriages birth registrations (Birth registrations) | | | | | 2001-2019 |
| NSW Admitted Patient Data Collection (Hospital inpatient data) | | | | | Jul 2001-2019 |
| Registry of Births, Deaths, and Marriages (RBDM) death registrations (Beath registrations) | | | | | 2001 -2019 |
| NSW Cause of Death Unit Record File Unit Record File (Bause of death records) | | | | | 2001-2019 |
| NSW Department of Communities and Justice, child protection reports and investigations (Child protection data) | | | | | 2004 -2019 |

1. More information on all data sources can be found at https://www.cherel.org.au/datasets Definitions: NSW: New South Wales

**Supplement 3. Study population and years data available for each birth cohort**

| Birth year | 2001 | 2002 | 2003 | 2004 | 2005 | 2006 | 2007 | 2008 | 2009 | 2010 | 2011 | 2012 | 2013 | 2014 | 2015 | 2016 | 2017 | 2018 | 2019 | Years of data |
|---|---|---|---|---|---|---|---|---|---|---|---|---|---|---|---|---|---|---|---|---|
| 2002 | Pregnancy | Birth | 1000 days | | | | | | | | | | | | | | | | | 19 |
| 2003 | | | Birth | | | | | | | | | | | | | | | | | 18 |
| 2004 | | | | Birth | | | | | | | | | | | | | | | | 17 |
| 2005 | | | | | Birth | | | | | | | | | | | | | | | 16 |
| 2006 | | | | | | Birth | | | | | | | | | | | | | | 15 |
| 2007 | | | | | | | Birth | | | | | | | | | | | | | 14 |
| 2008 | | | | | | | | Birth | | | | | | | | | | | | 13 |
| 2009 | | | | | | | | | Birth | | | | | | | | | | | 12 |
| 2010 | | | | | | | | | | Birth | | | | | | | | | | 11 |
| 2011 | | | | | | | | | | | Birth | | | | | | | | | 10 |
| 2012 | | | | | | | | | | | | Birth | | | | | | | | 9 |
| 2013 | | | | | | | | | | | | | Birth | | | | | | | 8 |
| 2014 | | | | | | | | | | | | | | Birth | | | | | | 7 |
| 2015 | | | | | | | | | | | | | | | Birth | | | | | 6 |
| 2016 | | | | | | | | | | | | | | | | Birth | | | | 5 |
| 2017 | | | | | | | | | | | | | | | | | Birth | | | 4 |

Calendar year (follow-up from gestation through to 2 years of age)

Study population = children born from 2008 to 2017

**Years data are available**

NSW Mental Health Ambulatory Data Collection (Mental health outpatients data)

NSW Emergency Department Data Collection (Emergency department presentations)

NSW Department of Communities and Justice, child protection reports and investigations (child protection data)

NSW Admitted Patient Data Collection (Hospital inpatient data)

Registry of Births, Deaths and Marriages death registrations, NSW Cause of Death Unit Record File Unit Record File (cause of death records)

NSW Perinatal Data Collection (formerly known as the Midwives Data Collection) (perinatal data)

NSW Registry of Births, Deaths and Marriages birth registrations (birth registrations)

NSW Controlled Drugs Data Collection - Opioid Treatment Program (opioid treatment registry)

NSW Department of Communities and Justice, Social housing applicant file and Tenancy file (public housing data)

**Supplement 4. Study population assembled from linked data sources**

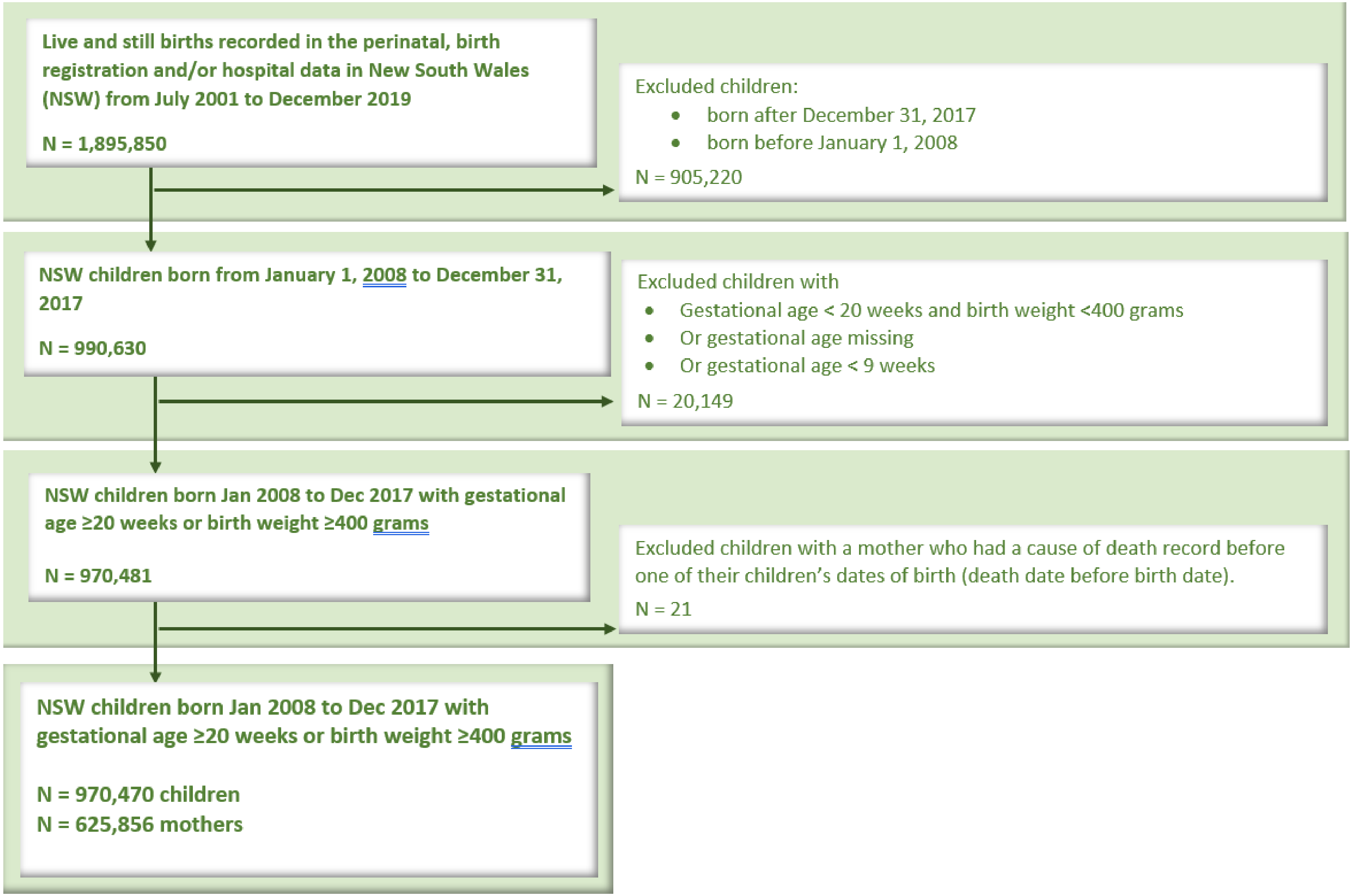

**Supplement 5: Data used to ascertain maternal substance use from six administrative data sources**

<table>
<tr><th>Details of data used</th><th>Codes/variables</th></tr>
<tr><td colspan="2">Hospital records (APDC)[1]</td></tr>
<tr><td rowspan="2">Primary and secondary diagnoses (ICD10-AM diagnostic codes) indicating use of substances in the mother’s records and children’s.</td><td>Mother records:<br>Any substance: Combined measure of other drugs and alcohol.<ul><li>Alcohol: O35.4, Z50.2, Z72.1, R78.0, Y90-91.9, T51-51.9, X45, X65, Y15, F10-10.9</li><li>Other drugs: Combined measure of ICD10-AM codes indicating opioids, cannabis, stimulants (cocaine, amphetamines, other stimulants), hallucinogens, solvents, sedatives and hypnotics, and drug unspecified or multiple drug:<ul><li>Opioids: T40.0-40.3, F11- F11.9</li><li>Cannabis T40.7, F12-12.9</li><li>Stimulants: including cocaine: R78.2, T40.5, F14-14.9, amphetamines: T43.61, T43.62, and other stimulants T43.69, F15-15.99</li><li>Sedatives or other: Sedatives and hypnotics: F13-13.99; Volatile and organic solvents: F18-18.9, T52, T52.0, T52.1, T52.8, T52.9; and Hallucinogens: R78.3, T40.8, T40.9, F16-16.99</li><li>Drug unspecified or multiple: O35.5, Z50.3, Z72.2, R78, F19-19.9, T40, T40.4, T40.6</li></ul></li></ul>Additional codes for sensitivity analysis:<br>Other drugs: Combined measure as above, with the addition of codes which could indicate poisoning from a substance that may be misused, but also could be attributed to the accidental poisoning from prescription medication: T41.22, T42, T42.3, T42.4, T42.6, T42.7, T43, T43.6, T43.60, T43.8, T43.9)</td></tr>
<tr><td>Child records:<br>Any substance: Combined measure of other drugs and alcohol.<ul><li>Alcohol: P04.3, Q86.0</li></ul>Other drugs: P96.1, P04.4</td></tr>
<tr><td colspan="2">Cause of Death records[1]</td></tr>
<tr><td>Underlying Cause of Death Diagnosis Code and Contributing causes of death (ICD-10) diagnosis codes indicating use of substances in the mother’s records and children’s.</td><td>Mother records:<br>Any substance: Combined measure of other drugs and alcohol.<ul><li>Alcohol: O35.4, Z50.2, Z72.1, R78.0, Y90-91.9, T51-51.9, X45, X65, Y15, F10-10.9</li><li>Other drugs: Combined measure of ICD10-AM codes indicating opioids, cannabis, stimulants (cocaine, amphetamines, other stimulants), hallucinogens, solvents, sedatives and hypnotics, and drug unspecified or multiple drug:<ul><li>Opioids: T40.0-40.3, F11- F11.9</li><li>Cannabis T40.7, F12-12.9</li><li>Stimulants: including cocaine: R78.2, T40.5, F14-14.9, amphetamines: T43.61, T43.62, and other stimulants T43.69, F15-15.99</li></ul></li></ul></td></tr>
</table>

| | |
|---|---|
| | • **Sedatives or other:** Sedatives and hypnotics: F13-13.99; Volatile and organic solvents: F18-18.9, T52, T52.0, T52.1, T52.8, T52.9; and Hallucinogens: R78.3, T40.8, T40.9, F16-16.99<br>• **Drug unspecified or multiple:** O35.5, Z50.3, Z72.2, R78, F19-19.9, T40, T40.4, T40.6<br><br>**Additional codes for sensitivity analysis:**<br>*Other drugs: Combined measure as above, with the addition of codes which could indicate poisoning from a substance that may be misused, but also could be attributed to the accidental poisoning from prescription medication: T41.22, T42, T42.3, T42.4, T42.6, T42.7, T43, T43.6, T43.60, T43.8, T43.9)* |
| | Child records:<br>Any substance: Combined measure of other drugs and alcohol.<br>• Alcohol: P04.3, Q86.0<br>• Other drugs: P96.1, P04.4 |
| Mental health outpatients records (MHAMB) [1] | |
| Mental Health Diagnosis Code and additional diagnosis codes (ICD-10) indicating use of substances in the mother's records. | **Mother records:**<br>**Any substance:** Combined measure of other drugs and alcohol.<br>• **Alcohol**: O35.4, Z50.2, Z72.1, R78.0, Y90-91.9, T51-51.9, X45, X65, Y15, F10-10.9<br>• **Other drugs:** Combined measure of ICD10-AM codes indicating opioids, cannabis, stimulants (cocaine, amphetamines, other stimulants), hallucinogens, solvents, sedatives and hypnotics, and drug unspecified or multiple drug:<br> ◦ **Opioids:** T40.0-40.3, F11- F11.9<br> ◦ **Cannabis** T40.7, F12-12.9<br> ◦ **Stimulants:** including cocaine: R78.2, T40.5, F14-14.9, amphetamines: T43.61, T43.62, and other stimulants T43.69, F15-15.99<br> ◦ **Sedatives or other:** Sedatives and hypnotics: F13-13.99; Volatile and organic solvents: F18-18.9, T52, T52.0, T52.1, T52.8, T52.9; and Hallucinogens: R78.3, T40.8, T40.9, F16-16.99<br> ◦ **Drug unspecified or multiple:** O35.5, Z50.3, Z72.2, R78, F19-19.9, T40, T40.4, T40.6<br><br>**Additional codes for sensitivity analysis:**<br>*Other drugs: Combined measure as above, with the addition of codes which could indicate poisoning from a substance that may be misused, but also could be attributed to the accidental poisoning from prescription medication: T41.22, T42, T42.3, T42.4, T42.6, T42.7, T43, T43.6, T43.60, T43.8, T43.9)* |
| Emergency Department Data Collection (EDDC) [1] | |

| | |
|---|---|
| Presenting issue, ICD-10AM and SNOMED codes indicating use of substances in the mother's records in mother's records | **Mother records:**<br>***ICD10-AM***<br>**Any substance:** Combined measure of other drugs and alcohol.<br>• **Alcohol**: O35.4, Z50.2, Z72.1, R78.0, Y90-91.9, T51-51.9, X45, X65, Y15, F10-10.9<br>• **Other drugs:** Combined measure of ICD10-AM codes indicating opioids, cannabis, stimulants (cocaine, amphetamines, other stimulants), hallucinogens, solvents, sedatives and hypnotics, and drug unspecified or multiple drug:<br> o **Opioids:** T40.0-40.3, F11- F11.9<br> o **Cannabis** T40.7, F12-12.9<br> o **Stimulants:** including cocaine: R78.2, T40.5, F14-14.9; amphetamines: T43.61, T43.62; and other stimulants T43.69, F15-15.99<br> o **Sedatives or other:** Sedatives and hypnotics: F13-13.99; Volatile and organic solvents: F18-18.9, T52, T52.0, T52.1, T52.8, T52.9; and Hallucinogens: R78.3, T40.8, T40.9, F16-16.99<br> o **Drug unspecified or multiple:** O35.5, Z50.3, Z72.2, R78, F19-19.9, T40, T40.4, T40.6<br><br>**Additional codes for sensitivity analysis:**<br>*Other drugs: Combined measure as above, with the addition of codes which could indicate poisoning from a substance that may be misused, but also could be attributed to the accidental poisoning from prescription medication: T41.22, T42, T42.3, T42.4, T42.6, T42.7, T43, T43.6, T43.60, T43.8, T43.9)*<br><br>*SNOMED CT*<br>**Any substance:** Combined measure of other drugs and alcohol.<br>• **Alcohol**: 281078001, 64297001, 15167005, 440652002, 20093000, 25702006, 66590003, 191882002, 191883007, 191891003, 228330005, 231467000, 268645007, 284591009, 304605000, 386449006, 713862009, 827094004, 1149333003, 10741871000119100, 4953006, 6749002, 7052005, 7200002, 8635005, 18653004, 21000000, 25966003, 29212009, 32553006, 34938008, 41083005, 42344001, 53936005, 57346004, 61144001, 67426006, 78524005, 79578000, 82047000, 82782008, 85561006, 87460008, 89507002, 95906008, 160592001, 169942003, 191475009, 191476005, 191477001, 191478006, 191480000, 191802004, 191804003, 191805002, 191811004, 191812006, 192206005, 192207001, 192208006, 192209003, 192210008, 192211007, 192212000, 192213005, 192214004, 192215003, 192216002, 207273009, 212806006, 212807002, 212808007, 212809004, 212811008, 212813006, 212814000, 212815004, 212816003, 212817007, 212818002, 212819005, 212820004, 213687005, 216633005, 216635003, 216636002, 216640006, 216645001, 216648004, 216651006, 221843007, 221844001, 221845000, 221846004, 221847008, 221848003, 221849006, 221850006, 221851005, 221852003, 222103001, 222104007, 222105008, 87106005, 222106009, 222107000, 222108005, 222110007, 222111006, 222112004, 222113009, 222114003, 222702003, 222703008, 222704002, 222705001, 222706000, 222707009, 222708004, 222709007, 222710002, 222711003, 222713000, 223333005, 223334004, 223335003, 223336002, 223337006, 223338001, |

<table>
<tr>
<td></td>
<td>
223339009, 223340006, 223341005, 223342003, 223343008, 223344002, 223345001, 223346000, 223347009, 223348004, 223349007, 228310006, 228315001, 228316000, 228317009, 228322009, 228323004, 228341007, 228350009, 228351008, 228353006, 228354000, 228357007, 230800004, 242263000, 242265007, 268639004, 268683008, 268684002, 268685001, 269765000, 274776000, 278363000, 287166006, 300939009, 304606004, 308742005, 315226008, 361267005, 441685000, 442669008, 442764005, 442766007, 713583005, 714829008, 10755041000119100, 288021000119107, 288041000119101, 219006, 24165007, 62213004, 28045007, 35637008, 38670004, 53041004, 53527002, 86933000, 102612005, 102897001, 135827004, 160573003, 160581002, 160593006, 160599005, 163184002, 183098002, 183486001, 191479003, 191481001, 191482008, 191803009, 191807005, 191809008, 191814007, 191815008, 191885000, 212810009, 212812001, 216632000, 216634004, 216637006, 216638001, 216639009, 216643008, 216644002, 216646000, 216649007, 216650007, 216652004, 216653009, 221842002, 228273003, 228281002, 228312003, 228313008, 228326007, 228358002, 228364009, 231464007, 231465008, 274257003, 292880007, 294420000, 302237007, 307730003, 311492009, 316322002, 316494009, 365967005, 365973006, 408945004, 408946003, 408947007, 408948002, 412198003, 413473000, 415685003, 417096006, 417633001, 419442005, 419572002, 420140004, 427013000, 429501006, 429775004, 431260004, 445628007;

- **Other drugs:** Combined measure of SNOMED CT codes indicating opioids, cannabis, stimulants (cocaine, amphetamines, other stimulants), hallucinogens, solvents, sedatives and hypnotics, and drug unspecified or multiple drug:
    - **Opioids or opioid treatment:** 5602001, 52866005, 75544000, 191820008, 191909007, 191912005, 191913000, 231477003, 231478008, 231479000, 231480002, 310653000, 428819003, 429512006, 792901003, 792902005, 1255015004, 1081000119105, 145121000119106, 1515351000168100, 1640611000168100, 1640661000168100, 13187008, 14784000, 60199004, 77721001, 87132004, 191819002, 216463005, 216464004, 242828004, 242829007, 242831003, 290182008, 290183003, 295161000, 295163002, 295164008, 295165009, 295170002, 295171003, 295172005, 295173000, 295174006, 295175007, 295176008, 295184007, 295186009, 426001001, 242253008;
    - **Cannabis:** 37344009, 39807006, 191838006, 191893000, 191894006, 428823006, 737336003, 26714005, 39951001, 63649001, 77355000, 85005007, 268641003, 703848005;
    - **Stimulants:** 21647008, 27956007, 31956009, 78267003, 191831000, 191916008, 191918009, 191919001, 191920007, 296321004, 428493006, 428659002, 429782000, 441527004, 442406005, 699449003, 762504005, 762505006, 762671008, 785277001, 1155772001, 1230071005, 1230072003, 1230082002, 1230084001, 1230085000, 1230086004, 1230087008, 1230088003, 1230089006, 1231319004, 145101000119102, 429001000124103, 12398571000119100, 8837000, 45421006, 46975003, 51493001, 80868005, 84758004, 290544006, 290545007, 296322006, 296323001, 428370001
    - **Sedatives or other:** : 7071007, 38247002, 50320000, 60901005, 64386003, 70340006, 74851005, 105549004, 191849000, 191856006, 191899001, 191900006, 231462006, 231468005, 268646008, 275471001, 426095000, 426590003, 427229002, 428495004, 428623008, 724713006, 724715004, 772999000, 1230074002, 1230077009,
</td>
</tr>
</table>

| | |
|---|---|
| | 1230081009, 1231160001, 86391000119101, 125851000119106, 145841000119107, 1382351000168100, 1382391000168100, 1382421000168100, 1382521000168100, 1382531000168100, 1382541000168100, 1382551000168100, 1382561000168100, 1382571000168100, 1382581000168100, 88320008, 191853003, 231473004;<br>○ **Drug unspecified or multiple:** 2403008, 6525002, 9769006, 23915005, 26416006, 49540005, 56876005, 61480009, 66214007, 70545002, 87106005, 91388009, 95607001, 105546006, 110281001, 182969009, 191934007, 191936009, 191937000, 191938005, 226034001, 228371004, 228372006, 228373001, 228374007, 228375008, 228376009, 228377000, 228378005, 228379002, 228381000, 228382007, 228383002, 228384008, 228386005, 228387001, 228388006, 228389003, 228390007, 228391006, 228394003, 228401002, 228415000, 228421001, 228422008, 228423003, 228425005, 228429004, 266707007, 307052004, 361049005, 361055000, 386450006, 386451005, 414054004, 414056002, 416262000, 416437003, 417252002, 417284009, 424848002, 425533007, 440664005, 442229000, 445273005, 708079007, 713775002, 741063003, 772808000, 1149322001, 1231333004, 1254812006, 1254974001, 1254987007, 1255017007, 1255020004, 1259017004, 1334001000168100, 1545151000168100, 1648321000168100, 1648331000168100, 16076691000119100, 11061003, 11196001, 11387009, 28368009, 32709003, 43242008, 50026000, 51339003, 74934004, 84584008, 191483003, 191484009, 191485005, 191486006, 191492000, 191494004, 191495003, 191496002, 191816009, 191865004, 191873008, 191939002, 228438002, 231451006, 231466009, 231481003, 231482005, 247702001, 290220008, 290221007, 290222000, 295154004, 295167001, 295169003, 295190006, 295193008, 295194002, 295213004, 297199006, 363101005, 363314000, 365984004, 396344000, 416119007, 429299000, 429672007, 442351006, 62213004 |
| Receipt (commencement or continuation) of opioid agonist treatment in the mother's records. | **Mother records:**<br>**Any substance:** *Receipt (commencement or continuation) of opioid agonist treatment during or before the study period/s, individuals with no treatment end date were assumed to be in treatment.*<br>• ***Other drugs:*** Receipt (commencement or continuation) of opioid agonist treatment during or before the study period/s, in*dividuals with no treatment end date were assumed to be in treatment*. |
| Child protection records | |
| Primary and other issue/s reported to the child protection helpline<br>**and**<br>Primary and other assessed Issue/s at field assessment indicating use of substances in the children's records. | Any substance: Combined measure of alcohol and other drugs, and the below variables indicating substance use:<br>• *Reported issues group* and/or *Field assessment group*:<br>○ Drug/alcohol use by carer<br>• *Primary helpline assessed issue, Other helpline assessed issues, Other field assessment assessed issue,* and *Primary field assessment assed issue:*<br>○ drug/alcohol abuse by carer,<br>○ Neglect: exposure to drug/alcohol use<br>○ Parental risk factor: substance use<br>○ Parents substance use led to emotional harm<br>• Alcohol: *Primary helpline assessed issue, Other helpline assessed issues, Other field assessment assessed issue,* and *Primary field assessment assed issue:*<br>○ alcohol abuse by carer |

| | |
|---|---|
| | • **Other drugs:** *Primary helpline assessed issue, Other helpline assessed issues, Other field assessment assessed issue,* and *Primary field assessment assed issue:*<br>o drug abuse by carer |
| Footnotes: 1. Definitions for ICD-10 AM and SNOMED CT codes provided in supplement 15 | |

**Supplement 6. Outcome ascertainment periods**

| | Name | Definition |
|---|---|---|
| 1 | First 1000 days | Conception to child's 2nd birthday |
| 2 | Prenatal period | Conception to <28 days post birth |
| 3 | Early life | 28 days to child's 2nd birthday |

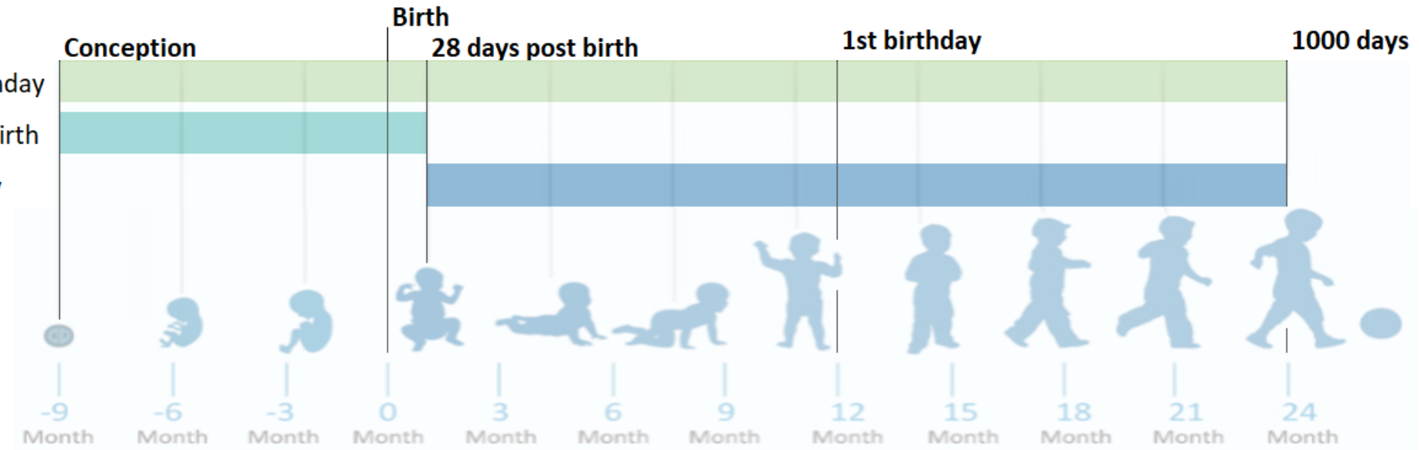

**Supplement 7. Supporting information for Table 1. Categorical maternal, pregnancy and child characteristics at birth of children with and without maternal substance use recorded during the first 1000 days**

| | Total | | No Maternal Substance use recorded in any data | | Maternal substance use recorded in ≥1 data sources | | | | | |
|---|---|---|---|---|---|---|---|---|---|---|
| | | | | | Child protection data[1] | | Health +/or death data | | Any data source | |
| | n | % | n | % | n | % | n | % | n | % |
| Total children | 970,470 | | 937,823 | | 26,045 | | 13,975 | | 32,647 | |
| Child's year of birth | | | | | | | | | | |
| 2008 | 96,288 | 9.9 | 91,731 | 9.8 | 3,936 | 15.1 | 1,616 | 11.6 | 4,557 | 14.0 |
| 2009 | 96,390 | 9.9 | 92,648 | 9.9 | 3,187 | 12.2 | 1,356 | 9.7 | 3,742 | 11.5 |
| 2010 | 96,427 | 9.9 | 93,401 | 10.0 | 2,337 | 9.0 | 1,414 | 10.1 | 3,026 | 9.3 |
| 2011 | 97,187 | 10 | 94,250 | 10.0 | 2,221 | 8.5 | 1,381 | 9.9 | 2,937 | 9.0 |
| 2012 | 99,458 | 10.2 | 96,398 | 10.3 | 2,357 | 9.0 | 1,430 | 10.2 | 3,060 | 9.4 |
| 2013 | 96,899 | 10.0 | 93,707 | 10.0 | 2,549 | 9.8 | 1,371 | 9.8 | 3,192 | 9.8 |
| 2014 | 97,279 | 10.0 | 94,026 | 10.0 | 2,566 | 9.9 | 1,419 | 10.2 | 3,253 | 10 |
| 2015 | 96,345 | 9.9 | 93,220 | 9.9 | 2,448 | 9.4 | 1,407 | 10.1 | 3,125 | 9.6 |
| 2016 | 98,472 | 10.1 | 95,559 | 10.2 | 2,256 | 8.7 | 1,321 | 9.5 | 2,913 | 8.9 |
| 2017 | 95,725 | 9.9 | 92,883 | 9.9 | 2,188 | 8.4 | 1,260 | 9.0 | 2,842 | 8.7 |
| Total, denominator for column | **970,470** | **100** | **937,823** | **100** | **26,045** | **100** | **13,975** | **100** | **32,647** | **100** |
| Child's sex | | | | | | | | | | |
| Male | 498,280 | 51.3 | <482,000 | <52.0 | 13,258 | 50.9 | <7,300 | <52.0 | <16,700 | <52.0 |
| Female | 472,128 | 48.6 | 456,167 | 48.6 | 12,787 | 49.1 | 6,772 | 48.5 | 15,961 | 48.9 |
| Intersex, indeterminate | 46 | <0.1 | <46 | <0.1 | 0 | 0 | <46 | <0.1 | <46 | <0.1 |
| Missing | 16 | <0.1 | 16 | <0.1 | 0 | 0 | 0 | 0 | 0 | 0 |
| Total, denominator for column | **970,470** | **100** | **937,823** | **100** | **26,045** | **100** | **13,975** | **100** | **32,647** | **100** |
| Aboriginal and/or Torres Strait Islander child[2] | | | | | | | | | | |
| No | 903,266 | 93.1 | 882,875 | 94.1 | 15,690 | 60.2 | 9,016 | 64.5 | 20,391 | 62.5 |
| Yes | 67,186 | 6.9 | 54,930 | 5.9 | 10,355 | 39.8 | 4,959 | 35.5 | 12,256 | 37.5 |
| Missing | 18 | <0.1 | 18 | <0.1 | 0 | 0 | 0 | 0 | 0 | 0 |
| Total, denominator for column | **970,470** | **100** | **937,823** | **100** | **26,045** | **100** | **13,975** | **100** | **32,647** | **100** |
| Preterm (<37 weeks) | | | | | | | | | | |
| No | 897,569 | 92.5 | 870,033 | 92.8 | 22,136 | 85 | 11,222 | 80.3 | 27,536 | 84.3 |
| Yes | 72,901 | 7.5 | 67,790 | 7.2 | 3,909 | 15 | 2,753 | 19.7 | 5,111 | 15.7 |
| Total, denominator for column | **970,470** | **100** | **937,823** | **100** | **26,045** | **100** | **13,975** | **100** | **32,647** | **100** |
| Small for gestational age | | | | | | | | | | |
| No | 873148 | 90.0 | 846820 | 90.3 | 21058 | 80.9 | 10866 | 77.8 | 26328 | 80.6 |
| Yes | 96420 | 9.9 | 90125 | 9.6 | 4974 | 19.1 | 3091 | 22.1 | 6295 | 19.3 |
| Missing | 902 | 0.1 | 878 | 0.1 | 13 | 0.0 | 18 | 0.1 | 24 | 0.1 |
| Total, denominator for column | **970470** | **100** | **937823** | **100** | **26045** | **100** | **13975** | **100** | **32647** | **100** |
| Low birth weight;<2500g | | | | | | | | | | |
| No | 908,670 | 93.6 | 881,340 | 94 | 22,008 | 84.5 | 11,012 | 78.8 | 27,330 | 83.7 |
| Yes | 61,030 | 6.3 | 55,734 | 5.9 | 4,027 | 15.5 | 2,947 | 21.1 | 5,296 | 16.2 |
| Missing | 770 | 0.1 | 749 | 0.1 | 10 | <0.1 | 16 | 0.1 | 21 | 0.1 |
| Total, denominator for column | **970,470** | **100** | **937,823** | **100** | **26,045** | **100** | **13,975** | **100** | **32,647** | **100** |
| Admitted to Neonatal Intensive care unit or Special care nursery (Data available for children born 2008-2015)[3] | | | | | | | | | | |
| Not recorded in data | 9 | <0.1 | 9 | <0.1 | 0 | <0.1 | 0 | <0.1 | 0 | <0.1 |
| No admission | 662,543 | 85.3 | 644,038 | 85.9 | 15,016 | 69.5 | 6,557 | 57.5 | 18,505 | 68.8 |
| Admitted to NICU or SCN | 113,167 | 14.6 | 104,803 | 14.0 | 6,566 | 30.4 | 4,826 | 42.4 | 8,364 | 31.1 |
| Missing | 554 | 0.1 | 531 | 0.1 | 19 | 0.1 | 11 | 0.1 | 23 | 0.1 |
| Total, denominator for column (children born 2008-2015) | **776,273** | **100** | **749,381** | **100** | **21,601** | **100** | **11,394** | **100** | **26,892** | **100** |
| Mothers age at delivery (years) | | | | | | | | | | |
| Missing | 13 | <0.1 | 13 | 0 | 0 | 0 | 0 | 0 | 0 | 0 |
| <20 | 27,512 | 2.8 | 22,726 | 2.4 | 4,150 | 15.9 | 1,382 | 9.9 | 4,786 | 14.7 |
| 24 | 120,840 | 12.5 | 112,028 | 11.9 | 7,136 | 27.4 | 3,401 | 24.3 | 8,812 | 27 |

| | Total | | No Maternal Substance use recorded in any data | | Maternal substance use recorded in **≥1 data sources** | | | | | |
|---|---|---|---|---|---|---|---|---|---|---|
| | | | | | Child protection data[1] | | Health +/or death data | | Any data source | |
| | n | % | n | % | n | % | n | % | n | % |
| 29 | 260,485 | 26.8 | 252,548 | 26.9 | 6,255 | 24 | 3,672 | 26.3 | 7,937 | 24.3 |
| 34 | 328,250 | 33.8 | 321,960 | 34.3 | 4,833 | 18.6 | 3,121 | 22.3 | 6,290 | 19.3 |
| 35 + | 233,370 | 24 | 228,548 | 24.4 | 3,671 | 14.1 | 2,399 | 17.2 | 4,822 | 14.8 |
| **Total, denominator for column** | **970,470** | **100** | **937,823** | **100** | **26,045** | **100** | **13,975** | **100** | **32,647** | **100** |
| **Aboriginal or Torres Strait Islander mother** | | | | | | | | | | |
| Not recorded in data | 21,722 | 2.2 | 19,028 | 2 | 1,934 | 7.4 | 1,228 | 8.8 | 2,694 | 8.3 |
| Aboriginal or Torres Strait Islander | 51,003 | 5.3 | 41,914 | 4.5 | 7,796 | 29.9 | 3,674 | 26.3 | 9,089 | 27.8 |
| Non-Aboriginal | 897,616 | 92.5 | 876,764 | 93.5 | 16,304 | 62.6 | 9,066 | 64.9 | 20,852 | 63.9 |
| Missing | 129 | <0.1 | 117 | <0.1 | 11 | <0.1 | 7 | 0.1 | 12 | <0.1 |
| **Total, denominator for column** | **970,470** | **100** | **937,823** | **100** | **26,045** | **100** | **13,975** | **100** | **32,647** | **100** |
| **Mother born in Australia** | | | | | | | | | | |
| No | 334,967 | 34.5 | 331,980 | 35.4 | 2,232 | 8.6 | 1,295 | 9.3 | 2,987 | 9.1 |
| Yes | 634,493 | 65.4 | 604,849 | 64.5 | 23,798 | 91.4 | 12,671 | 90.7 | 29,644 | 90.8 |
| Missing | 1,010 | 0.1 | 994 | 0.1 | 15 | 0.1 | 9 | 0.1 | 16 | 0 |
| **Total, denominator for column** | **970,470** | **100** | **937,823** | **100** | **26,045** | **100** | **13,975** | **100** | **32,647** | **100** |
| **Married or in de facto partnership[4]** | | | | | | | | | | |
| No | 137,720 | 14.2 | 118,820 | 12.7 | 15,560 | 59.7 | 8,087 | 57.9 | 18,900 | 57.9 |
| Yes | 808,753 | 83.3 | 795,798 | 84.9 | 9,845 | 37.8 | 5,538 | 39.6 | 12,955 | 39.7 |
| Missing | 23,997 | 2.5 | 23,205 | 2.5 | 640 | 2.5 | 350 | 2.5 | 792 | 2.4 |
| **Total, denominator for column** | **970,470** | **100** | **937,823** | **100** | **26,045** | **100** | **13,975** | **100** | **32,647** | **100** |
| **Private health insurance/patient[5]** | | | | | | | | | | |
| Not recorded in data | 58 | 0 | 58 | 0 | 0 | 0 | 0 | 0 | 0 | 0 |
| No | 764,546 | 78.8 | 732,503 | 78.1 | 25,745 | 98.8 | 13,632 | 97.5 | 32,043 | 98.1 |
| Yes | 205,866 | 21.2 | 205,262 | 21.9 | 300 | 1.2 | 343 | 2.5 | 604 | 1.9 |
| **Total, denominator for column** | **970,470** | **100** | **937,823** | **100** | **26,045** | **100** | **13,975** | **100** | **32,647** | **100** |
| **Mothers area of residence[6] (missing vales n=<5)** | | | | | | | | | | |
| Not recorded in data | 16,713 | 1.7 | 16,312 | 1.7 | 293 | 1.1 | 227 | 1.6 | 401 | 1.2 |
| Lived in major city | 751,815 | 77.5 | 732,726 | 78.1 | 15,059 | 57.8 | 8,719 | 62.4 | 19,089 | 58.5 |
| Lived in inner regional area | 150,122 | 15.5 | 140,645 | 15.0 | 7,678 | 29.5 | 3,698 | 26.5 | 9,477 | 29.0 |
| Lived in outer regional area | 47,059 | 4.8 | 43,823 | 4.7 | 2,630 | 10.1 | 1,188 | 8.5 | 3,236 | 9.9 |
| Lived in remote/very remote area | 4,759 | 0.4 | 4,317 | 0.4 | 385 | 1.9 | 143 | 1.5 | 444 | 1.8 |
| **Total, denominator for column** | **970,470** | **100** | **937,823** | **100** | **26,045** | **100** | **13,975** | **100** | **32,647** | **100** |
| **Area-level disadvantage[7]** | | | | | | | | | | |
| Not recorded in data | 370 | 0 | 357 | 0 | 7 | 0 | 11 | 0.1 | 13 | 0 |
| Quintile 1 (most disadvantaged) | 213,028 | 22 | 200,803 | 21.4 | 9,893 | 38 | 5,102 | 36.5 | 12,225 | 37.4 |
| Quintile 2 | 177,648 | 18.3 | 169,072 | 18 | 6,982 | 26.8 | 3,417 | 24.5 | 8,576 | 26.3 |
| Quintile 3 | 211,875 | 21.8 | 205,204 | 21.9 | 5,383 | 20.7 | 2,785 | 19.9 | 6,671 | 20.4 |
| Quintile 4 | 148,809 | 15.3 | 145,901 | 15.6 | 2,235 | 8.6 | 1,415 | 10.1 | 2,908 | 8.9 |
| Quintile 5 (least disadvantaged) | 218,740 | 22.5 | 216,486 | 23.1 | 1,545 | 5.9 | 1,245 | 8.9 | 2,254 | 6.9 |
| **Total, denominator for column** | **970,470** | **100** | **937,823** | **100** | **26,045** | **100** | **13,975** | **100** | **32,647** | **100** |
| **Living in or awaiting public housing two years prior to child's birth[8]** | | | | | | | | | | |
| Yes | 925,155 | 95.3 | 903,672 | 96.4 | 16,514 | 63.4 | 9,165 | 65.6 | 21,483 | 65.8 |
| No | 45,315 | 4.7 | 34,151 | 3.6 | 9,531 | 36.6 | 4,810 | 34.4 | 11,164 | 34.2 |
| **Total, denominator for column** | **970,470** | **100** | **937,823** | **100** | **26,045** | **100** | **13,975** | **100** | **32,647** | **100** |
| **Number of previous pregnancies** | | | | | | | | | | |
| Mother had no prior births | 420,220 | 43.3 | 409,737 | 43.7 | 7,975 | 30.6 | 4,511 | 32.3 | 10,483 | 32.1 |
| Mother had one prior birth | 326,902 | 33.7 | 318,947 | 34 | 6,201 | 23.8 | 3,434 | 24.6 | 7,955 | 24.4 |
| Mother had two or more prior births | 222,591 | 22.9 | 208,421 | 22.2 | 11,842 | 45.5 | 6,005 | 43 | 14,170 | 43.4 |
| Missing | 757 | 0.1 | 718 | 0.1 | 27 | 0.1 | 25 | 0.2 | 39 | 0.1 |
| **Total, denominator for column** | **970,470** | **100** | **937,823** | **100** | **26,045** | **100** | **13,975** | **100** | **32,647** | **100** |
| **Received antenatal care in first 20 weeks of pregnancy** | | | | | | | | | | |

| | Total | | No Maternal Substance use recorded in any data | | Maternal substance use recorded in **≥1 data sources** | | | | | |
|---|---|---|---|---|---|---|---|---|---|---|
| | | | | | Child protection data[1] | | Health +/or death data | | Any data source | |
| | n | % | n | % | n | % | n | % | n | % |
| No | 102125 | 10.5 | 94581 | 10.1 | 6256 | 24.0 | 3365 | 24.1 | 7544 | 23.1 |
| Yes | 856670 | 88.3 | 832730 | 88.8 | 18778 | 72.1 | 9993 | 71.5 | 23940 | 73.3 |
| Missing | 11675 | 1.2 | 10512 | 1.1 | 1011 | 3.9 | 617 | 4.4 | 1163 | 3.6 |
| **Total** | **970470** | **100** | **937823** | **100** | **26045** | **100** | **13975** | **100** | **32647** | **100** |
| Mother smoked during pregnancy[9] | | | | | | | | | | |
| No | 870,883 | 89.7 | 858,927 | 91.6 | 9,025 | 34.7 | 4,326 | 31 | 11,956 | 36.6 |
| Yes | 99,260 | 10.2 | 78,612 | 8.4 | 16,985 | 65.2 | 9,630 | 68.9 | 20,648 | 63.2 |
| Missing | 327 | <0.1 | 284 | <0.1 | 35 | 0.1 | 19 | 0.1 | 43 | 0.1 |
| Total, denominator for column | 970,470 | 100 | 937,823 | 100 | 26,045 | 100 | 13,975 | 100 | 32,647 | 100 |

1.The child protection data source includes maternal and other carer substance use; 2. Defined as child or parent identified as Aboriginal on any of the birth records (i.e. perinatal data collection, birth registration or hospital record; 3. Numbers and percents calculated for children born from 2008-2015 (N=776,273) when this variable was collected in the Perinatal data collection; 4. The month and year of birth for the mother was obtained from the birth registration, or the Perinatal Data Collection if the birth registration was unavailable, and used to calculate maternal age at childbirth, in years; 5. As recorded on the hospital birth record; 6. Based on the Accessibility/Remoteness Index of Australia (ARIA+) for the Statistical Local Area of residence at the child’s birth; 7. Based on the Index of Relative Socio-economic Advantage and Disadvantage for the Statistical Local Area of residence at the child’s birth; 9 Mother is living in public housing or is on the tenancy waitlist at any time 2 years prior to the child’s birth. 9. Variable derived from variables recorded in Perinatal Data Collection, indicates if the mother smoked at any time point during pregnancy.

**Supplement 8. Number, percentage, and 95% CIS of children with maternal substance use in the first 1000 days, ascertained from six data sources, for children born in NSW from 2008-2017**

| N = 970, 470 Data source | Single data sources: Cause of death[1] | | Mental health outpatients | | Opioid treatment register | | Emergency department | | Hospital | | Child protection[2] | | Combined data sources: Health +/or cause of death[3] | | Any data source[3] | |
|---|---|---|---|---|---|---|---|---|---|---|---|---|---|---|---|---|
| Substance type | n | % (95% CI) | n | % (95% CI) | n | % (95% CI) | n | % (95% CI) | n | % (95% CI) | n | % (95% CI) | n | % (95% CI) | n | % (95% CI) |
| **Maternal data sources (5 data sources)** | | | | | | | | | | | | | | | | |
| Any substance[4] | <64 | 0.01 (<0.01-0.01) | 964 | 0.10 (0.09-0.11) | 3,229 | 0.33 (0.32-0.34) | 2,203 | 0.23 (0.22-0.24) | 10,793 | 1.11 (1.09-1.13) | - | - | - | - | 12,956 | 1.34 (1.31-1.36) |
| Alcohol | - | - | 511 | <0.01 | - | - | 1,431 | 0.15 (0.14-0.16) | 3,046 | 0.31 (0.3-0.32) | - | - | - | - | 4,246 | 0.44 (0.42-0.45) |
| Other drug[5] | - | - | 473 | <0.01 | 3,229 | 0.33 (0.32-0.34) | 814 | 0.08 (0.08-0.09) | 8,985 | 0.93 (0.91-0.94) | - | - | - | - | 10,126 | 1.04 (1.02-1.06) |
| Opioid/opioid treatment | - | - | 30 | <0.01 | 3,229 | 0.33 (0.32-0.34) | 140 | 0.01 (0.01-0.02) | 3,063 | 0.32 (0.3-0.33) | - | - | - | - | 4,011 | 0.41 (0.4-0.43) |
| Cannabis | - | - | 107 | <0.01 | - | - | 43 | 0 (0-0.01) | 4,435 | 0.46 (0.44-0.47) | - | - | - | - | 4,505 | 0.46 (0.45-0.48) |
| Stimulant | - | - | 102 | <0.01 | - | - | 56 | 0.01 (0-0.01) | 2,117 | 0.22 (0.21-0.23) | - | - | - | - | 2,182 | 0.22 (0.22-0.23) |
| Sedatives or other[6] | - | - | 6 | <0.01 | - | - | 10 | 0 (0-0) | 549 | 0.06 (0.05-0.06) | - | - | - | - | 572 | 0.06 (0.05-0.06) |
| Unspecified or multiple[7] | - | - | 342 | <0.01 | - | - | 618 | 0.06 (0.06-0.07) | 1,858 | 0.19 (0.18-0.2) | - | - | - | - | 2,576 | 0.27 (0.26-0.28) |
| **Child data sources (3 data sources)** | | | | | | | | | | | | | | | | |
| Any substance[4] | <5 | <0.01 | - | - | - | - | - | - | 3,827 | 0.39 (0.38-0.41) | 26,045 | 2.68 (2.65-2.72) | - | - | 27,327 | 2.82 (2.78-2.85) |
| Alcohol | - | - | - | - | - | - | - | - | 62 | 0.01 (<0.01-0.01) | 10,702 | 1.1 (1.08-1.12) | - | - | 10,726 | 1.11 (1.08-1.13) |
| Other drug[5] | - | - | - | - | - | - | - | - | 3,787 | 0.39 (0.38-0.40) | 18,152 | 1.87 (1.84-1.9) | - | - | 19,559 | 2.02 (1.99-2.04) |
| **Child and maternal data sources combined (6 data sources)** | | | | | | | | | | | | | | | | |
| Any substance[4] | <64 | 0.01 (<0.01-0.01) | 964 | 0.10 (0.09-0.11) | 3,229 | 0.33 (0.32-0.34) | 2,203 | 0.23 (0.22-0.24) | 12,067 | 1.24 (1.22-1.27) | 26,045 | 2.68 (2.65-2.72) | 13,975 | 1.44 (1.42-1.46) | 32,647 | 3.36 (3.33-3.40) |
| Alcohol | - | - | 511 | <0.01 | - | - | 1,431 | 0.15 (0.14-0.16) | 3,074 | 0.32 (0.31-0.33) | 10,702 | 1.1 (1.08-1.12) | 11,175 | 1.15 (1.13-1.17) | 13,637 | 1.41 (1.38-1.43) |
| Other drug[5] | - | - | 473 | <0.01 | 3,229 | 0.33 (0.32-0.34) | 814 | 0.08 (0.08-0.09) | 10,282 | 1.06 (1.04-1.08) | 18,152 | 1.87 (1.84-1.9) | 4,272 | 0.44 (0.43-0.45) | 23,485 | 2.42 (2.39-2.45) |

1.For cause of death data, reporting is limited to the outcome "any substance use" due to low numbers in specific substance types; 2. The child protection data source includes maternal and other carer substance use; 3. The outcomes are not mutually exclusive across data sources or substance types; therefore, numbers in rows/columns do not sum to numbers reported in "All records and data sources" (rows) or the "Any substance use"outcome (columns); 4. Any substance use incudes the use of alcohol and or any drug use; 5. Other drug use includes the use of drugs other than alcohol and tobacco, inducing illicit drugs, misuse of prescription drugs, and use of opioid agonist treatment; 6. "Sedatives or other" includes the use of sedatives, hallucinogens, sedatives, solvents, or organic compounds, which were aggregated due to small cell sizes in individual substance categories; 7. "Unspecified or multiple substance use" includes ICD10AM codes that indicate multiple drug use or do not specify the type of drug or psychoactive substance.

**Supplement 9. Data for Figure 1. Number of children with maternal substance use in different combinations of data sources**

| Child protection | Child hospital | Child cause of death | Mother hospital | Mother cause of death | Mother mental health outpatient | Mother opioid treatment register | Mother emergency department | Number of data sources | n | N | % | SE | Lower 95% CI | Upper 95% CI |
|---|---|---|---|---|---|---|---|---|---|---|---|---|---|---|
| No | No | No | No | No | No | No | No | 0 | 937,823 | 970,470 | 0.9664 | 0.0002 | 0.9660 | 0.9667 |
| Yes | No | No | No | No | No | No | No | 1 | 18,672 | 970,470 | 0.0192 | 0.0001 | 0.0190 | 0.0195 |
| No | No | No | Yes | No | No | No | No | 1 | 3,283 | 970,470 | 0.0034 | 0.0001 | 0.0033 | 0.0035 |
| Yes | No | No | Yes | No | No | No | No | 2 | 3,057 | 970,470 | 0.0032 | 0.0001 | 0.0030 | 0.0033 |
| Yes | Yes | No | Yes | No | No | Yes | No | 4 | 1,052 | 970,470 | 0.0011 | 0.0000 | 0.0010 | 0.0011 |
| No | No | No | No | No | No | No | Yes | 1 | 809 | 970,470 | 0.0008 | 0.0000 | 0.0008 | 0.0009 |
| Yes | Yes | No | Yes | No | No | No | No | 3 | 549 | 970,470 | 0.0006 | 0.0000 | 0.0005 | 0.0006 |
| No | Yes | No | No | No | No | No | No | 1 | 544 | 970,470 | 0.0006 | 0.0000 | 0.0005 | 0.0006 |
| Yes | Yes | No | No | No | No | No | No | 2 | 473 | 970,470 | 0.0005 | 0.0000 | 0.0004 | 0.0005 |
| Yes | No | No | Yes | No | No | Yes | No | 3 | 392 | 970,470 | 0.0004 | 0.0000 | 0.0004 | 0.0004 |
| No | Yes | No | Yes | No | No | Yes | No | 3 | 385 | 970,470 | 0.0004 | 0.0000 | 0.0004 | 0.0004 |
| Yes | No | No | Yes | No | No | No | Yes | 3 | 375 | 970,470 | 0.0004 | 0.0000 | 0.0003 | 0.0004 |
| No | No | No | Yes | No | No | Yes | No | 2 | 306 | 970,470 | 0.0003 | 0.0000 | 0.0003 | 0.0004 |
| No | No | No | No | No | No | Yes | No | 1 | 275 | 970,470 | 0.0003 | 0.0000 | 0.0002 | 0.0003 |
| No | No | No | Yes | No | No | No | Yes | 2 | 274 | 970,470 | 0.0003 | 0.0000 | 0.0002 | 0.0003 |
| Yes | No | No | No | No | No | No | Yes | 2 | 230 | 970,470 | 0.0002 | 0.0000 | 0.0002 | 0.0003 |
| Yes | No | No | Yes | No | Yes | No | No | 3 | 221 | 970,470 | 0.0002 | 0.0000 | 0.0002 | 0.0003 |
| No | Yes | No | Yes | No | No | No | No | 2 | 211 | 970,470 | 0.0002 | 0.0000 | 0.0002 | 0.0002 |
| Yes | No | No | No | No | No | Yes | No | 2 | 189 | 970,470 | 0.0002 | 0.0000 | 0.0002 | 0.0002 |
| No | No | No | No | No | Yes | No | No | 1 | 186 | 970,470 | 0.0002 | 0.0000 | 0.0002 | 0.0002 |
| Yes | Yes | No | No | No | No | Yes | No | 3 | 141 | 970,470 | 0.0001 | 0.0000 | 0.0001 | 0.0002 |
| Yes | No | No | No | No | Yes | No | No | 2 | 133 | 970,470 | 0.0001 | 0.0000 | 0.0001 | 0.0002 |
| Yes | Yes | No | Yes | No | No | Yes | Yes | 5 | 116 | 970,470 | 0.0001 | 0.0000 | 0.0001 | 0.0001 |
| Yes | No | No | Yes | No | Yes | No | Yes | 4 | 90 | 970,470 | <0.0001 | <0.0001 | <0.0001 | <0.0001 |
| No | Yes | No | No | No | No | Yes | No | 2 | 87 | 970,470 | <0.0001 | <0.0001 | <0.0001 | <0.0001 |
| No | No | No | Yes | No | Yes | No | No | 2 | 84 | 970,470 | <0.0001 | <0.0001 | <0.0001 | <0.0001 |
| Yes | Yes | No | Yes | No | Yes | Yes | No | 5 | 56 | 970,470 | <0.0001 | <0.0001 | <0.0001 | <0.0001 |
| Yes | Yes | No | Yes | No | No | No | Yes | 4 | 51 | 970,470 | <0.0001 | <0.0001 | <0.0001 | <0.0001 |
| Yes | No | No | Yes | No | No | Yes | Yes | 4 | 50 | 970,470 | <0.0001 | <0.0001 | <0.0001 | <0.0001 |
| Yes | Yes | No | Yes | No | Yes | No | No | 4 | 32 | 970,470 | <0.0001 | <0.0001 | <0.0001 | <0.0001 |
| No | No | No | Yes | No | No | Yes | Yes | 3 | 30 | 970,470 | <0.0001 | <0.0001 | <0.0001 | <0.0001 |
| Yes | No | No | Yes | No | Yes | Yes | No | 4 | 23 | 970,470 | <0.0001 | <0.0001 | <0.0001 | <0.0001 |
| No | No | No | Yes | No | Yes | No | Yes | 3 | 22 | 970,470 | <0.0001 | <0.0001 | <0.0001 | <0.0001 |
| No | Yes | No | Yes | No | No | Yes | Yes | 4 | 21 | 970,470 | <0.0001 | <0.0001 | <0.0001 | <0.0001 |

| Child protection | Child hospital | Child cause of death | Mother hospital | Mother cause of death | Mother mental health outpatient | Mother opioid treatment register | Mother emergency department | Number of data sources | n | N | % | SE | Lower 95% CI | Upper 95% CI |
|---|---|---|---|---|---|---|---|---|---|---|---|---|---|---|
| Yes | Yes | No | Yes | No | Yes | No | Yes | 5 | 19 | 970,470 | <0.0001 | <0.0001 | <0.0001 | <0.0001 |
| Yes | Yes | No | Yes | No | Yes | Yes | Yes | 6 | 19 | 970,470 | <0.0001 | <0.0001 | <0.0001 | <0.0001 |
| Yes | No | No | No | Yes | No | No | No | 2 | 17 | 970,470 | <0.0001 | <0.0001 | <0.0001 | <0.0001 |
| No | No | No | No | Yes | No | No | No | 1 | 15 | 970,470 | <0.0001 | <0.0001 | <0.0001 | <0.0001 |
| Yes | No | No | No | No | Yes | No | Yes | 3 | 15 | 970,470 | <0.0001 | <0.0001 | <0.0001 | <0.0001 |
| Yes | No | No | No | No | No | Yes | Yes | 3 | 14 | 970,470 | <0.0001 | <0.0001 | <0.0001 | <0.0001 |
| No | No | No | No | No | No | Yes | Yes | 2 | 11 | 970,470 | <0.0001 | <0.0001 | <0.0001 | <0.0001 |
| No | No | No | No | No | Yes | No | Yes | 2 | 10 | 970,470 | <0.0001 | <0.0001 | <0.0001 | <0.0001 |
| Yes | No | No | Yes | No | Yes | Yes | Yes | 5 | 10 | 970,470 | <0.0001 | <0.0001 | <0.0001 | <0.0001 |
| No | Yes | No | Yes | No | No | No | Yes | 3 | 9 | 970,470 | <0.0001 | <0.0001 | <0.0001 | <0.0001 |
| Yes | Yes | No | Yes | Yes | No | Yes | No | 5 | 9 | 970,470 | <0.0001 | <0.0001 | <0.0001 | <0.0001 |
| No | No | No | Yes | No | Yes | Yes | No | 3 | 8 | 970,470 | <0.0001 | <0.0001 | <0.0001 | <0.0001 |
| No | Yes | No | Yes | No | Yes | No | No | 3 | 7 | 970,470 | <0.0001 | <0.0001 | <0.0001 | <0.0001 |
| No | Yes | No | Yes | No | Yes | Yes | Yes | 5 | 6 | 970,470 | <0.0001 | <0.0001 | <0.0001 | <0.0001 |
| Yes | Yes | No | No | No | No | No | Yes | 3 | 6 | 970,470 | <0.0001 | <0.0001 | <0.0001 | <0.0001 |
| Yes | Yes | No | No | No | Yes | No | No | 3 | 6 | 970,470 | <0.0001 | <0.0001 | <0.0001 | <0.0001 |
| Yes | No | No | Yes | Yes | No | No | No | 3 | 5 | 970,470 | <0.0001 | <0.0001 | <0.0001 | <0.0001 |
| Yes | Yes | No | No | No | No | Yes | Yes | 4 | 5 | 970,470 | <0.0001 | <0.0001 | <0.0001 | <0.0001 |
| Yes | No | No | Yes | Yes | No | Yes | No | 4 | <5 | 970,470 | <0.0001 | <0.0001 | <0.0001 | <0.0001 |
| No | No | No | Yes | No | Yes | Yes | Yes | 4 | <5 | 970,470 | <0.0001 | <0.0001 | <0.0001 | <0.0001 |
| No | Yes | No | No | No | No | No | Yes | 2 | <5 | 970,470 | <0.0001 | <0.0001 | <0.0001 | <0.0001 |
| No | Yes | No | No | No | Yes | No | No | 2 | <5 | 970,470 | <0.0001 | <0.0001 | <0.0001 | <0.0001 |
| No | Yes | No | Yes | No | Yes | Yes | No | 4 | <5 | 970,470 | <0.0001 | <0.0001 | <0.0001 | <0.0001 |
| No | No | No | Yes | Yes | No | Yes | No | 3 | <5 | 970,470 | <0.0001 | <0.0001 | <0.0001 | <0.0001 |
| Yes | Yes | No | No | No | Yes | Yes | No | 4 | <5 | 970,470 | <0.0001 | <0.0001 | <0.0001 | <0.0001 |
| Yes | Yes | No | Yes | Yes | No | Yes | Yes | 6 | <5 | 970,470 | <0.0001 | <0.0001 | <0.0001 | <0.0001 |
| Yes | Yes | Yes | Yes | No | No | Yes | No | 5 | <5 | 970,470 | <0.0001 | <0.0001 | <0.0001 | <0.0001 |
| No | No | No | No | No | Yes | Yes | No | 2 | <5 | 970,470 | <0.0001 | <0.0001 | <0.0001 | <0.0001 |
| No | No | No | No | Yes | No | Yes | No | 2 | <5 | 970,470 | <0.0001 | <0.0001 | <0.0001 | <0.0001 |
| No | Yes | No | No | No | Yes | Yes | No | 3 | <5 | 970,470 | <0.0001 | <0.0001 | <0.0001 | <0.0001 |
| No | Yes | No | Yes | No | Yes | No | Yes | 4 | <5 | 970,470 | <0.0001 | <0.0001 | <0.0001 | <0.0001 |
| No | Yes | No | Yes | Yes | No | Yes | No | 4 | <5 | 970,470 | <0.0001 | <0.0001 | <0.0001 | <0.0001 |
| Yes | No | No | No | No | Yes | Yes | No | 3 | <5 | 970,470 | <0.0001 | <0.0001 | <0.0001 | <0.0001 |
| Yes | No | No | Yes | Yes | Yes | No | No | 4 | <5 | 970,470 | <0.0001 | <0.0001 | <0.0001 | <0.0001 |
| Yes | No | Yes | No | No | No | No | No | 2 | <5 | 970,470 | <0.0001 | <0.0001 | <0.0001 | <0.0001 |
| Yes | Yes | No | No | No | Yes | Yes | Yes | 5 | <5 | 970,470 | <0.0001 | <0.0001 | <0.0001 | <0.0001 |

| Child protection | Child hospital | Child cause of death | Mother hospital | Mother cause of death | Mother mental health outpatient | Mother opioid treatment register | Mother emergency department | Number of data sources | n | N | % | SE | Lower 95% CI | Upper 95% CI |
|---|---|---|---|---|---|---|---|---|---|---|---|---|---|---|
| Yes | Yes | No | No | Yes | No | No | No | 3 | <5 | 970,470 | <0.0001 | <0.0001 | <0.0001 | <0.0001 |
| Yes | Yes | No | Yes | Yes | No | No | No | 4 | <5 | 970,470 | <0.0001 | <0.0001 | <0.0001 | <0.0001 |
| Yes | Yes | No | Yes | Yes | No | No | Yes | 5 | <5 | 970,470 | <0.0001 | <0.0001 | <0.0001 | <0.0001 |
| Yes | Yes | Yes | No | No | No | No | No | 3 | <5 | 970,470 | <0.0001 | <0.0001 | <0.0001 | <0.0001 |

Footnotes: Total number of children with values <5 is 37; 1. n = numerator, number of children with maternal substance use recorded in the combination of data sources; N = denominator, total children born from 2008-2017; [illegible] error; 95% CI: 95% confidence interval

**Supplement 10. Most common combinations of substance types recorded in maternal data sources, among children born in NSW from 2008-2017 (n=970,470)**

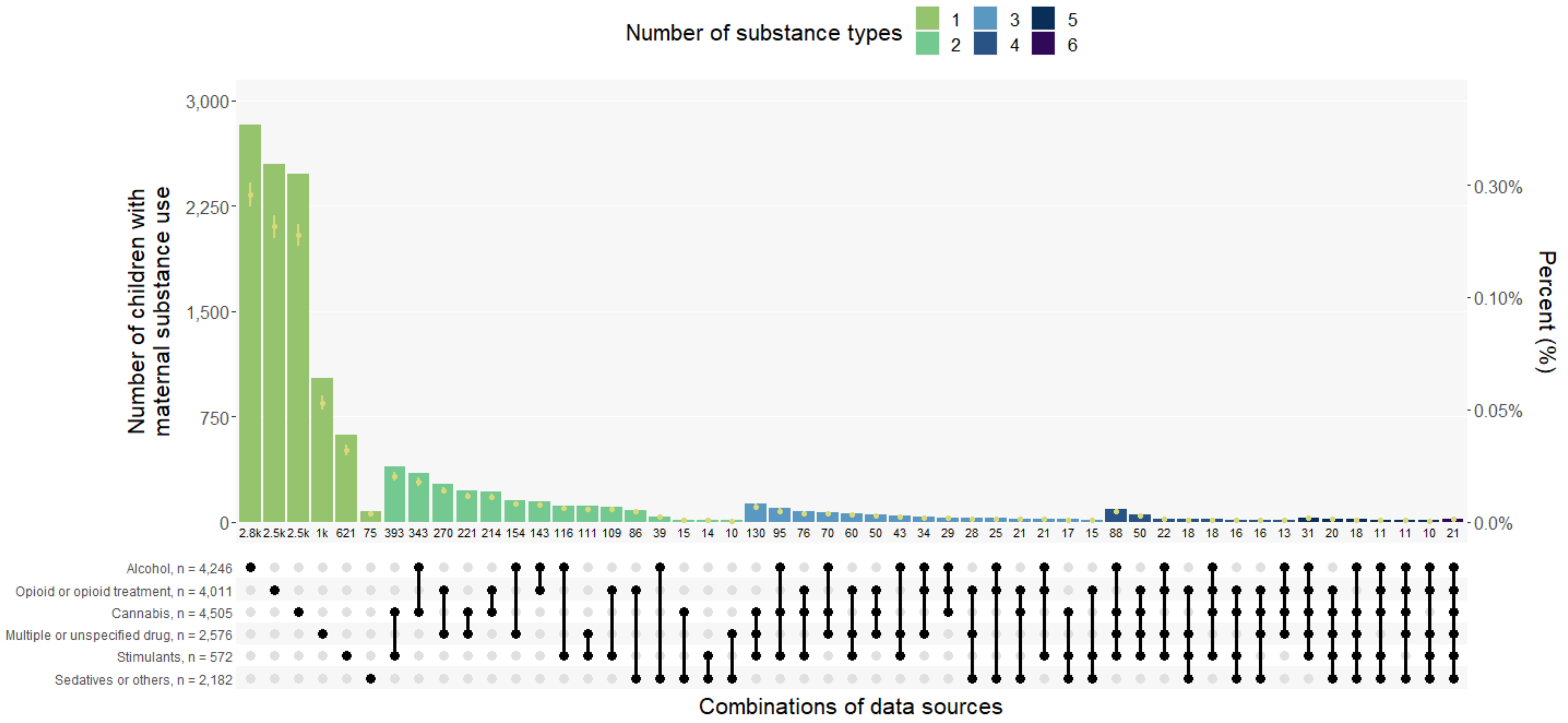

Footnotes: 86 children were not included in upset plot as we excluded combinations of substances with less than 10 children. All values for numerators, denominators, and 95% confidence intervals provided in supplement 10. Definition: “Sedatives or other” includes the use of sedatives, hallucinogens, sedatives, solvents, or organic compounds, which were aggregated due to small cell sizes in individual substance categories; “Unspecified or multiple substance use” includes ICD10AM codes that indicate multiple drug use or do not specify the type of drug or psychoactive substance.

**Supplement 11. Data to accompany Supplement 9. Number of children with maternal substance use in different combinations of substance types recorded in maternal records**

| Opioid | Alcohol | Cannabis | Sedatives or other[1] | Unspecified or multiple[2] | Stimulant | Number of substance types | N[3] | N | % | SE | Lower 95% CI | Upper 95% CI |
|---|---|---|---|---|---|---|---|---|---|---|---|---|
| No | No | No | No | No | No | 0 | 957495 | 970470 | 0.9866 | 0.0001 | 0.9864 | 0.9869 |
| No | Yes | No | No | No | No | 1 | 2825 | 970470 | 0.0029 | 0.0001 | 0.0028 | 0.0030 |
| Yes | No | No | No | No | No | 1 | 2550 | 970470 | 0.0026 | 0.0001 | 0.0025 | 0.0027 |
| No | No | Yes | No | No | No | 1 | 2477 | 970470 | 0.0026 | 0.0001 | 0.0025 | 0.0027 |
| No | No | No | No | Yes | No | 1 | 1026 | 970470 | 0.0011 | 0.0000 | 0.0010 | 0.0011 |
| No | No | No | No | No | Yes | 1 | 621 | 970470 | 0.0006 | 0.0000 | 0.0006 | 0.0007 |
| No | No | Yes | No | No | Yes | 2 | 393 | 970470 | 0.0004 | 0.0000 | 0.0004 | 0.0004 |
| No | Yes | Yes | No | No | No | 2 | 343 | 970470 | 0.0004 | 0.0000 | 0.0003 | 0.0004 |
| Yes | No | No | No | Yes | No | 2 | 270 | 970470 | 0.0003 | 0.0000 | 0.0002 | 0.0003 |
| No | No | Yes | No | Yes | No | 2 | 221 | 970470 | 0.0002 | 0.0000 | 0.0002 | 0.0003 |
| Yes | No | Yes | No | No | No | 2 | 214 | 970470 | 0.0002 | 0.0000 | 0.0002 | 0.0003 |
| No | Yes | No | No | Yes | No | 2 | 154 | 970470 | 0.0002 | 0.0000 | 0.0001 | 0.0002 |
| Yes | Yes | No | No | No | No | 2 | 143 | 970470 | 0.0001 | 0.0000 | 0.0001 | 0.0002 |
| No | No | Yes | No | Yes | Yes | 3 | 130 | 970470 | 0.0001 | 0.0000 | 0.0001 | 0.0002 |
| No | Yes | No | No | No | Yes | 2 | 116 | 970470 | 0.0001 | 0.0000 | 0.0001 | 0.0001 |
| No | No | No | No | Yes | Yes | 2 | 111 | 970470 | 0.0001 | 0.0000 | 0.0001 | 0.0001 |
| Yes | No | No | No | No | Yes | 2 | 109 | 970470 | 0.0001 | 0.0000 | 0.0001 | 0.0001 |
| No | Yes | Yes | No | No | Yes | 3 | 95 | 970470 | <0.0001 | <0.0001 | <0.0001 | <0.0001 |
| No | Yes | Yes | No | Yes | Yes | 4 | 88 | 970470 | <0.0001 | <0.0001 | <0.0001 | <0.0001 |
| Yes | No | No | Yes | No | No | 2 | 86 | 970470 | <0.0001 | <0.0001 | <0.0001 | <0.0001 |
| Yes | No | Yes | No | No | Yes | 3 | 76 | 970470 | <0.0001 | <0.0001 | <0.0001 | <0.0001 |
| No | No | No | Yes | No | No | 1 | 75 | 970470 | <0.0001 | <0.0001 | <0.0001 | <0.0001 |
| No | Yes | Yes | No | Yes | No | 3 | 70 | 970470 | <0.0001 | <0.0001 | <0.0001 | <0.0001 |
| Yes | No | No | No | Yes | Yes | 3 | 60 | 970470 | <0.0001 | <0.0001 | <0.0001 | <0.0001 |
| Yes | No | Yes | No | Yes | No | 3 | 50 | 970470 | <0.0001 | <0.0001 | <0.0001 | <0.0001 |
| Yes | No | Yes | No | Yes | Yes | 4 | 50 | 970470 | <0.0001 | <0.0001 | <0.0001 | <0.0001 |
| No | Yes | No | No | Yes | Yes | 3 | 43 | 970470 | <0.0001 | <0.0001 | <0.0001 | <0.0001 |
| No | Yes | No | Yes | No | No | 2 | 39 | 970470 | <0.0001 | <0.0001 | <0.0001 | <0.0001 |
| Yes | Yes | No | No | Yes | No | 3 | 34 | 970470 | <0.0001 | <0.0001 | <0.0001 | <0.0001 |
| Yes | Yes | Yes | No | Yes | Yes | 5 | 31 | 970470 | <0.0001 | <0.0001 | <0.0001 | <0.0001 |
| Yes | Yes | Yes | No | No | No | 3 | 29 | 970470 | <0.0001 | <0.0001 | <0.0001 | <0.0001 |
| Yes | No | No | Yes | Yes | No | 3 | 28 | 970470 | <0.0001 | <0.0001 | <0.0001 | <0.0001 |
| Yes | Yes | No | Yes | No | No | 3 | 25 | 970470 | <0.0001 | <0.0001 | <0.0001 | <0.0001 |

| Opioid | Alcohol | Cannabis | Sedatives or other[1] | Unspecified or multiple[2] | Stimulant | Number of substance types | N[3] | N | % | SE | Lower 95% CI | Upper 95% CI |
|---|---|---|---|---|---|---|---|---|---|---|---|---|
| Yes | Yes | No | No | Yes | Yes | 4 | 22 | 970470 | <0.0001 | <0.0001 | <0.0001 | <0.0001 |
| Yes | No | Yes | Yes | No | No | 3 | 21 | 970470 | <0.0001 | <0.0001 | <0.0001 | <0.0001 |
| Yes | Yes | No | No | No | Yes | 3 | 21 | 970470 | <0.0001 | <0.0001 | <0.0001 | <0.0001 |
| Yes | Yes | Yes | Yes | Yes | Yes | 6 | 21 | 970470 | <0.0001 | <0.0001 | <0.0001 | <0.0001 |
| Yes | No | Yes | Yes | Yes | Yes | 5 | 20 | 970470 | <0.0001 | <0.0001 | <0.0001 | <0.0001 |
| No | Yes | Yes | Yes | Yes | Yes | 5 | 18 | 970470 | <0.0001 | <0.0001 | <0.0001 | <0.0001 |
| Yes | No | No | Yes | Yes | Yes | 4 | 18 | 970470 | <0.0001 | <0.0001 | <0.0001 | <0.0001 |
| Yes | Yes | Yes | No | No | Yes | 4 | 18 | 970470 | <0.0001 | <0.0001 | <0.0001 | <0.0001 |
| No | No | Yes | Yes | No | Yes | 3 | 17 | 970470 | <0.0001 | <0.0001 | <0.0001 | <0.0001 |
| Yes | No | Yes | Yes | No | Yes | 4 | 16 | 970470 | <0.0001 | <0.0001 | <0.0001 | <0.0001 |
| Yes | No | Yes | Yes | Yes | No | 4 | 16 | 970470 | <0.0001 | <0.0001 | <0.0001 | <0.0001 |
| No | No | Yes | Yes | No | No | 2 | 15 | 970470 | <0.0001 | <0.0001 | <0.0001 | <0.0001 |
| Yes | No | No | Yes | No | Yes | 3 | 15 | 970470 | <0.0001 | <0.0001 | <0.0001 | <0.0001 |
| No | No | No | Yes | No | Yes | 2 | 14 | 970470 | <0.0001 | <0.0001 | <0.0001 | <0.0001 |
| Yes | Yes | Yes | No | Yes | No | 4 | 13 | 970470 | <0.0001 | <0.0001 | <0.0001 | <0.0001 |
| Yes | Yes | Yes | Yes | No | Yes | 5 | 11 | 970470 | <0.0001 | <0.0001 | <0.0001 | <0.0001 |
| Yes | Yes | Yes | Yes | Yes | No | 5 | 11 | 970470 | <0.0001 | <0.0001 | <0.0001 | <0.0001 |
| No | No | No | Yes | Yes | No | 2 | 10 | 970470 | <0.0001 | <0.0001 | <0.0001 | <0.0001 |
| Yes | Yes | No | Yes | Yes | Yes | 5 | 10 | 970470 | <0.0001 | <0.0001 | <0.0001 | <0.0001 |
| No | No | No | Yes | Yes | Yes | 3 | 9 | 970470 | <0.0001 | <0.0001 | <0.0001 | <0.0001 |
| No | Yes | Yes | Yes | No | No | 3 | 9 | 970470 | <0.0001 | <0.0001 | <0.0001 | <0.0001 |
| No | Yes | Yes | Yes | Yes | No | 4 | 9 | 970470 | <0.0001 | <0.0001 | <0.0001 | <0.0001 |
| Yes | Yes | No | Yes | Yes | No | 4 | 9 | 970470 | <0.0001 | <0.0001 | <0.0001 | <0.0001 |
| No | Yes | No | Yes | No | Yes | 3 | 8 | 970470 | <0.0001 | <0.0001 | <0.0001 | <0.0001 |
| Yes | Yes | Yes | Yes | No | No | 4 | 8 | 970470 | <0.0001 | <0.0001 | <0.0001 | <0.0001 |
| No | Yes | No | Yes | Yes | No | 3 | 7 | 970470 | <0.0001 | <0.0001 | <0.0001 | <0.0001 |
| No | No | Yes | Yes | Yes | No | 3 | 6 | 970470 | <0.0001 | <0.0001 | <0.0001 | <0.0001 |
| No | Yes | No | Yes | Yes | Yes | 4 | 6 | 970470 | <0.0001 | <0.0001 | <0.0001 | <0.0001 |
| Yes | Yes | No | Yes | No | Yes | 4 | 6 | 970470 | <0.0001 | <0.0001 | <0.0001 | <0.0001 |
| No | No | Yes | Yes | Yes | Yes | 4 | ≤5 | 970470 | <0.0001 | <0.0001 | <0.0001 | <0.0001 |
| No | Yes | Yes | Yes | No | Yes | 4 | ≤5 | 970470 | <0.0001 | <0.0001 | <0.0001 | <0.0001 |

Footnotes: 1. "Sedatives or other" includes the use of sedatives, hallucinogens, sedatives, solvents, or organic compounds, which were aggregated due to small cell sizes in individual substance categories;2. "Unspecified or multiple substance use" includes ICD10AM codes that indicate multiple drug use or do not specify the type of drug or psychoactive substance. 3. Total number of children with values ≤5 is 9. Definitions: n = numerator, number of children with maternal substance use recorded in the combination of data sources; N = denominator, total children born from 2008-2017; SE: Standard error; 95% CI: 95% confidence interval.

**Supplement 12. Data to accompany Figure 2. Child with maternal substance use in the first 1000 days of life, ascertained from different combinations of data sources, by child's year of birth**

| Year of birth | N[1] | Data source/s | N[2] | % | 95% CI |
|---|---|---|---|---|---|
| 2008 | 96,288 | Child protection records[2] | 3,936 | 4.09 | 3.96 - 4.21 |
| 2009 | 96,390 | Child protection records[2] | 3,187 | 3.31 | 3.19 - 3.42 |
| 2010 | 96,427 | Child protection records[2] | 2,337 | 2.42 | 2.33 - 2.52 |
| 2011 | 97,187 | Child protection records[2] | 2,221 | 2.29 | 2.19 - 2.38 |
| 2012 | 99,458 | Child protection records[2] | 2,357 | 2.37 | 2.28 - 2.46 |
| 2013 | 96,899 | Child protection records[2] | 2,549 | 2.63 | 2.53 - 2.73 |
| 2014 | 97,279 | Child protection records[2] | 2,566 | 2.64 | 2.54 - 2.74 |
| 2015 | 96,345 | Child protection records[2] | 2,448 | 2.54 | 2.44 - 2.64 |
| 2016 | 98,472 | Child protection records[2] | 2,256 | 2.29 | 2.20 - 2.38 |
| 2017 | 95,725 | Child protection records[2] | 2,188 | 2.29 | 2.19 - 2.38 |
| 2008 | 96,288 | Five health and death records[3] | 1,616 | 1.68 | 1.60 - 1.76 |
| 2009 | 96,390 | Five health and death records | 1,356 | 1.41 | 1.33 - 1.48 |
| 2010 | 96,427 | Five health and death records | 1,414 | 1.47 | 1.39 - 1.54 |
| 2011 | 97,187 | Five health and death records | 1,381 | 1.42 | 1.35 - 1.5 |
| 2012 | 99,458 | Five health and death records | 1,430 | 1.44 | 1.36 - 1.51 |
| 2013 | 96,899 | Five health and death records | 1,371 | 1.41 | 1.34 - 1.49 |
| 2014 | 97,279 | Five health and death records | 1,419 | 1.46 | 1.38 - 1.53 |
| 2015 | 96,345 | Five health and death records | 1,407 | 1.46 | 1.38 - 1.54 |
| 2016 | 98,472 | Five health and death records | 1,321 | 1.34 | 1.27 - 1.41 |
| 2017 | 95,725 | Five health and death records | 1,260 | 1.32 | 1.24 - 1.39 |
| 2008 | 96,288 | All six mother and child records[4] | 4,557 | 4.73 | 4.60 - 4.87 |
| 2009 | 96,390 | All six mother and child records | 3,742 | 3.88 | 3.76 – 4.00 |
| 2010 | 96,427 | All six mother and child records | 3,026 | 3.14 | 3.03 - 3.25 |
| 2011 | 97,187 | All six mother and child records | 2,937 | 3.02 | 2.91 - 3.13 |
| 2012 | 99,458 | All six mother and child records | 3,060 | 3.08 | 2.97 - 3.18 |
| 2013 | 96,899 | All six mother and child records | 3,192 | 3.29 | 3.18 - 3.41 |
| 2014 | 97,279 | All six mother and child records | 3,253 | 3.34 | 3.23 - 3.46 |
| 2015 | 96,345 | All six mother and child records | 3,125 | 3.24 | 3.13 - 3.36 |
| 2016 | 98,472 | All six mother and child records | 2,913 | 2.96 | 2.85 - 3.06 |
| 2017 | 95,725 | All six mother and child records | 2,842 | 2.97 | 2.86 - 3.08 |

Footnotes: 1. Denominator value, the total number of children born each year; 2. Numerator values, the number of children with maternal substance use recorded in data source/s born each year; 3. The child protection data source includes maternal and other carer substance use 4. Substance use ascertained from maternal hospital, cause of death, mental health outpatients, opioid treatment registry and, emergency department, and child hospital and cause of death records; 5. Substance use ascertained from maternal hospital, cause of death, mental health outpatients, opioid treatment registry, emergency department and child protection records, and from child hospital, cause of death and child protection records. Definitions: CI: Confidence interval.

**Supplement 13. Data to accompany Figure 3. Number and percentage of children born in NSW from 2008-2017 (N=970,470) with maternal substance use records during three ascertainment periods and six administrative data sources for children and mothers**

| Time window | Prenatal (Conception to <28 days post birth) | | | Early life (28 days post-birth to 2nd birthday) | | | First 1000 days[2] (Conception to 2nd birthday) | | |
|---|---|---|---|---|---|---|---|---|---|
| Substance type | n[1] | % | 95% CIs | n | % | 95% CIs | n | % | 95% CIs |
| **Any substance use** | | | | | | | | | |
| All six mother and child records[3] | 13,987 | 1.44 | 1.42 - 1.46 | 26,198 | 2.70 | 2.67 - 2.73 | 32,647 | 3.36 | 3.33 - 3.40 |
| Child protection records | 10,434 | 1.08 | 1.05 - 1.1 | 20,556 | 2.12 | 2.09 - 2.15 | 26,045 | 2.68 | 2.65 - 2.72 |
| Five health and death records[4] | 7,089 | 0.73 | 0.71 - 0.75 | 9,396 | 0.97 | 0.95 - 0.99 | 13,975 | 1.44 | 1.42 - 1.46 |
| **Alcohol use** | | | | | | | | | |
| All six mother and child records[3] | 3,776 | 0.39 | 0.38 - 0.4 | 11,262 | 1.16 | 1.14 - 1.18 | 13,637 | 1.41 | 1.38 - 1.43 |
| Child protection records | 3,046 | 0.31 | 0.3 - 0.32 | 8,659 | 0.89 | 0.87 - 0.91 | 10,702 | 1.1 | 1.08 - 1.12 |
| Five health and death records[4] | 1,072 | 0.11 | 0.1 - 0.12 | 3,490 | 0.36 | 0.35 - 0.37 | 4,272 | 0.44 | 0.43 - 0.45 |
| **Other drug use** | | | | | | | | | |
| All six mother and child records[3] | 11,605 | 1.2 | 1.17 - 1.22 | 17,479 | 1.8 | 1.77 - 1.83 | 23,485 | 2.42 | 2.39 - 2.45 |
| Child protection records | 8,311 | 0.86 | 0.84 - 0.87 | 13,109 | 1.35 | 1.33 - 1.37 | 18,152 | 1.87 | 1.84 - 1.9 |
| Five health and death records[4] | 6,376 | 0.66 | 0.64 - 0.67 | 6,946 | 0.72 | 0.7 - 0.73 | 11,175 | 1.15 | 1.13 - 1.17 |

Definitions: CI: Confidence interval. 1. Numerator value/number of children with substance use recorded in data source/s. 2. The outcomes are not mutually exclusive across each time period; therefore, the number of children in the prenatal period and early life periods do not sum to the total number in the Frist 1000 days. 3. Substance use ascertained from maternal hospital, cause of death, mental health outpatients, opioid treatment registry, emergency department and child protection records; 4. Substance use ascertained from maternal hospital, cause of death, mental health outpatients, opioid treatment registry, and emergency department and child protection records.

**Supplement 14. Sensitivity analysis: Number and percentage of children with maternal substance use in the first 1000 days, ascertained from six data sources, for children born in NSW from 2008-2017 (N=970,470) using additional ICD codes for substance use ascertainment from maternal hospital, mental health and cause of death records**

| Data source | Single data sources | | | | | | | | | | | | Combined data sources | | | |
|---|---|---|---|---|---|---|---|---|---|---|---|---|---|---|---|---|
| | Cause of death[1] | | Mental health outpatients | | Opioid treatment register | | Emergency department | | Hospital | | Child protection[2] | | Health +/or cause of death[3] | | Any data source[3] | |
| Substance type | n | %[3] | n | %[3] | n | %[3] | n | %[3] | n | %[3] | n | %[3] | n | %[3] | n | %[3] |
| **Maternal data sources** (5 data sources) | | | | | | | | | | | | | | | | |
| Any substance[4] | <73 | <0.1 | 1,042 | 0.1 | 3,229 | 0.3 | 2,264 | 0.2 | 10,998 | 1.1 | - | - | - | - | 13,201 | 1.4 |
| Alcohol | - | - | 511 | 0.1 | - | - | 1,431 | 0.2 | 3,046 | 0.3 | - | - | - | - | 4,246 | 0.4 |
| Other drug[5] | - | - | 562 | <0.1 | 3,229 | 0.3 | 877 | 0.1 | 9,268 | 1.0 | - | - | - | - | 10,467 | 1.1 |
| Opioid or opioid treatment | - | - | 30 | <0.1 | 3,229 | 0.3 | 144 | <0.1 | 3,064 | 0.3 | - | - | - | - | 4,013 | 0.4 |
| Cannabis | - | - | 107 | <0.1 | - | - | 43 | <0.1 | 4,435 | 0.5 | - | - | - | - | 4,505 | 0.5 |
| Stimulant | - | - | 104 | <0.1 | - | - | 62 | <0.1 | 2,118 | 0.2 | - | - | - | - | 2,191 | 0.2 |
| Sedatives or other[6] | - | - | 23 | <0.1 | - | - | 66 | <0.1 | 1,020 | 0.1 | - | - | - | - | 1,088 | 0.1 |
| Unspecified or multiple[7] | - | - | 342 | <0.1 | - | - | 618 | <0.1 | 1,860 | 0.2 | - | - | - | - | 2,578 | 0.3 |
| **Child data sources** (3 data sources) | | | | | | | | | | | | | | | | |
| Any substance[4] | <5 | <0.1 | - | - | - | - | - | - | 3,827 | 0.4 | 26,045 | 2.7 | - | - | **27,327** | **2.8** |
| Alcohol | - | - | - | - | - | - | - | - | 62 | <0.1 | 10,702 | 1.1 | - | - | **10,726** | **1.1** |
| Other drug[5] | - | - | - | - | - | - | - | - | 3,787 | 0.4 | 18,152 | 1.9 | - | - | **19,559** | **2.0** |
| **Child and maternal data sources combined** (6 data sources)[2] | | | | | | | | | | | | | | | | |
| Any substance[4] | 73 | <0.1 | 1,042 | 0.1 | 3,229 | 0.3 | 2,264 | 0.2 | 12,272 | 1.3 | 26,045 | 2.7 | **14,219** | **1.5** | **32,840** | **3.4** |
| Alcohol | - | - | 511 | 0.1 | - | - | 1,431 | 0.2 | 3,074 | 0.3 | 10,702 | 1.1 | **11,514** | **1.2** | **13,637** | **1.4** |
| Other drug[5] | - | - | 562 | 0.1 | 3,229 | 0.3 | 877 | 0.1 | 10,564 | 1.1 | 18,152 | 1.9 | **4,272** | **0.4** | **23,768** | **2.4** |

1.For cause of death data, reporting is limited to the outcome "any substance use" due to low numbers in specific substance types; 2. The child protection data source includes maternal and other carer substance use; 3. The outcomes are not mutually exclusive across data sources or substance types; therefore, numbers in rows/columns do not sum to numbers reported in "All records and data sources" (rows) or the "Any substance use"outcome (columns); 3. 95% confidence intervals provided in supplement 7; 4. Any substance use incudes the use of alcohol and or any drug use; 5. Other drug use includes the use of drugs other than alcohol and tobacco, inducing illicit drugs, misuse of prescription drugs, and use of opioid agonist treatment; 6. "Sedatives or other" includes the use of sedatives, hallucinogens, sedatives, solvents, or organic compounds, which were aggregated due to small cell sizes in individual substance categories; 7. "Unspecified or multiple substance use" includes ICD10AM codes that indicate multiple drug use or do not specify the type of drug or psychoactive substance.

**Supplement 15. Sensitivity analysis: number and percentage of children with maternal substance use in the first 1000 days, ascertained from five data sources, for children born in NSW from 2008-2017 (N=970,470)**

| Data source | Single data sources | | | | | | | | | | | | Combined data sources | | | |
|---|---|---|---|---|---|---|---|---|---|---|---|---|---|---|---|---|
| | Cause of death[1] | | Mental health outpatients | | Opioid treatment register | | Emergency department | | Hospital | | Child protection[2] | | Health +/or cause of death[3] | | Any data source[3] | |
| Substance type | n | % | n | % | n | % | n | % | n | % | n | % | n | % | n | % |
| **Maternal data sources** (5 data sources) | | | | | | | | | | | | | | | | |
| Any substance[4] | <64 | <0.1 | 964 | 0.1 | 3,229 | 0.3 | 2,206 | 0.2 | 10,793 | 1.1 | - | - | - | - | 12,956 | 1.3 |
| Alcohol | - | - | 511 | <0.1 | - | - | 1,431 | 0.1 | 3,046 | 0.3 | - | - | - | - | 4,246 | 0.4 |
| Other drug[5] | - | - | 473 | <0.1 | 3,229 | 0.3 | 817 | 0.1 | 8,985 | 0.9 | - | - | - | - | 10,126 | 1.0 |
| Cannabis | - | - | 107 | <0.1 | - | - | 43 | <0.1 | 4,435 | 0.5 | - | - | - | - | 4,505 | 0.5 |
| Opioid or opioid treatment | - | - | 30 | <0.1 | 3,229 | 0.3 | 144 | <0.1 | 3,063 | 0.3 | - | - | - | - | 4,011 | 0.4 |
| Stimulant | - | - | 102 | <0.1 | - | - | 56 | <0.1 | 2,117 | 0.2 | - | - | - | - | 2,182 | 0.2 |
| Sedatives or other[6] | - | - | 6 | <0.1 | - | - | 10 | <0.1 | 549 | 0.1 | - | - | - | - | 572 | 0.1 |
| Unspecified or multiple[7] | - | - | 342 | <0.1 | - | - | 618 | 0.1 | 1,858 | 0.2 | - | - | - | - | 2,576 | 0.3 |
| **Child data sources** (3 data sources) | | | | | | | | | | | | | | | | |
| Any substance[4] | <5 | <0.1 | - | - | - | - | - | - | 3,827 | 0.4 | 26,045 | 2.7 | - | - | 27,327 | 2.8 |
| Alcohol | - | - | - | - | - | - | - | - | 62 | <0.1 | 10,702 | 1.1 | - | - | 10,726 | 1.1 |
| Other drug[5] | - | - | - | - | - | - | - | - | 3,787 | 0.4 | 18,152 | 1.9 | - | - | 19,559 | 2.0 |
| **Child and maternal data sources combined** (6 data sources)[3] | | | | | | | | | | | | | | | | |
| Any substance[4] | 64 | <0.1 | 964 | 0.1 | 3,229 | 0.3 | 2,203 | 0.2 | 12,067 | 1.2 | 26,045 | 2.7 | 13,975 | 1.4 | 32,647 | 3.4 |
| Alcohol | - | - | 511 | <0.1 | - | - | 1,431 | 0.1 | 3,074 | 0.3 | 10,702 | 1.1 | 11,175 | 1.2 | 13,637 | 1.4 |
| Other drug[5] | - | - | 473 | <0.1 | 3,229 | 0.3 | 814 | 0.1 | 10,282 | 1.1 | 18,152 | 1.9 | 4,272 | 0.4 | 23,485 | 2.4 |

1. For cause of death data, reporting is limited to the outcome "any substance use" due to low numbers in specific substance types; 2. Child protection data includes all carers 3. The outcomes are not mutually exclusive across data sources or substance types; therefore, numbers in rows/columns do not sum to numbers reported in "All records and data sources" (rows) or the "Any substance use"outcome (columns); 4. Any substance use includes the use of alcohol and or any drug use; 5. Other drug use includes the use of drugs other than alcohol and tobacco, inducing illicit drugs, misuse of prescription drugs, and use of opioid agonist treatment; 6. "Sedatives or other" includes the use of sedatives, hallucinogens, sedatives, solvents, or organic compounds, which were aggregated due to small cell sizes in individual substance categories; 7. "Unspecified or multiple substance use" includes ICD10AM codes that indicate multiple drug use or do not specify the type of drug or psychoactive substance.

**Supplement 16. Limitations of data sources used in analysis**

| Data source | Additional information, limitations and potential sources of bias |
|---|---|
| Perinatal data | • NSW perinatal data collection does not include children who are born very premature or underweight, and those who do not have a birth registration. However, both birth data sources have high population coverage and registration of births among disadvantaged populations has greatly improved over recent years. |
| NSW Department of Communities and Justice, child protection reports (child protection report data) | • Child protection records only represent a small number of the children who are maltreated and reported to child protection services, missing those children who are not reported.[1]<br>• Carer substance use may be under-ascertained in the prenatal period, as child protection records for prenatal reports do not included details on the reason for report, meaning that substance use could not be ascertained from *prenatal reports*. However, all children with a prenatal report in our study went on to have subsequent contact with child protection after pregnancy, when we were able to ascertain a reason for referral.<br>• Maternal substance use reported or assessed by child protection is included in carer drug and/or alcohol use issues recorded in child protection data; as such, it is not possible to differentiate maternal from other carer substance use from this data source. We addressed this limitation by: (i) noting the inclusion of carer substance use from child protection data in the maternal substance use outcome derived from health, death, and child protection data in results; (ii) using 'maternal and other carer' rather than 'maternal' substance use for estimates from child protection data alone; and (iii) conducting an additional analysis estimating the prevalence of maternal substance use derived only from health and death records.<br>• However, Australian data show in most cases of substantiated maltreatment the subjects of the investigation are mothers (79% of all neglect cases which includes carer substance use, 60% of all maltreatment cases).[2, 3] Furthermore, in this study at the time of birth 60% of the mothers in the maternal substance use group were single. |
| NSW Admitted Patient Data Collection (Hospital inpatient data) | • Surveillance bias may mean that some populations are more likely to be screened during pregnancy and at birth for substance use, such as women who absent, or present late to prenatal care, have low maternal age,[4, 5] or are among racially and ethnically minoritized populations.[5, 6] |
| NSW Emergency Department Data Collection (emergency department presentations) | • Emergency records only report the primary presenting reason, missing other conditions that may be contributing to the person's emergency presentation.<br>• There is also variation in electronic medical record systems and practices across NSW emergency departments, resulting in variable diagnosis coding practices.<br>• Primary presenting reasons coded using SNOMED-CT coding systems can represent symptoms or investigations, and are not restricted to diagnoses.<br>• The EDDC is not necessarily representative of all NSW emergency departments, as not all emergency departments contribute data to the emergency department data collection (EDDC). For example, only public hospitals contribute to the EDDC and less |

| | |
|---|---|
| | emergency departments in rural and regional areas contribute than in other areas. However, the proportion of participating emergency departments has increased, from around 30% in 1996 to around 60% in 2010. |
| NSW Mental Health Ambulatory Data Collection (Mental health outpatient data) | • Mental health outpatients' appointments only represent the publicly funded mental health services, missing a significant market of private health providers and services. |
| NSW Cause of Death Unit Record File Unit Record File (death registrations) | • Substance use may be under-ascertained in cause of death records as coroners do not routinely screen for all substances (e.g., volatile solvents) unless there is reason to suspect their use.[7] |
