## Supplement 5-5a for "Prevalence of maternal substance use during pregnancy and first two years of life: A whole-population cohort of 970,470 Australian children born 2008-2017"

### Supplement 5a. Definitions of ICD-10 AM and/or SNOMED CT[1] codes used to ascertain maternal substance use from four data sources available in the NSW child e-cohort

| Code description | Code | Analysis | Substance type |
|---|---|---|---|
| **ICD-10 AM codes** | | | |
| **Child records** | | | |
| P96.1 Neonatal withdrawal symptoms from maternal use of drugs of addiction | P96.1 | Main analysis | Other drug |
| P04.4 Fetus and newborn affected by maternal use of drugs of addiction | P04.4 | Main analysis | Other drug |
| P04.3 Fetus and newborn affected by maternal use of alcohol | P04.3 | Main analysis | Alcohol |
| Q86.0 Fetal alcohol syndrome (dysmorphic) | Q86.0 | Main analysis | Alcohol |
| **Mother records** | | | |
| **Alcohol** | | | |
| O35.4 Maternal care for (suspected) damage to fetus from alcohol | O35.4 | Main analysis | Alcohol |
| Z50.2 Alcohol rehabilitation | Z50.2 | Main analysis | Alcohol |
| Z72.1 Alcohol use | Z72.1 | Main analysis | Alcohol |
| R78.0 Finding of alcohol in blood | R78.0 | Main analysis | Alcohol |
| Y90 Evidence of alcohol involvement determined by blood alcohol level | Y90 | Main analysis | Alcohol |
| Y90.0 Blood alcohol level of less than 20 mg/100 ml | Y90.0 | Main analysis | Alcohol |
| Y90.1 Blood alcohol level of 20-39 mg/100 ml | Y90.1 | Main analysis | Alcohol |
| Y90.2 Blood alcohol level of 40-59 mg/100 ml | Y90.2 | Main analysis | Alcohol |
| Y90.3 Blood alcohol level of 60-79 mg/100 ml | Y90.3 | Main analysis | Alcohol |
| Y90.4 Blood alcohol level of 80-99 mg/100 ml | Y90.4 | Main analysis | Alcohol |
| Y90.5 Blood alcohol level of 100-119 mg/100 ml | Y90.5 | Main analysis | Alcohol |
| Y90.6 Blood alcohol level of 120-199 mg/100 ml | Y90.6 | Main analysis | Alcohol |
| Y90.7 Blood alcohol level of 200-239 mg/100 ml | Y90.7 | Main analysis | Alcohol |
| Y90.8 Blood alcohol level of 240 mg/100 ml or more | Y90.8 | Main analysis | Alcohol |
| Y90.9 Presence of alcohol in blood, level not specified | Y90.9 | Main analysis | Alcohol |
| Y91 Evidence of alcohol involvement determined by level of intoxication | Y91 | Main analysis | Alcohol |
| Y91.0 Mild alcohol intoxication | Y91.0 | Main analysis | Alcohol |
| Y91.1 Moderate alcohol intoxication | Y91.1 | Main analysis | Alcohol |
| Y91.2 Severe alcohol intoxication | Y91.2 | Main analysis | Alcohol |
| Y91.3 Very severe alcohol intoxication | Y91.3 | Main analysis | Alcohol |
| Y91.9 Alcohol involvement, not otherwise specified | Y91.9 | Main analysis | Alcohol |
| T51 Toxic effect of alcohol | T51 | Main analysis | Alcohol |
| T51.0 Ethanol | T51.0 | Main analysis | Alcohol |
| T51.1 Methanol | T51.1 | Main analysis | Alcohol |
| T51.2 2-Propanol | T51.2 | Main analysis | Alcohol |
| T51.8 Other alcohols | T51.8 | Main analysis | Alcohol |
| T51.9 Alcohol, unspecified | T51.9 | Main analysis | Alcohol |
| X45 Accidental poisoning by and exposure to alcohol | X45 | Main analysis | Alcohol |

| Code description | Code | Analysis | Substance type |
|---|---|---|---|
| X65 Intentional self-poisoning by and exposure to alcohol | X65 | Main analysis | Alcohol |
| Y15 Poisoning by and exposure to alcohol, undetermined intent | Y15 | Main analysis | Alcohol |
| F10 Mental and behavioural disorders due to use of alcohol | F10 | Main analysis | Alcohol |
| F10.0 Mental and behavioural disorders due to use of alcohol, acute intoxication | F10.0 | Main analysis | Alcohol |
| F10.1 Mental and behavioural disorders due to use of alcohol, harmful use | F10.1 | Main analysis | Alcohol |
| F10.2 Mental and behavioural disorders due to use of alcohol, dependence syndrome | F10.2 | Main analysis | Alcohol |
| F10.3 Mental and behavioural disorders due to use of alcohol, withdrawal state | F10.3 | Main analysis | Alcohol |
| F10.4 Mental and behavioural disorders due to use of alcohol, withdrawal state with delirium | F10.4 | Main analysis | Alcohol |
| F10.5 Mental and behavioural disorders due to use of alcohol, psychotic disorder | F10.5 | Main analysis | Alcohol |
| F10.6 Mental and behavioural disorders due to use of alcohol, amnesic syndrome | F10.6 | Main analysis | Alcohol |
| F10.7 Mental and behavioural disorders due to use of alcohol, residual and late-onset psychotic disorder | F10.7 | Main analysis | Alcohol |
| F10.8 Mental and behavioural disorders due to use of alcohol, other mental and behavioural disorders | F10.8 | Main analysis | Alcohol |
| F10.9 Mental and behavioural disorders due to use of alcohol, unspecified mental and behavioural disorder | F10.9 | Main analysis | Alcohol |
| **Opioids** | | | |
| T40.0 Opium | T40.0 | Main analysis | Opioids |
| T40.1 Heroin | T40.1 | Main analysis | Opioids |
| T40.2 Other opioids | T40.2 | Main analysis | Opioids |
| T40.3 Methadone | T40.3 | Main analysis | Opioids |
| F11 Mental and behavioural disorders due to use of opioids | F11 | Main analysis | Opioids |
| F11.0 Mental and behavioural disorders due to use of opioids, acute intoxication | F11.0 | Main analysis | Opioids |
| F11.1 Mental and behavioural disorders due to use of opioids, harmful use | F11.1 | Main analysis | Opioids |
| F11.2 Mental and behavioural disorders due to use of opioids, dependence syndrome | F11.2 | Main analysis | Opioids |
| F11.3 Mental and behavioural disorders due to use of opioids, withdrawal state | F11.3 | Main analysis | Opioids |
| F11.4 Mental and behavioural disorders due to use of opioids, withdrawal state with delirium | F11.4 | Main analysis | Opioids |
| F11.5 Mental and behavioural disorders due to use of opioids, psychotic disorder | F11.5 | Main analysis | Opioids |
| F11.6 Mental and behavioural disorders due to use of opioids, amnesic syndrome | F11.6 | Main analysis | Opioids |
| F11.7 Mental and behavioural disorders due to use of opioids, residual and late-onset psychotic disorder | F11.7 | Main analysis | Opioids |
| F11.8 Mental and behavioural disorders due to use of opioids, other mental and behavioural disorders | F11.8 | Main analysis | Opioids |
| F11.9 Mental and behavioural disorders due to use of opioids, unspecified mental and behavioural disorder | F11.9 | Main analysis | Opioids |
| R78.1 Finding of opiate drug in blood | R78.1 | Sensitivity analysis | Opioids |
| **Cannabis/Cannabinoids** | | | |
| T40.7 Cannabis (derivatives) | T40.7 | Main analysis | Cannabis |
| F12 Mental and behavioural disorders due to use of cannabinoids | F12 | Main analysis | Cannabis |
| F12.0 Mental and behavioural disorders due to use of cannabinoids, acute intoxication | F12.0 | Main analysis | Cannabis |
| F12.1 Mental and behavioural disorders due to use of cannabinioids, harmful use | F12.1 | Main analysis | Cannabis |
| F12.2 Mental and behavioural disorders due to use of cannabinioids, dependence syndrome | F12.2 | Main analysis | Cannabis |
| F12.3 Mental and behavioural disorders due to use of cannabinioids, withdrawal state | F12.3 | Main analysis | Cannabis |
| F12.4 Mental and behavioural disorders due to use of cannabinioids, withdrawal state with delirium | F12.4 | Main analysis | Cannabis |

| Code description | Code | Analysis | Substance type |
|---|---|---|---|
| F12.5 Mental and behavioural disorders due to use of cannabinioids, psychotic disorder | F12.5 | Main analysis | Cannabis |
| F12.6 Mental and behavioural disorders due to use of cannabinioids, amnesic syndrome | F12.6 | Main analysis | Cannabis |
| F12.7 Mental and behavioural disorders due to use of cannabinioids, residual and late-onset psychotic disorder | F12.7 | Main analysis | Cannabis |
| F12.8 Mental and behavioural disorders due to use of cannabinioids, other mental and behavioural disorders | F12.8 | Main analysis | Cannabis |
| F12.9 Mental and behavioural disorders due to use of cannabinioids, unspecified mental and behavioural disorder | F12.9 | Main analysis | Cannabis |
| **STIMULANTS** | | | |
| **Cocaine** | | | |
| R78.2 Finding of cocaine in blood | R78.2 | Main analysis | Stimulants |
| T40.5 Cocaine | T40.5 | Main analysis | Stimulants |
| F14 Mental and behavioural disorders due to use of cocaine | F14 | Main analysis | Stimulants |
| F14.0Mental and behavioural disorders due to use of cocaine, acute intoxication | F14.0 | Main analysis | Stimulants |
| F14.1 Mental and behavioural disorders due to use of cocaine, harmful use | F14.1 | Main analysis | Stimulants |
| F14.2 Mental and behavioural disorders due to use of cocaine, dependence syndrome | F14.2 | Main analysis | Stimulants |
| F14.3 Mental and behavioural disorders due to use of cocaine, withdrawal state | F14.3 | Main analysis | Stimulants |
| F14.4 Mental and behavioural disorders due to use of cocaine, withdrawal state with delirium | F14.4 | Main analysis | Stimulants |
| F14.5 Mental and behavioural disorders due to use of cocaine, psychotic disorder | F14.5 | Main analysis | Stimulants |
| F14.6 Mental and behavioural disorders due to use of cocaine, amnesic syndrome | F14.6 | Main analysis | Stimulants |
| F14.7 Mental and behavioural disorders due to use of cocaine, residual and late-onset psychotic disorder | F14.7 | Main analysis | Stimulants |
| F14.8 Mental and behavioural disorders due to use of cocaine, other mental and behavioural disorders | F14.8 | Main analysis | Stimulants |
| F14.9 Mental and behavioural disorders due to use of cocaine, unspecified mental and behavioural disorder | F14.9 | Main analysis | Stimulants |
| **Other stimulants** | | | |
| T43.69 Other psychostimulants with potential for use disorder | T43.69 | Main analysis | Stimulants |
| F15 Mental and behavioural disorders due to use other stimulants, including caffeine | F15 | Main analysis | Stimulants |
| F15.0 Mental and behavioural disorders due to use other stimulants including caffeine, acute intoxication | F15.0 | Main analysis | Stimulants |
| F15.00 Mental and behavioural disorders due to use other stimulants including caffeine, acute intoxication, unspecified stimulants | F15.00 | Main analysis | Stimulants |
| F15.01 Mental and behavioural disorders due to use other stimulants including caffeine, acute intoxication, methylamphetamine | F15.01 | Main analysis | Stimulants |
| F15.02 Mental and behavioural disorders due to use other stimulants including caffeine, acute intoxication, methylenedioxy methylamphetamine | F15.02 | Main analysis | Stimulants |
| F15.09 Mental and behavioural disorders due to use other stimulants including caffeine, acute intoxication, other specified stimulants | F15.09 | Main analysis | Stimulants |
| F15.1 Mental and behavioural disorders due to use other stimulants including caffeine, harmful use | F15.1 | Main analysis | Stimulants |
| F15.10 Mental and behavioural disorders due to use other stimulants including caffeine, harmful use, unspecified stimulants | F15.10 | Main analysis | Stimulants |
| F15.11 Mental and behavioural disorders due to use other stimulants including caffeine, harmful use, methylamphetamine | F15.11 | Main analysis | Stimulants |
| F15.12 Mental and behavioural disorders due to use other stimulants including caffeine, harmful use, methylenedioxy methylamphetamine | F15.12 | Main analysis | Stimulants |
| F15.19 Mental and behavioural disorders due to use other stimulants including caffeine, harmful use, other specified stimulants | F15.19 | Main analysis | Stimulants |
| F15.2 Mental and behavioural disorders due to use other stimulants including caffeine, dependence syndrome | F15.2 | Main analysis | Stimulants |
| F15.20 Mental and behavioural disorders due to use other stimulants including caffeine, dependence syndrome, unspecified stimulants | F15.20 | Main analysis | Stimulants |
| F15.21 Mental and behavioural disorders due to use other stimulants including caffeine, dependence syndrome, methylamphetamine | F15.21 | Main analysis | Stimulants |

| Code description | Code | Analysis | Substance type |
|---|---|---|---|
| F15.22 Mental and behavioural disorders due to use other stimulants including caffeine, dependence syndrome, methylenedioxy methylamphetamine | F15.22 | Main analysis | Stimulants |
| F15.29 Mental and behavioural disorders due to use other stimulants including caffeine, dependence syndrome, other specified stimulants | F15.29 | Main analysis | Stimulants |
| F15.3 Mental and behavioural disorders due to use other stimulants including caffeine, withdrawal state | F15.3 | Main analysis | Stimulants |
| F15.30 Mental and behavioural disorders due to use other stimulants including caffeine, withdrawal state, unspecified stimulants | F15.30 | Main analysis | Stimulants |
| F15.31 Mental and behavioural disorders due to use other stimulants including caffeine, withdrawal state, methylamphetamine | F15.31 | Main analysis | Stimulants |
| F15.32 Mental and behavioural disorders due to use other stimulants including caffeine, withdrawal state, methylenedioxy methylamphetamine | F15.32 | Main analysis | Stimulants |
| F15.39 Mental and behavioural disorders due to use other stimulants including caffeine, withdrawal state, other specified stimulants | F15.39 | Main analysis | Stimulants |
| F15.4 Mental and behavioural disorders due to use other stimulants including caffeine, withdrawal state with delirium | F15.4 | Main analysis | Stimulants |
| F15.40 Mental and behavioural disorders due to use other stimulants including caffeine, withdrawal state with delirium, unspecified stimulants | F15.40 | Main analysis | Stimulants |
| F15.41 Mental and behavioural disorders due to use other stimulants including caffeine, withdrawal state with delirium, methylamphetamine | F15.41 | Main analysis | Stimulants |
| F15.42 Mental and behavioural disorders due to use other stimulants including caffeine, withdrawal state with delirium, methylamphetamine | F15.42 | Main analysis | Stimulants |
| F15.49 Mental and behavioural disorders due to use other stimulants including caffeine, withdrawal state with delirium, other specified stimulants | F15.49 | Main analysis | Stimulants |
| F15.5Mental and behavioural disorders due to use other stimulants including caffeine, psychotic disorder | F15.5 | Main analysis | Stimulants |
| F15.50 Mental and behavioural disorders due to use other stimulants including caffeine, psychotic disorder, unspecified stimulants | F15.50 | Main analysis | Stimulants |
| F15.51 Mental and behavioural disorders due to use other stimulants including caffeine, psychotic disorder, methylamphetamine | F15.51 | Main analysis | Stimulants |
| F15.52 Mental and behavioural disorders due to use other stimulants including caffeine, psychotic disorder, methylenedioxy methylamphetamine | F15.52 | Main analysis | Stimulants |
| F15.59 Mental and behavioural disorders due to use other stimulants including caffeine, psychotic disorder, other specified stimulants | F15.59 | Main analysis | Stimulants |
| F15.6 Mental and behavioural disorders due to use other stimulants including caffeine, amnesic syndrome | F15.6 | Main analysis | Stimulants |
| F15.60 Mental and behavioural disorders due to use other stimulants including caffeine, amnesic syndrome, unspecified stimulants | F15.60 | Main analysis | Stimulants |
| F15.61 Mental and behavioural disorders due to use other stimulants including caffeine, amnesic syndrome, methlyamphetamine | F15.61 | Main analysis | Stimulants |
| F15.62 Mental and behavioural disorders due to use other stimulants including caffeine, amnesic syndrome, methylenedioxy methlyamphetamine | F15.62 | Main analysis | Stimulants |
| F15.69 Mental and behavioural disorders due to use other stimulants including caffeine, amnesic syndrome, other specified stimulants | F15.69 | Main analysis | Stimulants |
| F15.7Mental and behavioural disorders due to use other stimulants including caffeine, residual and late-onset psychotic disorder | F15.7 | Main analysis | Stimulants |
| F15.70 Mental and behavioural disorders due to use other stimulants including caffeine, residual and late-onset psychotic disorder, unspecified stimulants | F15.70 | Main analysis | Stimulants |
| F15.71 Mental and behavioural disorders due to use other stimulants including caffeine, residual and late-onset psychotic disorder, methylamphetamine | F15.71 | Main analysis | Stimulants |
| F15.72 Mental and behavioural disorders due to use other stimulants including caffeine, residual and late-onset psychotic disorder, methylenedioxy methylamphetamine | F15.72 | Main analysis | Stimulants |
| F15.79 Mental and behavioural disorders due to use other stimulants including caffeine, residual and late-onset psychotic disorder, other specified stimulants | F15.79 | Main analysis | Stimulants |
| F15.8 Mental and behavioural disorders due to use other stimulants including caffeine, other mental and behavioural disorders | F15.8 | Main analysis | Stimulants |

| Code description | Code | Analysis | Substance type |
|---|---|---|---|
| F15.80 Mental and behavioural disorders due to use other stimulants including caffeine, other mental and behavioural disorders, unspecified stimulants | F15.80 | Main analysis | Stimulants |
| F15.81 Mental and behavioural disorders due to use other stimulants including caffeine, other mental and behavioural disorders, methylamphetamine | F15.81 | Main analysis | Stimulants |
| F15.82 Mental and behavioural disorders due to use other stimulants including caffeine, other mental and behavioural disorders, methylenedioxy methylamphetamine | F15.82 | Main analysis | Stimulants |
| F15.89 Mental and behavioural disorders due to use other stimulants including caffeine, other mental and behavioural disorders, other specified stimulants | F15.89 | Main analysis | Stimulants |
| F15.9 Mental and behavioural disorders due to use other stimulants including caffeine, unspecified mental and behavioural disorder | F15.9 | Main analysis | Stimulants |
| F15.90 Mental and behavioural disorders due to use other stimulants including caffeine, unspecified mental and behavioural disorder, unspecified stimulants | F15.90 | Main analysis | Stimulants |
| F15.91 Mental and behavioural disorders due to use other stimulants including caffeine, unspecified mental and behavioural disorder, methylamphetamine | F15.91 | Main analysis | Stimulants |
| F15.92 Mental and behavioural disorders due to use other stimulants including caffeine, unspecified mental and behavioural disorder, methylenedioxy methylamphetamine | F15.92 | Main analysis | Stimulants |
| F15.99 Mental and behavioural disorders due to use other stimulants including caffeine, unspecified mental and behavioural disorder, other specified stimulants | F15.99 | Main analysis | Stimulants |
| T43.6 Psychostimulants with potential for use disorder | T43.6 | Sensitivity analysis | Stimulants |
| T43.60 Unspecified psychostimulants with potential for use disorder | T43.60 | Sensitivity analysis | Stimulants |
| **Methamphetamines** | | | |
| T43.61 Methylamphetamine | T43.61 | Main analysis | Stimulants |
| T43.62 Methylenedioxy methamphetamine | T43.62 | Main analysis | Stimulants |
| **Sedatives or Other** | | | |
| **Sedatives, and hypnotics,** | | | |
| F13 Mental and behavioural disorders due to use of sedatives or hypnotics | F13 | Main analysis | Sedatives and others |
| F13.0 Mental and behavioural disorders due to use of sedatives or hypnotics, acute intoxication | F13.0 | Main analysis | Sedatives and others |
| F13.00 Mental and behavioural disorders due to use of sedatives or hypnotics, acute intoxication, unspecified sedative or hypnotic | F13.00 | Main analysis | Sedatives and others |
| F13.01 Mental and behavioural disorders due to use of sedatives or hypnotics, acute intoxication, gamma hydroxybutyrate | F13.01 | Main analysis | Sedatives and others |
| F13.09 Mental and behavioural disorders due to use of sedatives or hypnotics, acute intoxication, other specified sedative or hypnotic | F13.09 | Main analysis | Sedatives and others |
| F13.1 Mental and behavioural disorders due to use of sedatives or hypnotics, harmful use | F13.1 | Main analysis | Sedatives and others |
| F13.10 Mental and behavioural disorders due to use of sedatives or hypnotics, harmful use, unspecified sedative or hypnotic | F13.10 | Main analysis | Sedatives and others |
| F13.11 Mental and behavioural disorders due to use of sedatives or hypnotics, harmful use, gamma hydroxybutyrate | F13.11 | Main analysis | Sedatives and others |
| F13.19 Mental and behavioural disorders due to use of sedatives or hypnotics, harmful use, other specified sedative or hypnotic | F13.19 | Main analysis | Sedatives and others |
| F13.2 Mental and behavioural disorders due to use of sedatives or hypnotics, dependence syndrome | F13.2 | Main analysis | Sedatives and others |
| F13.20 Mental and behavioural disorders due to use of sedatives or hypnotics, dependence syndrome, unspecified sedative or hypnotic | F13.20 | Main analysis | Sedatives and others |
| F13.21 Mental and behavioural disorders due to use of sedatives or hypnotics, dependence syndrome, gamma hydroxybutyrate | F13.21 | Main analysis | Sedatives and others |
| F13.29 Mental and behavioural disorders due to use of sedatives or hypnotics, dependence syndrome, other specified sedative or hypnotic | F13.29 | Main analysis | Sedatives and others |
| F13.3Mental and behavioural disorders due to use of sedatives or hypnotics, withdrawal state | F13.3 | Main analysis | Sedatives and others |
| F13.30 Mental and behavioural disorders due to use of sedatives or hypnotics, withdrawal state, unspecified sedative or hypnotic | F13.30 | Main analysis | Sedatives and others |

| Code description | Code | Analysis | Substance type |
|---|---|---|---|
| F13.31 Mental and behavioural disorders due to use of sedatives or hypnotics, withdrawal state, gamma hydroxybutyrate | F13.31 | Main analysis | Sedatives and others |
| F13.39 Mental and behavioural disorders due to use of sedatives or hypnotics, withdrawal state, other specified sedative or hypnotic | F13.39 | Main analysis | Sedatives and others |
| F13.4 Mental and behavioural disorders due to use of sedatives or hypnotics, withdrawal state with delirium | F13.4 | Main analysis | Sedatives and others |
| F13.40 Mental and behavioural disorders due to use of sedatives or hypnotics, withdrawal state with delirium, unspecified sedative or hypnotic | F13.40 | Main analysis | Sedatives and others |
| F13.41 Mental and behavioural disorders due to use of sedatives or hypnotics, withdrawal state with delirium, gamma hydroxybutyrate | F13.41 | Main analysis | Sedatives and others |
| F13.49 Mental and behavioural disorders due to use of sedatives or hypnotics, withdrawal state with delirium, other specified sedative or hypnotic | F13.49 | Main analysis | Sedatives and others |
| F13.5Mental and behavioural disorders due to use of sedatives or hypnotics, psychotic disorder | F13.5 | Main analysis | Sedatives and others |
| F13.50 Mental and behavioural disorders due to use of sedatives or hypnotics, psychotic disorder, unspecified sedative or hypnotic | F13.50 | Main analysis | Sedatives and others |
| F13.51 Mental and behavioural disorders due to use of sedatives or hypnotics, psychotic disorder, gamma hydroxybutyrate | F13.51 | Main analysis | Sedatives and others |
| F13.59 Mental and behavioural disorders due to use of sedatives or hypnotics, psychotic disorder, other specified sedative or hypnotic | F13.59 | Main analysis | Sedatives and others |
| F13.6 Mental and behavioural disorders due to use of sedatives or hypnotics, amnesic syndrome | F13.6 | Main analysis | Sedatives and others |
| F13.60 Mental and behavioural disorders due to use of sedatives or hypnotics, amnesic syndrome, unspecified sedative or hypnotic | F13.60 | Main analysis | Sedatives and others |
| F13.61 Mental and behavioural disorders due to use of sedatives or hypnotics, amnesic syndrome, gamma hydroxybutyrate | F13.61 | Main analysis | Sedatives and others |
| F13.69 Mental and behavioural disorders due to use of sedatives or hypnotics, amnesic syndrome, other specified sedative or hypnotic | F13.69 | Main analysis | Sedatives and others |
| F13.7Mental and behavioural disorders due to use of sedatives or hypnotics, residual and late-onset psychotic disorder | F13.7 | Main analysis | Sedatives and others |
| F13.70 Mental and behavioural disorders due to use of sedatives or hypnotics, residual and late-onset psychotic disorder, unspecified sedative or hypnotic | F13.70 | Main analysis | Sedatives and others |
| F13.71 Mental and behavioural disorders due to use of sedatives or hypnotics, residual and late-onset psychotic disorder, gamma hydroxybutyrate | F13.71 | Main analysis | Sedatives and others |
| F13.79 Mental and behavioural disorders due to use of sedatives or hypnotics, residual and late-onset psychotic disorder, other specified sedative or hypnotic | F13.79 | Main analysis | Sedatives and others |
| F13.8 Mental and behavioural disorders due to use of sedatives or hypnotics, other mental and behavioural disorders | F13.8 | Main analysis | Sedatives and others |
| F13.80 Mental and behavioural disorders due to use of sedatives or hypnotics, other mental and behavioural disorders, unspecified sedative or hypnotic | F13.80 | Main analysis | Sedatives and others |
| F13.81 Mental and behavioural disorders due to use of sedatives or hypnotics, other mental and behavioural disorders, gamma hydroxybutyrate | F13.81 | Main analysis | Sedatives and others |
| F13.89 Mental and behavioural disorders due to use of sedatives or hypnotics, other mental and behavioural disorders, other specified sedative or hypnotic | F13.89 | Main analysis | Sedatives and others |
| F13.9 Mental and behavioural disorders due to use of sedatives or hypnotics, unspecified mental and behavioural disorder | F13.9 | Main analysis | Sedatives and others |
| F13.90 Mental and behavioural disorders due to use of sedatives or hypnotics, unspecified mental and behavioural disorder, unspecified sedative or hypnotic | F13.90 | Main analysis | Sedatives and others |
| F13.91 Mental and behavioural disorders due to use of sedatives or hypnotics, unspecified mental and behavioural disorder, gamma hydroxybutyrate | F13.91 | Main analysis | Sedatives and others |
| F13.99 Mental and behavioural disorders due to use of sedatives or hypnotics, unspecified mental and behavioural disorder, other specified sedative or hypnotic | F13.99 | Main analysis | Sedatives and others |
| T42 Poisoning by antiepileptic, sedative-hypnotic and antiparkinsonism drugs | T42 | Sensitivity analysis | Sedatives and others |
| T42.6 Other antiepileptic and sedative-hypnotic drugs | T42.6 | Sensitivity analysis | Sedatives and others |
| T42.7 Antiepileptic and sedative-hypnotic drugs, unspecified | T42.7 | Sensitivity analysis | Sedatives and others |
| T42.3 Barbiturates | T42.3 | Sensitivity analysis | Sedatives and others |

| Code description | Code | Analysis | Substance type |
|---|---|---|---|
| T42.4 Benzodiazepines | T42.4 | Sensitivity analysis | Sedatives and others |
| **Hallucinogens** | | | |
| R78.3 Finding of hallucinogen in blood | R78.3 | Main analysis | Sedatives and others |
| T40.9 Other and unspecified psychodysleptics [hallucinogens] | T40.9 | Main analysis | Sedatives and others |
| F16 Mental and behavioural disorders due to use hallucinogens | F16 | Main analysis | Sedatives and others |
| F16.0 Mental and behavioural disorders due to use hallucinogens, acute intoxication | F16.0 | Main analysis | Sedatives and others |
| F16.00 Mental and behavioural disorders due to use hallucinogens, acute intoxication, unspecified hallucinogen | F16.00 | Main analysis | Sedatives and others |
| F16.01 Mental and behavioural disorders due to use hallucinogens, acute intoxication, ketamine | F16.01 | Main analysis | Sedatives and others |
| F16.09 Mental and behavioural disorders due to use hallucinogens, acute intoxication, other specified hallucinogen | F16.09 | Main analysis | Sedatives and others |
| F16.1 Mental and behavioural disorders due to use hallucinogens, harmful use | F16.1 | Main analysis | Sedatives and others |
| F16.10 Mental and behavioural disorders due to use hallucinogens, harmful use, unspecified hallucinogen | F16.10 | Main analysis | Sedatives and others |
| F16.11 Mental and behavioural disorders due to use hallucinogens, harmful use, ketamine | F16.11 | Main analysis | Sedatives and others |
| F16.19 Mental and behavioural disorders due to use hallucinogens, harmful use, other specified hallucinogen | F16.19 | Main analysis | Sedatives and others |
| F16.2 Mental and behavioural disorders due to use hallucinogens, dependence syndrome | F16.2 | Main analysis | Sedatives and others |
| F16.20 Mental and behavioural disorders due to use hallucinogens, dependence syndrome, unspecified hallucinogen | F16.20 | Main analysis | Sedatives and others |
| F16.21 Mental and behavioural disorders due to use hallucinogens, dependence syndrome, ketamine | F16.21 | Main analysis | Sedatives and others |
| F16.29 Mental and behavioural disorders due to use hallucinogens, dependence syndrome, other specified hallucinogen | F16.29 | Main analysis | Sedatives and others |
| F16.3 Mental and behavioural disorders due to use hallucinogens, withdrawal state | F16.3 | Main analysis | Sedatives and others |
| F16.30 Mental and behavioural disorders due to use hallucinogens, withdrawal state, unspecified hallucinogen | F16.30 | Main analysis | Sedatives and others |
| F16.31 Mental and behavioural disorders due to use hallucinogens, withdrawal state, Ketamine | F16.31 | Main analysis | Sedatives and others |
| F16.39 Mental and behavioural disorders due to use hallucinogens, withdrawal state, other specified hallucinogen | F16.39 | Main analysis | Sedatives and others |
| F16.4 Mental and behavioural disorders due to use hallucinogens, withdrawal state with delirium | F16.4 | Main analysis | Sedatives and others |
| F16.40 Mental and behavioural disorders due to use hallucinogens, withdrawal state with delirium, unspecified hallucinogen | F16.40 | Main analysis | Sedatives and others |
| F16.41 Mental and behavioural disorders due to use hallucinogens, withdrawal state with delirium, ketamine | F16.41 | Main analysis | Sedatives and others |
| F16.49 Mental and behavioural disorders due to use hallucinogens, withdrawal state with delirium, other specified hallucinogen | F16.49 | Main analysis | Sedatives and others |
| F16.5 Mental and behavioural disorders due to use hallucinogens, psychotic disorder | F16.5 | Main analysis | Sedatives and others |
| F16.50 Mental and behavioural disorders due to use hallucinogens, psychotic disorder, unspecified hallucinogen | F16.50 | Main analysis | Sedatives and others |
| F16.51 Mental and behavioural disorders due to use hallucinogens, psychotic disorder, ketamine | F16.51 | Main analysis | Sedatives and others |
| F16.59 Mental and behavioural disorders due to use hallucinogens, psychotic disorder, other specified hallucinogen | F16.59 | Main analysis | Sedatives and others |
| F16.6 Mental and behavioural disorders due to use hallucinogens, amnesic syndrome | F16.6 | Main analysis | Sedatives and others |
| F16.60 Mental and behavioural disorders due to use hallucinogens, amnesic syndrome, unspecified hallucinogen | F16.60 | Main analysis | Sedatives and others |
| F16.61 Mental and behavioural disorders due to use hallucinogens, amnesic syndrome, ketamine | F16.61 | Main analysis | Sedatives and others |
| F16.69 Mental and behavioural disorders due to use hallucinogens, amnesic syndrome, other specified hallucinogen | F16.69 | Main analysis | Sedatives and others |
| F16.7 Mental and behavioural disorders due to use hallucinogens, residual and late-onset psychotic disorder | F16.7 | Main analysis | Sedatives and others |
| F16.70 Mental and behavioural disorders due to use hallucinogens, residual and late-onset psychotic disorder, unspecified hallucinogen | F16.70 | Main analysis | Sedatives and others |
| F16.71 Mental and behavioural disorders due to use hallucinogens, residual and late-onset psychotic disorder, ketamine | F16.71 | Main analysis | Sedatives and others |
| F16.79 Mental and behavioural disorders due to use hallucinogens, residual and late-onset psychotic disorder, other specified hallucinogen | F16.79 | Main analysis | Sedatives and others |
| F16.8 Mental and behavioural disorders due to use hallucinogens, other mental and behavioural disorders | F16.8 | Main analysis | Sedatives and others |

| Code description | Code | Analysis | Substance type |
|---|---|---|---|
| F16.80 Mental and behavioural disorders due to use hallucinogens, other mental and behavioural disorders, unspecified hallucinogen | F16.80 | Main analysis | Sedatives and others |
| F16.81 Mental and behavioural disorders due to use hallucinogens, other mental and behavioural disorders, ketamine | F16.81 | Main analysis | Sedatives and others |
| F16.89 Mental and behavioural disorders due to use hallucinogens, other mental and behavioural disorders, other specified hallucinogen | F16.89 | Main analysis | Sedatives and others |
| F16.9 Mental and behavioural disorders due to use hallucinogens, unspecified mental and behavioural disorder | F16.9 | Main analysis | Sedatives and others |
| F16.91 Mental and behavioural disorders due to use hallucinogens, unspecified mental and behavioural disorder, ketamine | F16.91 | Main analysis | Sedatives and others |
| F16.99 Mental and behavioural disorders due to use hallucinogens, unspecified mental and behavioural disorder, other specified hallucinogen | F16.99 | Main analysis | Sedatives and others |
| T40.8 Lysergide [LSD] | T40.8 | Main analysis | Sedatives and others |
| T41.22 Ketamine | T41.22 | Sensitivity analysis | Sedatives and others |
| **Unspecified or multiple** | | | |
| **Drug unspecified or multiple** | | | |
| O35.5 Maternal care for (suspected) damage to fetus by drugs ("drug addiction" in WHO) | O35.5 | Main analysis | Drug unspecified or multiple |
| Z50.3 Drug rehabilitation | Z50.3 | Main analysis | Drug unspecified or multiple |
| Z72.2 Drug use | Z72.2 | Main analysis | Drug unspecified or multiple |
| R78 Findings of drugs and other substances, not normally found in blood | R78 | Main analysis | Drug unspecified or multiple |
| R78.4 Finding of other drugs of addictive potential in blood | R78.4 | Sensitivity analysis | Drug unspecified or multiple |
| **Psychotropic drug** | | | |
| R78.5 Finding of psychotropic drug in blood | R78.5 | Sensitivity analysis | Drug unspecified or multiple |
| T43.9 Psychotropic drug, unspecified | T43.9 | Sensitivity analysis | Drug unspecified or multiple |
| T43 Poisoning by psychotropic drugs, not elsewhere classified | T43 | Sensitivity analysis | Drug unspecified or multiple |
| T43.8 Other psychotropic drugs, not elsewhere classified | T43.8 | Sensitivity analysis | Drug unspecified or multiple |
| **Multiple drug** | | | |
| F19 Mental and behavioural disorders due to multiple drug use and use other psychoactive substances | F19 | Main analysis | Drug unspecified or multiple |
| F19.0 Mental and behavioural disorders due to multiple drug use and use of psychoactive substances, acute intoxication | F19.0 | Main analysis | Drug unspecified or multiple |
| F19.1 Mental and behavioural disorders due to multiple drug use and use of psychoactive substances, harmful use | F19.1 | Main analysis | Drug unspecified or multiple |
| F19.2 Mental and behavioural disorders due to multiple drug use and use of psychoactive substances, dependence syndrome | F19.2 | Main analysis | Drug unspecified or multiple |
| F19.3 Mental and behavioural disorders due to multiple drug use and use of psychoactive substances, withdrawal state | F19.3 | Main analysis | Drug unspecified or multiple |
| F19.4 Mental and behavioural disorders due to multiple drug use and use of psychoactive substances, withdrawal state with delirium | F19.4 | Main analysis | Drug unspecified or multiple |
| F19.5 Mental and behavioural disorders due to multiple drug use and use of psychoactive substances, psychotic disorder | F19.5 | Main analysis | Drug unspecified or multiple |
| F19.6 Mental and behavioural disorders due to multiple drug use and use of psychoactive substances, amnesic syndrome | F19.6 | Main analysis | Drug unspecified or multiple |
| F19.7 Mental and behavioural disorders due to multiple drug use and use of psychoactive substances, residual and late-onset psychotic disorder | F19.7 | Main analysis | Drug unspecified or multiple |
| F19.8 Mental and behavioural disorders due to multiple drug use and use of psychoactive substances, other mental and behavioural disorders | F19.8 | Main analysis | Drug unspecified or multiple |
| F19.9 Mental and behavioural disorders due to multiple drug use and use of psychoactive substances, unspecified mental and behavioural disorder | F19.9 | Main analysis | Drug unspecified or multiple |
| **Narcotics and psychodysleptics** | | | |
| T40 Poisoning by narcotics and psychodysleptics [hallucinogens] | T40 | Main analysis | Drug unspecified or multiple |

| Code description | Code | Analysis | Substance type |
|---|---|---|---|
| T40.4 Other synthetic narcotics | T40.4 | Main analysis | Drug unspecified or multiple |
| T40.6 Other and unspecified narcotics | T40.6 | Main analysis | Drug unspecified or multiple |
| **Solvents** | | | |
| F18 Mental and behavioural disorders due to use of volatile solvents | F18 | Main analysis | Sedatives and others |
| F18.0 Mental and behavioural disorders due to use of volatile solvents, acute intoxication | F18.0 | Main analysis | Sedatives and others |
| F18.1 Mental and behavioural disorders due to use of volatile solvents, harmful use | F18.1 | Main analysis | Sedatives and others |
| F18.2 Mental and behavioural disorders due to use of volatile solvents, dependence syndrome | F18.2 | Main analysis | Sedatives and others |
| F18.3 Mental and behavioural disorders due to use of volatile solvents, withdrawal state | F18.3 | Main analysis | Sedatives and others |
| F18.4 Mental and behavioural disorders due to use of volatile solvents, withdrawal state with delirium | F18.4 | Main analysis | Sedatives and others |
| F18.5 Mental and behavioural disorders due to use of volatile solvents, psychotic disorder | F18.5 | Main analysis | Sedatives and others |
| F18.6 Mental and behavioural disorders due to use of volatile solvents, amnesic syndrome | F18.6 | Main analysis | Sedatives and others |
| F18.7 Mental and behavioural disorders due to use of volatile solvents, residual and late-onset psychotic disorder | F18.7 | Main analysis | Sedatives and others |
| F18.8 Mental and behavioural disorders due to use of volatile solvents, other mental and behavioural disorders | F18.8 | Main analysis | Sedatives and others |
| F18.9 Mental and behavioural disorders due to use of volatile solvents, unspecified mental and behavioural disorder | F18.9 | Main analysis | Sedatives and others |
| T52 Toxic effect of organic solvents | T52 | Main analysis | Sedatives and others |
| T52.0 Petroleum products | T52.0 | Main analysis | Sedatives and others |
| T52.1 Benzene | T52.1 | Main analysis | Sedatives and others |
| T52.8 Other organic solvents | T52.8 | Main analysis | Sedatives and others |
| T52.9 Organic solvent, unspecified | T52.9 | Main analysis | Sedatives and others |

### SNOMED CT-Codes

| Definition | SNOMED code | ICD-10 Equivalent[2] | Analysis | Substance type |
|---|---|---|---|---|
| | **Alcohol** | | | **Alcohol** |
| Alcohol rehabilitation and detoxification | 20093000 | | Main analysis | Alcohol |
| Alcohol intoxication | 25702006 | | Main analysis | Alcohol |
| Alcohol dependence | 66590003 | | Main analysis | Alcohol |
| Nondependent alcohol abuse, continuous | 191882002 | | Main analysis | Alcohol |
| Nondependent alcohol abuse, episodic | 191883007 | | Main analysis | Alcohol |
| Nondependent cannabis abuse | 191891003 | | Main analysis | Alcohol |
| Total time drunk alcohol | 228330005 | | Main analysis | Alcohol |
| Absinthe addiction | 231467000 | | Main analysis | Alcohol |
| Nondependent alcohol abuse | 268645007 | | Main analysis | Alcohol |
| Persistent alcohol abuse | 284591009 | | Main analysis | Alcohol |
| Methanol abuse | 304605000 | | Main analysis | Alcohol |
| Substance use treatment: alcohol withdrawal | 386449006 | | Main analysis | Alcohol |
| Severe alcohol dependence | 713862009 | | Main analysis | Alcohol |
| Alcohol detoxification | 827094004 | | Main analysis | Alcohol |

**SNOMED CT-Codes**

| Definition | SNOMED code | ICD-10 Equivalent[2] | Analysis | Substance type |
|---|---|---|---|---|
| Acute alcohol intoxication | 1149333003 | | Main analysis | Alcohol |
| Alcohol dependence in pregnancy | 10741871000119100 | | Main analysis | Alcohol |
| Toxic effect of butyl alcohol | 4953006 | T51 | Main analysis | Alcohol |
| Toxic effect of propyl alcohol | 6749002 | T51 | Main analysis | Alcohol |
| Alcohol hallucinosis | 7052005 | F10 | Main analysis | Alcohol |
| Alcoholism | 7200002 | F10 | Main analysis | Alcohol |
| Alcohol withdrawal delirium | 8635005 | F10 | Main analysis | Alcohol |
| Alcohol intoxication delirium | 18653004 | F10 | Main analysis | Alcohol |
| Idiosyncratic intoxication | 21000000 | F10 | Main analysis | Alcohol |
| Metabolic acidosis due to methanol | 25966003 | T51 | Main analysis | Alcohol |
| Alcohol-induced organic mental disorder | 29212009 | F10 | Main analysis | Alcohol |
| Hangover | 32553006 | F10 | Main analysis | Alcohol |
| Alcohol-induced anxiety disorder | 34938008 | F10 | Main analysis | Alcohol |
| Alcohol-induced sleep disorder | 41083005 | F10 | Main analysis | Alcohol |
| Alcohol-induced psychosis | 42344001 | F10 | Main analysis | Alcohol |
| Alcohol-induced mood disorder | 53936005 | F10 | Main analysis | Alcohol |
| Toxic effect of fusel oil | 57346004 | T51 | Main analysis | Alcohol |
| Alcohol-induced psychotic disorder with delusions | 61144001 | F10 | Main analysis | Alcohol |
| Toxic effect of alcohol | 67426006 | T51 | Main analysis | Alcohol |
| Alcohol-induced sexual dysfunction | 78524005 | F10 | Main analysis | Alcohol |
| Alcohol paranoia | 79578000 | F10 | Main analysis | Alcohol |
| Diarrhoea due to alcohol intake | 82047000 | T51 | Main analysis | Alcohol |
| Alcohol poisoning | 82782008 | T51 | Main analysis | Alcohol |
| Uncomplicated alcohol withdrawal | 85561006 | F10 | Main analysis | Alcohol |
| Toxic effect of amyl alcohol | 87460008 | T51 | Main analysis | Alcohol |
| Toxic effect of denatured alcohol | 89507002 | T51 | Main analysis | Alcohol |
| Drug interaction with alcohol | 95906008 | T51 | Main analysis | Alcohol |
| Alcohol intake above recommended sensible limits | 160592001 | F10 | Main analysis | Alcohol |
| Maternal alcohol abuse | 169942003 | F10 | Main analysis | Alcohol |
| Chronic alcoholic brain syndrome | 191475009 | F10 | Main analysis | Alcohol |
| Alcohol withdrawal hallucinosis | 191476005 | F10 | Main analysis | Alcohol |
| Pathological alcohol intoxication | 191477001 | F10 | Main analysis | Alcohol |
| Alcoholic paranoia | 191478006 | F10 | Main analysis | Alcohol |
| Alcohol withdrawal syndrome | 191480000 | F10 | Main analysis | Alcohol |
| Acute alcoholic intoxication in alcoholism | 191802004 | F10 | Main analysis | Alcohol |

| SNOMED CT-Codes | | | | |
|---|---|---|---|---|
| **Definition** | **SNOMED code** | **ICD-10 Equivalent[2]** | **Analysis** | **Substance type** |
| Continuous acute alcoholic intoxication in alcoholism | 191804003 | F10 | Main analysis | Alcohol |
| Episodic acute alcoholic intoxication in alcoholism | 191805002 | F10 | Main analysis | Alcohol |
| Continuous chronic alcoholism | 191811004 | F10 | Main analysis | Alcohol |
| Episodic chronic alcoholism | 191812006 | F10 | Main analysis | Alcohol |
| Mental and behavioral disorders due to use of alcohol (disorder) | 192206005 | F10 | Main analysis | Alcohol |
| Mental and behavioral disorders due to use of alcohol: acute intoxication (disorder) | 192207001 | F10 | Main analysis | Alcohol |
| Mental and behavioral disorders due to use of alcohol: harmful use (disorder) | 192208006 | F10 | Main analysis | Alcohol |
| Mental and behavioural disorders due to use of alcohol: dependence syndrome) or (chronic alcoholism [& (addiction) or (dipsomania)]) (disorder) | 192209003 | F10 | Main analysis | Alcohol |
| Mental and behavioral disorders due to use of alcohol: withdrawal state (disorder) | 192210008 | F10 | Main analysis | Alcohol |
| Mental and behavioral disorders due to use of alcohol: withdrawal state with delirium (disorder) | 192211007 | F10 | Main analysis | Alcohol |
| Mental and behavioural disorders due to use of alcohol: psychotic disorder (& [hallucinosis] or [jealousy] or [paranoia] or [psychosis NOS] | 192212000 | F10 | Main analysis | Alcohol |
| Mental and behavioral disorders due to use of alcohol: amnesic syndrome (disorder) | 192213005 | F10 | Main analysis | Alcohol |
| Mental and behavioural disorders due to use of alcohol: residual and late-onset psychotic disorder) or (chronic alcoholic brain syndrome [& dementia NOS] | 192214004 | F10 | Main analysis | Alcohol |
| Mental and behavioral disorders due to use of alcohol: other mental and behavioral disorders (disorder) | 192215003 | F10 | Main analysis | Alcohol |
| Mental and behavioral disorders due to use of alcohol: unspecified mental and behavioral disorder (disorder) | 192216002 | F10 | Main analysis | Alcohol |
| Alcohol blood level excessive (situation) | 207273009 | R78.0 | Main analysis | Alcohol |
| Ethyl alcohol causing toxic effect (disorder) | 212806006 | T51 | Main analysis | Alcohol |
| Grain alcohol causing toxic effect | 212807002 | T51 | Main analysis | Alcohol |
| Ethyl alcohol causing toxic effect NOS (disorder) | 212808007 | T51 | Main analysis | Alcohol |
| Methyl alcohol causing toxic effect | 212809004 | T51 | Main analysis | Alcohol |
| Wood alcohol causing toxic effect (disorder) | 212811008 | T51 | Main analysis | Alcohol |
| Toxic effect of isopropyl alcohol | 212813006 | T51 | Main analysis | Alcohol |
| Dimethyl carbinol causing toxic effect (disorder) | 212814000 | T51 | Main analysis | Alcohol |
| Isopropanol causing toxic effect (disorder) | 212815004 | T51 | Main analysis | Alcohol |
| Rubbing alcohol causing toxic effect (disorder) | 212816003 | T51 | Main analysis | Alcohol |
| Isopropyl alcohol causing toxic effect NOS (disorder) | 212817007 | T51 | Main analysis | Alcohol |
| Fusel oil causing toxic effect NOS (disorder) | 212818002 | T51 | Main analysis | Alcohol |
| Other alcohol causing toxic effect (disorder) | 212819005 | T51 | Main analysis | Alcohol |
| Alcohol causing toxic effect NOS (disorder) | 212820004 | T51 | Main analysis | Alcohol |
| Toxic effect of other alcohols (disorder) | 213687005 | T51 | Main analysis | Alcohol |
| Accidental poisoning by alcoholic beverage | 216633005 | T51 | Main analysis | Alcohol |
| Accidental poisoning by denatured alcohol | 216635003 | T51 | Main analysis | Alcohol |

### SNOMED CT-Codes

| Definition | SNOMED code | ICD-10 Equivalent[2] | Analysis | Substance type |
|---|---|---|---|---|
| Accidental poisoning by methylated spirit | 216636002 | T51 | Main analysis | Alcohol |
| Accidental poisoning by methanol | 216640006 | T51 | Main analysis | Alcohol |
| Accidental poisoning by isopropyl alcohol | 216645001 | T51 | Main analysis | Alcohol |
| Accidental poisoning by rubbing alcohol substitute | 216648004 | T51 | Main analysis | Alcohol |
| Accidental poisoning by fusel oil | 216651006 | T51 | Main analysis | Alcohol |
| Accidental poisoning by and exposure to alcohol, occurrence at home (event) | 221843007 | X45 | Main analysis | Alcohol |
| Accidental poisoning by and exposure to alcohol, occurrence in residential institution (event) | 221844001 | X45 | Main analysis | Alcohol |
| Accidental poisoning by and exposure to alcohol, occurrence at school, other institution and public administrative area (event) | 221845000 | X45 | Main analysis | Alcohol |
| Accidental poisoning by and exposure to alcohol, occurrence at sports and athletics area (event) | 221846004 | X45 | Main analysis | Alcohol |
| Accidental poisoning by and exposure to alcohol, occurrence on street and highway (event) | 221847008 | X45 | Main analysis | Alcohol |
| Accidental poisoning by and exposure to alcohol, occurrence at trade and service area (event) | 221848003 | X45 | Main analysis | Alcohol |
| Accidental poisoning by and exposure to alcohol, occurrence at industrial and construction area (event) | 221849006 | X45 | Main analysis | Alcohol |
| Accidental poisoning by and exposure to alcohol, occurrence on farm (event) | 221850006 | X45 | Main analysis | Alcohol |
| Accidental poisoning by and exposure to alcohol, occurrence at other specified place (event) | 221851005 | X45 | Main analysis | Alcohol |
| Accidental poisoning by and exposure to alcohol, occurrence at unspecified place (event) | 221852003 | X45 | Main analysis | Alcohol |
| Intentional self-poisoning by and exposure to alcohol (event) | 222103001 | X65 | Main analysis | Alcohol |
| Intentional self-poisoning by and exposure to alcohol, occurrence at home (event) | 222104007 | X65 | Main analysis | Alcohol |
| Intentional self-poisoning by and exposure to alcohol, occurrence in residential institution (event) | 222105008 | X65 | Main analysis | Alcohol |
| Intentional self-poisoning by and exposure to alcohol, occurrence at school, other institution and public administrative area (event) | 222106009 | X65 | Main analysis | Alcohol |
| Intentional self-poisoning by and exposure to alcohol, occurrence at sports and athletics area (event) | 222107000 | X65 | Main analysis | Alcohol |
| Intentional self-poisoning by and exposure to alcohol, occurrence on street and highway (event) | 222108005 | X65 | Main analysis | Alcohol |
| Intentional self-poisoning by and exposure to alcohol, occurrence at trade and service area (event) | 222110007 | X65 | Main analysis | Alcohol |
| Intentional self-poisoning by and exposure to alcohol, occurrence at industrial and construction area (event) | 222111006 | X65 | Main analysis | Alcohol |
| Intentional self-poisoning by and exposure to alcohol, occurrence on farm (event) | 222112004 | X65 | Main analysis | Alcohol |
| Intentional self-poisoning by and exposure to alcohol, occurrence at other specified place (event) | 222113009 | X65 | Main analysis | Alcohol |
| Intentional self-poisoning by and exposure to alcohol, occurrence at unspecified place (event) | 222114003 | X65 | Main analysis | Alcohol |
| Poisoning by and exposure to alcohol, undetermined intent (event) | 222702003 | Y15 | Main analysis | Alcohol |
| Poisoning by and exposure to alcohol, occurrence at home, undetermined intent (event) | 222703008 | Y15 | Main analysis | Alcohol |
| Poisoning by and exposure to alcohol, occurrence in residential institution, undetermined intent (event) | 222704002 | Y15 | Main analysis | Alcohol |
| Poisoning by and exposure to alcohol, occurrence at school, other institution and public administrative area, undetermined intent (event) | 222705001 | Y15 | Main analysis | Alcohol |
| Poisoning by and exposure to alcohol, occurrence at sports and athletics area, undetermined intent (event) | 222706000 | Y15 | Main analysis | Alcohol |
| Poisoning by and exposure to alcohol, occurrence on street and highway, undetermined intent (event) | 222707009 | Y15 | Main analysis | Alcohol |

| SNOMED CT-Codes | | | | |
|---|---|---|---|---|
| **Definition** | **SNOMED code** | **ICD-10 Equivalent[2]** | **Analysis** | **Substance type** |
| Poisoning by and exposure to alcohol, occurrence at trade and service area, undetermined intent (event) | 222708004 | Y15 | Main analysis | Alcohol |
| Poisoning by and exposure to alcohol, occurrence at industrial and construction area, undetermined intent (event) | 222709007 | Y15 | Main analysis | Alcohol |
| Poisoning by and exposure to alcohol, occurrence on farm, undetermined intent (event) | 222710002 | Y15 | Main analysis | Alcohol |
| Poisoning by and exposure to alcohol, occurrence at other specified place, undetermined intent (event) | 222711003 | Y15 | Main analysis | Alcohol |
| Poisoning by and exposure to alcohol, occurrence at unspecified place, undetermined intent (event) | 222713000 | Y15 | Main analysis | Alcohol |
| Evidence of alcohol involvement determined by blood alcohol level (navigational concept) | 223333005 | Y90 | Main analysis | Alcohol |
| Evidence of alcohol involvement determined by blood alcohol level of less than 20 mg/100 ml (navigational concept) | 223334004 | Y90 | Main analysis | Alcohol |
| Evidence of alcohol involvement determined by blood alcohol level of 20-39 mg/100 ml (navigational concept) | 223335003 | Y90 | Main analysis | Alcohol |
| Evidence of alcohol involvement determined by blood alcohol level of 40-59 mg/100 ml (navigational concept) | 223336002 | Y90 | Main analysis | Alcohol |
| Evidence of alcohol involvement determined by blood alcohol level of 60-79 mg/100 ml (navigational concept) | 223337006 | Y90 | Main analysis | Alcohol |
| Evidence of alcohol involvement determined by blood alcohol level of 80-99 mg/100 ml (navigational concept) | 223338001 | Y90 | Main analysis | Alcohol |
| Evidence of alcohol involvement determined by blood alcohol level of 100-119 mg/100 ml (navigational concept) | 223339009 | Y90 | Main analysis | Alcohol |
| Evidence of alcohol involvement determined by blood alcohol level of 120-199 mg/100 ml (navigational concept) | 223340006 | Y90 | Main analysis | Alcohol |
| Evidence of alcohol involvement determined by blood alcohol level of 200-239 mg/100 ml (navigational concept) | 223341005 | Y90 | Main analysis | Alcohol |
| Evidence of alcohol involvement determined by blood alcohol level of 240 mg/100 ml or more (navigational concept) | 223342003 | Y90 | Main analysis | Alcohol |
| Evidence of alcohol involvement determined by presence of alcohol in blood, level not specified (navigational concept) | 223343008 | Y90 | Main analysis | Alcohol |
| Evidence of alcohol involvement determined by level of intoxication (navigational concept) | 223344002 | Y91 | Main analysis | Alcohol |
| Evidence of alcohol involvement determined by level of intoxication, mild alcohol intoxication (navigational concept) | 223345001 | Y91 | Main analysis | Alcohol |
| Evidence of alcohol involvement determined by level of intoxication, moderate alcohol intoxication (navigational concept) | 223346000 | Y91 | Main analysis | Alcohol |
| Evidence of alcohol involvement determined by level of intoxication, severe alcohol intoxication (navigational concept) | 223347009 | Y91 | Main analysis | Alcohol |
| Evidence of alcohol involvement determined by level of intoxication, very severe alcohol intoxication (navigational concept) | 223348004 | Y91 | Main analysis | Alcohol |
| Evidence of alcohol involvement determined by level of intoxication, alcohol involvement, not otherwise specified (navigational concept) | 223349007 | Y91 | Main analysis | Alcohol |
| Drinks in morning to get rid of hangover | 228310006 | F10 | Main analysis | Alcohol |
| Binge drinker | 228315001 | F10 | Main analysis | Alcohol |

### SNOMED CT-Codes

| Definition | SNOMED code | ICD-10 Equivalent[2] | Analysis | Substance type |
|---|---|---|---|---|
| Alcoholic binges exceeding sensible amounts | 228316000 | F10 | Main analysis | Alcohol |
| Alcoholic binges exceeding safe amounts | 228317009 | F10 | Main analysis | Alcohol |
| Drinking episode | 228322009 | F10 | Main analysis | Alcohol |
| Drinking bout | 228323004 | F10 | Main analysis | Alcohol |
| Unable to abstain from drinking | 228341007 | F10 | Main analysis | Alcohol |
| Behavioural tolerance to alcohol | 228350009 | F10 | Main analysis | Alcohol |
| Physical tolerance to alcohol | 228351008 | F10 | Main analysis | Alcohol |
| Reverse tolerance to alcohol | 228353006 | F10 | Main analysis | Alcohol |
| Drink driving | 228354000 | F10 | Main analysis | Alcohol |
| Persistent effect of alcohol | 228357007 | F10 | Main analysis | Alcohol |
| Alcoholic coma | 230800004 | Y91 | Main analysis | Alcohol |
| Accidental exposure to alcohol | 242263000 | X45 | Main analysis | Alcohol |
| Accidental exposure to ethanol | 242265007 | X45 | Main analysis | Alcohol |
| Chronic alcoholism (disorder) | 268639004 | F10 | Main analysis | Alcohol |
| Mental and behavioral disorders due to use of alcohol: dependence syndrome (disorder) | 268683008 | F10 | Main analysis | Alcohol |
| Mental and behavioral disorders due to use of alcohol: psychotic disorder (disorder) | 268684002 | F10 | Main analysis | Alcohol |
| Mental and behavioral disorders due to use of alcohol: residual and late-onset psychotic disorder (disorder) | 268685001 | F10 | Main analysis | Alcohol |
| Accidental poisoning by alcohol | 269765000 | T51 | Main analysis | Alcohol |
| Finding of alcohol in blood | 274776000 | R78.0 | Main analysis | Alcohol |
| Alcoholic macrocytosis | 278363000 | X45 | Main analysis | Alcohol |
| Accidental poisoning with ethyl alcohol | 287166006 | T51 | Main analysis | Alcohol |
| Abstinent alcoholic | 300939009 | F10 | Main analysis | Alcohol |
| Ethanol abuse (finding) | 304606004 | F10 | Main analysis | Alcohol |
| Alcohol withdrawal-induced convulsion | 308742005 | F10 | Main analysis | Alcohol |
| Pain in lymph nodes after alcohol consumption | 315226008 | T51 | Main analysis | Alcohol |
| Alcohol-related fit | 361267005 | Y91 | Main analysis | Alcohol |
| Ethanol in blood specimen above reference range | 441685000 | R78.0 | Main analysis | Alcohol |
| Ethanol in blood specimen above legal threshold for operating vehicle | 442669008 | R78.0 | Main analysis | Alcohol |
| Poisoning by benzene | 442764005 | T51 | Main analysis | Alcohol |
| Alcohol in blood specimen above reference range | 442766007 | R78.0 | Main analysis | Alcohol |
| Mild alcohol dependence | 713583005 | F10 | Main analysis | Alcohol |
| Moderate alcohol dependence | 714829008 | F10 | Main analysis | Alcohol |
| Disorder due to alcohol abuse | 288021000119107 | F10 | Main analysis | Alcohol |
| Perceptual disturbance due to alcohol withdrawal | 288041000119101 | F10 | Main analysis | Alcohol |
| Alcohol dependence in childbirth | 10755041000119100 | F10 | Main analysis | Alcohol |

### SNOMED CT-Codes

| Definition | SNOMED code | ICD-10 Equivalent[2] | Analysis | Substance type |
|---|---|---|---|---|
| current drinking | 219006 | | Main analysis | Alcohol |
| Alcoholism counselling | 24165007 | | Main analysis | Alcohol |
| Smell of alcohol on breath | 28045007 | | Main analysis | Alcohol |
| Alcohol rehabilitation | 35637008 | | Main analysis | Alcohol |
| Referral to alcoholism rehabilitation service (procudeure) | 38670004 | | Main analysis | Alcohol |
| Substance with alcohol structure | 53041004 | | Main analysis | Alcohol |
| Alcoholic beverage | 53527002 | | Main analysis | Alcohol |
| Heavy drinker | 86933000 | | Main analysis | Alcohol |
| Alcohol intolerance | 102612005 | | Main analysis | Alcohol |
| Feeling intoxicated | 102897001 | | Main analysis | Alcohol |
| Under care of community alcohol team | 135827004 | | Main analysis | Alcohol |
| Alcohol intake | 160573003 | | Main analysis | Alcohol |
| Suspect alcohol abuse - denied | 160581002 | | Main analysis | Alcohol |
| Alcohol intake within recommended sensible limits | 160593006 | | Main analysis | Alcohol |
| Alcohol consumption NOS | 160599005 | | Main analysis | Alcohol |
| O/E - breath - alcohol smell | 163184002 | | Main analysis | Alcohol |
| Alcohol leaflet given | 183098002 | | Main analysis | Alcohol |
| Admitted to alcohol detoxification centre | 183486001 | | Main analysis | Alcohol |
| Other alcoholic psychosis | 191479003 | | Main analysis | Alcohol |
| Other alcoholic psychosis NOS | 191481001 | | Main analysis | Alcohol |
| Alcoholic psychosis NOS | 191482008 | | Main analysis | Alcohol |
| Acute alcoholic intoxication, unspecified, in alcoholism | 191803009 | | Main analysis | Alcohol |
| Acute alcoholic intoxication in alcoholism NOS | 191807005 | | Main analysis | Alcohol |
| Unspecified chronic alcoholism | 191809008 | | Main analysis | Alcohol |
| Chronic alcoholism NOS | 191814007 | | Main analysis | Alcohol |
| Alcohol dependence syndrome NOS | 191815008 | | Main analysis | Alcohol |
| Nondepenedent alcohol abuse, NOS | 191885000 | | Main analysis | Alcohol |
| Methanol causing toxic effect | 212810009 | | Main analysis | Alcohol |
| Methyl alcohol causing toxic effect NOS | 212812001 | | Main analysis | Alcohol |
| Accidental poisoning by alcohol, NEC | 216632000 | | Main analysis | Alcohol |
| Accidental poisoning by other ethyl alcohol and its products | 216634004 | | Main analysis | Alcohol |
| Accidental poisoning by grain alcohol NOS | 216637006 | | Main analysis | Alcohol |
| Accidental poisoning by ethanol, NOS | 216638001 | | Main analysis | Alcohol |
| Accidental poisoning by ethyl alcohol NOS | 216639009 | | Main analysis | Alcohol |
| Accidental poisoning by wood alcohol | 216643008 | | Main analysis | Alcohol |

### SNOMED CT-Codes

| Definition | SNOMED code | ICD-10 Equivalent[2] | Analysis | Substance type |
|---|---|---|---|---|
| Accidental poisoning by methyl alcohol NOS | 216644002 | | Main analysis | Alcohol |
| Accidental poisoning by dimethyl carbinol | 216646000 | | Main analysis | Alcohol |
| Accidental poisoning by secondary propyl alcohol | 216649007 | | Main analysis | Alcohol |
| Accidental poisoning by isopropyl alcohol NOS | 216650007 | | Main analysis | Alcohol |
| Accidental poisoning by other alcohols | 216652004 | | Main analysis | Alcohol |
| Accidental poisoning by alcohol NOS | 216653009 | | Main analysis | Alcohol |
| [X]Accidental poisoning by and exposure to alcohol | 221842002 | | Main analysis | Alcohol |
| Finding relating to alcohol drinking behaviour | 228273003 | | Main analysis | Alcohol |
| Problem drinker (finding) | 228281002 | | Main analysis | Alcohol |
| Drinks alcohol evenly through week | 228312003 | | Main analysis | Alcohol |
| Drinks alcohol unevenly through week | 228313008 | | Main analysis | Alcohol |
| binge drinking | 228326007 | | Main analysis | Alcohol |
| Feels effect of alcohol at work | 228358002 | | Main analysis | Alcohol |
| Feels afraid of being an alcoholic | 228364009 | | Main analysis | Alcohol |
| Inebriety NOS | 231464007 | | Main analysis | Alcohol |
| Drunkenness NOS | 231465008 | | Main analysis | Alcohol |
| [D]Alcohol blood elevated | 274257003 | | Main analysis | Alcohol |
| Alcohol products adverse reaction | 292880007 | | Main analysis | Alcohol |
| Alcohol products allergy | 294420000 | | Main analysis | Alcohol |
| [V] Alcohol use | 302237007 | | Main analysis | Alcohol |
| [V] Alcohol use | 307730003 | | Main analysis | Alcohol |
| Ingestible alcohol | 311492009 | | Main analysis | Alcohol |
| [V]Alcohol rehabilitation | 316322002 | | Main analysis | Alcohol |
| [V]Alcohol abuse counselling and surveillance | 316494009 | | Main analysis | Alcohol |
| Alcohol intake - finding | 365967005 | | Main analysis | Alcohol |
| Pattern of alcohol consumption through week - finding | 365973006 | | Main analysis | Alcohol |
| Alcohol abuse prevention | 408945004 | | Main analysis | Alcohol |
| Alcohol abuse prevention assessment | 408946003 | | Main analysis | Alcohol |
| Alcohol abuse prevention education | 408947007 | | Main analysis | Alcohol |
| Alcohol abuse prevention management | 408948002 | | Main analysis | Alcohol |
| Alcohol + dextrose | 412198003 | | Main analysis | Alcohol |
| Alcohol consumption counselling | 413473000 | | Main analysis | Alcohol |
| Suspected alcohol abuse | 415685003 | | Main analysis | Alcohol |
| Referral to community drug and alcohol team | 417096006 | | Main analysis | Alcohol |
| Alcohol induced hallucinations | 417633001 | | Main analysis | Alcohol |

| SNOMED CT-Codes | | | | |
|---|---|---|---|---|
| **Definition** | **SNOMED code** | **ICD-10 Equivalent[2]** | **Analysis** | **Substance type** |
| Ethanol | 419442005 | | Main analysis | Alcohol |
| Alcohol agent | 419572002 | | Main analysis | Alcohol |
| Allergy to ethanol | 420140004 | | Main analysis | Alcohol |
| Alcohol consumption during pregnancy | 427013000 | | Main analysis | Alcohol |
| Alcohol withdrawal scale | 429501006 | | Main analysis | Alcohol |
| Alcohol intake exceeds recommended daily limit | 429775004 | | Main analysis | Alcohol |
| Referral to specialist alcohol treatment service | 431260004 | | Main analysis | Alcohol |
| Assessment using alcohol withdrawal scale | 445628007 | | Main analysis | Alcohol |
| Alcohol abuse | 15167005 | | Main analysis | Alcohol |
| **Cannabis/Cannabinoids** | | | | |
| Cannabis use disorder | 37344009 | | Main analysis | Cannabis |
| Cannabis intoxication delirium | 39807006 | | Main analysis | Cannabis |
| Cannabis dependence, episodic | 191838006 | | Main analysis | Cannabis |
| Nondependent cannabis abuse, continuous | 191893000 | | Main analysis | Cannabis |
| Nondependent cannabis abuse, episodic | 191894006 | | Main analysis | Cannabis |
| Cannabis misuse | 428823006 | | Main analysis | Cannabis |
| Synthetic cannabinoid abuse | 737336003 | | Main analysis | Cannabis |
| Mental and behavioural disorders due to use of cannabinoids | 26714005 | F12 | Main analysis | Cannabis |
| Mental and behavioural disorders due to use of cannabinoids | 39951001 | F12 | Main analysis | Cannabis |
| Mental and behavioural disorders due to use of cannabinoids | 63649001 | F12 | Main analysis | Cannabis |
| Mental and behavioural disorders due to use of cannabinoids | 77355000 | F12 | Main analysis | Cannabis |
| Mental and behavioural disorders due to use of cannabinoids | 85005007 | F12 | Main analysis | Cannabis |
| Mental and behavioural disorders due to use of cannabinoids | 268641003 | F12 | Main analysis | Cannabis |
| Mental and behavioural disorders due to use of cannabinoids | 703848005 | F12 | Main analysis | Cannabis |
| **Drug unspecified or multiple** | | | | |
| Psychoactive substance dependence | 2403008 | | Main analysis | Drug unspecified or multiple |
| Dependent drug abuse | 6525002 | | Main analysis | Drug unspecified or multiple |
| Drug habituation | 9769006 | | Main analysis | Drug unspecified or multiple |
| Combined alcohol and drug rehabilitation and detoxification | 23915005 | | Main analysis | Drug unspecified or multiple |
| Drug abuse | 26416006 | | Main analysis | Drug unspecified or multiple |
| Non dependent drug abuse | 49540005 | | Main analysis | Drug unspecified or multiple |
| Drug rehabilitation and detoxification | 56876005 | | Main analysis | Drug unspecified or multiple |
| Drug detoxification | 61480009 | | Main analysis | Drug unspecified or multiple |
| Detoxification psychiatric therapy for alcoholism | 64297001 | | Main analysis | Drug unspecified or multiple |
| Substance use disorder | 66214007 | | Main analysis | Drug unspecified or multiple |

### SNOMED CT-Codes

| Definition | SNOMED code | ICD-10 Equivalent[2] | Analysis | Substance type |
|---|---|---|---|---|
| Narcotic drug user | 70545002 | | Main analysis | Drug unspecified or multiple |
| Combined alcohol and drug detoxification | 87106005 | | Main analysis | Drug unspecified or multiple |
| Psychoactive substance abuse | 91388009 | | Main analysis | Drug unspecified or multiple |
| Maternal drug use | 95607001 | | Main analysis | Drug unspecified or multiple |
| Occasional drug abuser | 105546006 | | Main analysis | Drug unspecified or multiple |
| Chronic drug abuse | 110281001 | | Main analysis | Drug unspecified or multiple |
| Dependent drug detoxification | 182969009 | | Main analysis | Drug unspecified or multiple |
| Nondependent mixed drug abuse | 191934007 | | Main analysis | Drug unspecified or multiple |
| Nondependent mixed drug abuse, continuous | 191936009 | | Main analysis | Drug unspecified or multiple |
| Nondependent mixed drug abuse, episodic | 191937000 | | Main analysis | Drug unspecified or multiple |
| Injecting drug user | 226034001 | | Main analysis | Drug unspecified or multiple |
| Long-term drug misuser | 228371004 | | Main analysis | Drug unspecified or multiple |
| Poly-drug misuser | 228372006 | | Main analysis | Drug unspecified or multiple |
| Drug addict | 228373001 | | Main analysis | Drug unspecified or multiple |
| Notified addict | 228374007 | | Main analysis | Drug unspecified or multiple |
| Misuses drugs orally | 228375008 | | Main analysis | Drug unspecified or multiple |
| Inhales drugs | 228376009 | | Main analysis | Drug unspecified or multiple |
| Smokes drugs | 228377000 | | Main analysis | Drug unspecified or multiple |
| Smokes drugs in cigarette form | 228378005 | | Main analysis | Drug unspecified or multiple |
| Smokes drugs through a pipe | 228379002 | | Main analysis | Drug unspecified or multiple |
| Sniffs drugs | 228381000 | | Main analysis | Drug unspecified or multiple |
| Misuses drugs vaginally | 228382007 | | Main analysis | Drug unspecified or multiple |
| Misuses drugs rectally | 228383002 | | Main analysis | Drug unspecified or multiple |
| Misuses drugs sublingually | 228384008 | | Main analysis | Drug unspecified or multiple |
| Injects drugs subcutaneously | 228386005 | | Main analysis | Drug unspecified or multiple |
| Injects drugs intramuscularly | 228387001 | | Main analysis | Drug unspecified or multiple |
| Intravenous drug user | 228388006 | | Main analysis | Drug unspecified or multiple |
| Groin injector | 228389003 | | Main analysis | Drug unspecified or multiple |
| Frequency of drug misuse | 228390007 | | Main analysis | Drug unspecified or multiple |
| Drug injection behaviour | 228391006 | | Main analysis | Drug unspecified or multiple |
| Drug injecting equipment hygiene | 228394003 | | Main analysis | Drug unspecified or multiple |
| Cleaning of drug injection equipment | 228401002 | | Main analysis | Drug unspecified or multiple |
| Priority of drug-related activities | 228415000 | | Main analysis | Drug unspecified or multiple |
| Time devoted to drug-related activities | 228421001 | | Main analysis | Drug unspecified or multiple |
| Time spent obtaining drugs | 228422008 | | Main analysis | Drug unspecified or multiple |

### SNOMED CT-Codes

| Definition | SNOMED code | ICD-10 Equivalent[2] | Analysis | Substance type |
|---|---|---|---|---|
| Time spent taking drugs | 228423003 | | Main analysis | Drug unspecified or multiple |
| Routine of drug-related activities | 228425005 | | Main analysis | Drug unspecified or multiple |
| Drug-related rituals | 228429004 | | Main analysis | Drug unspecified or multiple |
| Drug addiction therapy | 266707007 | | Main analysis | Drug unspecified or multiple |
| Illicit drug use | 307052004 | | Main analysis | Drug unspecified or multiple |
| Habitual drug user | 361049005 | | Main analysis | Drug unspecified or multiple |
| Substance abuser | 361055000 | | Main analysis | Drug unspecified or multiple |
| Substance use treatment: drug withdrawal | 386450006 | | Main analysis | Drug unspecified or multiple |
| Substance use treatment: overdose | 386451005 | | Main analysis | Drug unspecified or multiple |
| Drug dependence home detoxification | 414054004 | | Main analysis | Drug unspecified or multiple |
| Drug dependence self detoxification | 414056002 | | Main analysis | Drug unspecified or multiple |
| Continuous use of drugs | 416262000 | | Main analysis | Drug unspecified or multiple |
| Preoccupied with substance misuse | 416437003 | | Main analysis | Drug unspecified or multiple |
| Episodic use of drugs | 417252002 | | Main analysis | Drug unspecified or multiple |
| Current drug user | 417284009 | | Main analysis | Drug unspecified or multiple |
| Recreational drug user | 424848002 | | Main analysis | Drug unspecified or multiple |
| Episodic drug abuse | 425533007 | | Main analysis | Drug unspecified or multiple |
| Drug use behaviour during pregnancy | 440664005 | | Main analysis | Drug unspecified or multiple |
| Amount of money spent per day on drug habit | 442229000 | | Main analysis | Drug unspecified or multiple |
| Polysubstance abuse | 445273005 | | Main analysis | Drug unspecified or multiple |
| Illicit drug overdose | 708079007 | | Main analysis | Drug unspecified or multiple |
| Novel psychoactive substance misuse | 713775002 | | Main analysis | Drug unspecified or multiple |
| Illicit drug injection in last 12 months | 741063003 | | Main analysis | Drug unspecified or multiple |
| Intoxication caused by recreational drug misuse | 772808000 | | Main analysis | Drug unspecified or multiple |
| Intoxication | 1149322001 | | Main analysis | Drug unspecified or multiple |
| Psychostimulant dependence episodic | 1231333004 | | Main analysis | Drug unspecified or multiple |
| Substance dependence | 1254812006 | | Main analysis | Drug unspecified or multiple |
| Substance dependence, episodic | 1254974001 | | Main analysis | Drug unspecified or multiple |
| Substance dependence in childbirth | 1254987007 | | Main analysis | Drug unspecified or multiple |
| Combined substance dependence, excluding opioid, continuous | 1255017007 | | Main analysis | Drug unspecified or multiple |
| Combined substance dependence, excluding opioid, in remission | 1255020004 | | Main analysis | Drug unspecified or multiple |
| Postpartum substance dependence | 1259017004 | | Main analysis | Drug unspecified or multiple |
| Patient self-medicating with illicit substance | 1334001000168100 | | Main analysis | Drug unspecified or multiple |
| Drug addiction therapy using non-opioid medicine | 1545151000168100 | | Main analysis | Drug unspecified or multiple |
| Management of simple withdrawal from substance use | 1648321000168100 | | Main analysis | Drug unspecified or multiple |

### SNOMED CT-Codes

| Definition | SNOMED code | ICD-10 Equivalent[2] | Analysis | Substance type |
|---|---|---|---|---|
| Management of complex withdrawal from substance use | 1648331000168100 | | Main analysis | Drug unspecified or multiple |
| Episodic polysubstance dependence | 16076691000119100 | | Main analysis | Drug unspecified or multiple |
| Mental and behavioural disorders due to multiple drug use and use of other psychoactive substances | 11061003 | F22 | Main analysis | Drug unspecified or multiple |
| Poisoning by other and unspecified narcotics | 11196001 | T40.6 | Main analysis | Drug unspecified or multiple |
| Mental and behavioural disorders due to multiple drug use and use of other psychoactive substances | 11387009 | F23 | Main analysis | Drug unspecified or multiple |
| Mental and behavioural disorders due to multiple drug use and use of other psychoactive substances | 28368009 | F25 | Main analysis | Drug unspecified or multiple |
| Mental and behavioural disorders due to multiple drug use and use of other psychoactive substances | 32709003 | F26 | Main analysis | Drug unspecified or multiple |
| Mental and behavioural disorders due to multiple drug use and use of other psychoactive substances | 43242008 | F27 | Main analysis | Drug unspecified or multiple |
| Mental and behavioural disorders due to multiple drug use and use of other psychoactive substances | 50026000 | F28 | Main analysis | Drug unspecified or multiple |
| Mental and behavioural disorders due to multiple drug use and use of other psychoactive substances | 51339003 | F29 | Main analysis | Drug unspecified or multiple |
| Mental and behavioural disorders due to multiple drug use and use of other psychoactive substances | 74934004 | F32 | Main analysis | Drug unspecified or multiple |
| Mental and behavioural disorders due to multiple drug use and use of other psychoactive substances | 84584008 | F33 | Main analysis | Drug unspecified or multiple |
| Mental and behavioural disorders due to multiple drug use and use of other psychoactive substances | 191483003 | F19 | Main analysis | Drug unspecified or multiple |
| Mental and behavioural disorders due to multiple drug use and use of other psychoactive substances | 191484009 | F19 | Main analysis | Drug unspecified or multiple |
| Mental and behavioural disorders due to multiple drug use and use of other psychoactive substances | 191485005 | F19 | Main analysis | Drug unspecified or multiple |
| Mental and behavioural disorders due to multiple drug use and use of other psychoactive substances | 191486006 | F19 | Main analysis | Drug unspecified or multiple |
| Mental and behavioural disorders due to multiple drug use and use of other psychoactive substances | 191492000 | F19 | Main analysis | Drug unspecified or multiple |
| Mental and behavioural disorders due to multiple drug use and use of other psychoactive substances | 191494004 | F19 | Main analysis | Drug unspecified or multiple |
| Mental and behavioural disorders due to multiple drug use and use of other psychoactive substances | 191495003 | F19 | Main analysis | Drug unspecified or multiple |
| Mental and behavioural disorders due to multiple drug use and use of other psychoactive substances | 191496002 | F19 | Main analysis | Drug unspecified or multiple |
| Mental and behavioural disorders due to multiple drug use and use of other psychoactive substances | 191816009 | F19 | Main analysis | Drug unspecified or multiple |
| Mental and behavioural disorders due to multiple drug use and use of other psychoactive substances | 191865004 | F19 | Main analysis | Drug unspecified or multiple |
| Mental and behavioural disorders due to multiple drug use and use of other psychoactive substances | 191873008 | F19 | Main analysis | Drug unspecified or multiple |
| Mental and behavioural disorders due to multiple drug use and use of other psychoactive substances | 191939002 | F19 | Main analysis | Drug unspecified or multiple |
| Mental and behavioural disorders due to multiple drug use and use of other psychoactive substances | 228438002 | F19 | Main analysis | Drug unspecified or multiple |
| Mental and behavioural disorders due to multiple drug use and use of other psychoactive substances | 231451006 | F19 | Main analysis | Drug unspecified or multiple |
| Mental and behavioural disorders due to multiple drug use and use of other psychoactive substances | 231466009 | F19 | Main analysis | Drug unspecified or multiple |
| Mental and behavioural disorders due to multiple drug use and use of other psychoactive substances | 231481003 | F19 | Main analysis | Drug unspecified or multiple |
| Mental and behavioural disorders due to multiple drug use and use of other psychoactive substances | 231482005 | F19 | Main analysis | Drug unspecified or multiple |
| Mental and behavioural disorders due to multiple drug use and use of other psychoactive substances | 247702001 | F19 | Main analysis | Drug unspecified or multiple |
| Poisoning by other and unspecified narcotics | 290220008 | T40.6 | Main analysis | Drug unspecified or multiple |
| Poisoning by other and unspecified narcotics | 290221007 | T40.6 | Main analysis | Drug unspecified or multiple |
| Poisoning by other and unspecified narcotics | 290222000 | T40.6 | Main analysis | Drug unspecified or multiple |
| Poisoning by other synthetic narcotics | 295154004 | T40.4 | Main analysis | Drug unspecified or multiple |
| Poisoning by other synthetic narcotics | 295167001 | T40.4 | Main analysis | Drug unspecified or multiple |

| SNOMED CT-Codes | | | | |
|---|---|---|---|---|
| **Definition** | **SNOMED code** | **ICD-10 Equivalent[2]** | **Analysis** | **Substance type** |
| Poisoning by other synthetic narcotics | 295169003 | T40.4 | Main analysis | Drug unspecified or multiple |
| Poisoning by other synthetic narcotics | 295190006 | T40.4 | Main analysis | Drug unspecified or multiple |
| Poisoning by other synthetic narcotics | 295193008 | T40.4 | Main analysis | Drug unspecified or multiple |
| Poisoning by other synthetic narcotics | 295194002 | T40.4 | Main analysis | Drug unspecified or multiple |
| Poisoning by other and unspecified narcotics | 295213004 | T40.6 | Main analysis | Drug unspecified or multiple |
| Poisoning by other and unspecified narcotics | 297199006 | T40.6 | Main analysis | Drug unspecified or multiple |
| Mental and behavioural disorders due to multiple drug use and use of other psychoactive substances | 363101005 | F19 | Main analysis | Drug unspecified or multiple |
| Mental and behavioural disorders due to multiple drug use and use of other psychoactive substances | 363314000 | F19 | Main analysis | Drug unspecified or multiple |
| Mental and behavioural disorders due to multiple drug use and use of other psychoactive substances | 365984004 | F19 | Main analysis | Drug unspecified or multiple |
| Mental and behavioural disorders due to multiple drug use and use of other psychoactive substances | 396344000 | F19 | Main analysis | Drug unspecified or multiple |
| Mental and behavioural disorders due to multiple drug use and use of other psychoactive substances | 416119007 | F19 | Main analysis | Drug unspecified or multiple |
| Mental and behavioural disorders due to multiple drug use and use of other psychoactive substances | 429299000 | F19 | Main analysis | Drug unspecified or multiple |
| Mental and behavioural disorders due to multiple drug use and use of other psychoactive substances | 429672007 | F19 | Main analysis | Drug unspecified or multiple |
| Mental and behavioural disorders due to multiple drug use and use of other psychoactive substances | 442351006 | F19 | Main analysis | Drug unspecified or multiple |
| Combined alcohol and drug rehabilitation | 62213004 | | Main analysis | Drug unspecified or multiple |
| **Opioids** | | | | |
| Opioid use disorder | 5602001 | F11 | Main analysis | Opioids |
| Opioid intoxication delirium | 52866005 | | Main analysis | Opioids |
| Opioid dependence | 75544000 | | Main analysis | Opioids |
| Episodic opioid dependence | 191820008 | | Main analysis | Opioids |
| Nondependent opioid abuse | 191909007 | | Main analysis | Opioids |
| Nondependent opioid abuse, continuous | 191912005 | | Main analysis | Opioids |
| Nondependent opioid abuse, episodic | 191913000 | | Main analysis | Opioids |
| Heroin dependence | 231477003 | | Main analysis | Opioids |
| Methadone dependence | 231478008 | | Main analysis | Opioids |
| Morphine dependence | 231479000 | | Main analysis | Opioids |
| Opium dependence | 231480002 | | Main analysis | Opioids |
| Drug addiction therapy using methadone | 310653000 | | Main analysis | Opioids |
| Opiate misuse | 428819003 | | Main analysis | Opioids |
| Methadone misuse | 429512006 | | Main analysis | Opioids |
| Drug addiction therapy using buprenorphine | 792901003 | | Main analysis | Opioids |
| Drug addiction therapy using buprenorphine and naloxone | 792902005 | | Main analysis | Opioids |
| Combined opioid with non-opioid substance dependence, episodic | 1255015004 | | Main analysis | Opioids |
| Opioid dependence, on agonist therapy | 1081000119105 | | Main analysis | Opioids |
| Intravenous nondependent opioid abuse | 145121000119106 | | Main analysis | Opioids |

| SNOMED CT-Codes | | | | |
|---|---|---|---|---|
| **Definition** | **SNOMED code** | **ICD-10 Equivalent[2]** | **Analysis** | **Substance type** |
| Opioid agonist treatment | 1515351000168100 | | Main analysis | Opioids |
| Maintenance of opioid agonist treatment | 1640611000168100 | | Main analysis | Opioids |
| Initiation of opioid agonist treatment | 1640661000168100 | | Main analysis | Opioids |
| Poisoning by heroin | 13187008 | T40.1 | Main analysis | Opioids |
| Mental and behavioural disorders due to use of opioids | 14784000 | F11 | Main analysis | Opioids |
| Poisoning by methadone | 60199004 | T40.3 | Main analysis | Opioids |
| Mental and behavioural disorders due to use of opioids | 77721001 | F11 | Main analysis | Opioids |
| Mental and behavioural disorders due to use of opioids | 87132004 | F11 | Main analysis | Opioids |
| Mental and behavioural disorders due to use of opioids | 191819002 | F11 | Main analysis | Opioids |
| Poisoning by heroin | 216463005 | T40.1 | Main analysis | Opioids |
| Poisoning by methadone | 216464004 | T40.4 | Main analysis | Opioids |
| Poisoning by other opioids | 242828004 | T40.2 | Main analysis | Opioids |
| Poisoning by heroin | 242829007 | T40.1 | Main analysis | Opioids |
| Poisoning by methadone | 242831003 | T40.5 | Main analysis | Opioids |
| Poisoning by heroin | 290182008 | T40.1 | Main analysis | Opioids |
| Poisoning by heroin | 290183003 | T40.1 | Main analysis | Opioids |
| Poisoning by methadone | 295161000 | T40.6 | Main analysis | Opioids |
| Poisoning by methadone | 295163002 | T40.6 | Main analysis | Opioids |
| Poisoning by methadone | 295164008 | T40.6 | Main analysis | Opioids |
| Poisoning by other opioids | 295165009 | T40.3 | Main analysis | Opioids |
| Poisoning by other opioids | 295170002 | T40.2 | Main analysis | Opioids |
| Poisoning by other opioids | 295171003 | T40.2 | Main analysis | Opioids |
| Poisoning by other opioids | 295172005 | T40.2 | Main analysis | Opioids |
| Poisoning by other opioids | 295173000 | T40.2 | Main analysis | Opioids |
| Poisoning by heroin | 295174006 | T40.1 | Main analysis | Opioids |
| Poisoning by heroin | 295175007 | T40.1 | Main analysis | Opioids |
| Poisoning by heroin | 295176008 | T40.1 | Main analysis | Opioids |
| Poisoning by other opioids | 295184007 | T40.2 | Main analysis | Opioids |
| Poisoning by other opioids | 295186009 | T40.2 | Main analysis | Opioids |
| Mental and behavioural disorders due to use of opioids | 426001001 | F11 | Main analysis | Opioids |
| Overdose of opiate | 242253008 | | Main analysis | Opioids |
| **Sedatives and others** | | | | |
| Sedative, hypnotic AND/OR anxiolytic intoxication delirium | 5444000 | | Sensitivity analysis | Sedatives and others |
| PCP abuse | 7071007 | | Main analysis | Sedatives and others |
| Hallucinogen dependence | 38247002 | | Main analysis | Sedatives and others |

### SNOMED CT-Codes

| Definition | SNOMED code | ICD-10 Equivalent[2] | Analysis | Substance type |
|---|---|---|---|---|
| Hallucinogen intoxication | 50320000 | | Main analysis | Sedatives and others |
| Inhalant intoxication | 60901005 | | Main analysis | Sedatives and others |
| Sedative abuse | 64386003 | | Main analysis | Sedatives and others |
| Inhalant abuse | 70340006 | | Main analysis | Sedatives and others |
| Hallucinogen abuse | 74851005 | | Main analysis | Sedatives and others |
| Abuses volatile solvents | 105549004 | | Main analysis | Sedatives and others |
| Hallucinogen dependence, continuous | 191849000 | | Main analysis | Sedatives and others |
| Glue sniffing dependence, episodic | 191856006 | | Main analysis | Sedatives and others |
| Nondependent hallucinogen abuse, continuous | 191899001 | | Main analysis | Sedatives and others |
| Nondependent hallucinogen abuse, episodic | 191900006 | | Main analysis | Sedatives and others |
| Barbiturate abuse | 231462006 | | Main analysis | Sedatives and others |
| Lysergic acid diethylamide dependence | 231468005 | | Main analysis | Sedatives and others |
| Nondependent hallucinogen abuse | 268646008 | | Main analysis | Sedatives and others |
| Psychostimulant dependence | 275471001 | | Main analysis | Sedatives and others |
| Continuous inhalant abuse | 426095000 | | Main analysis | Sedatives and others |
| Drug abuse, continuous | 426590003 | | Main analysis | Sedatives and others |
| Episodic inhalant abuse | 427229002 | | Main analysis | Sedatives and others |
| Solvent misuse | 428495004 | | Main analysis | Sedatives and others |
| Barbiturate misuse | 428623008 | | Main analysis | Sedatives and others |
| Ketamine abuse | 724713006 | | Main analysis | Sedatives and others |
| Dependence due to ketamine | 724715004 | | Main analysis | Sedatives and others |
| Harmful use of hypnotic | 772999000 | | Main analysis | Sedatives and others |
| Nondependent hypnotic abuse, episodic | 1230074002 | | Main analysis | Sedatives and others |
| Nondependent hypnotic abuse, continuous | 1230077009 | | Main analysis | Sedatives and others |
| Nondependent hypnotic abuse | 1230081009 | | Main analysis | Sedatives and others |
| Hypnotic dependence, episodic | 1231160001 | | Main analysis | Sedatives and others |
| Inhalant dependence, continuous | 1382581000168100 | | Main analysis | Sedatives and others |
| Continuous sedative abuse | 145841000119107 | | Main analysis | Sedatives and others |
| Episodic phencyclidine abuse | 1382351000168100 | | Main analysis | Sedatives and others |
| Aerosol inhalation dependence | 1382391000168100 | | Main analysis | Sedatives and others |
| Petrol sniffing dependence | 1382421000168100 | | Main analysis | Sedatives and others |
| Paint sniffing dependence | 1382521000168100 | | Main analysis | Sedatives and others |
| Aerosol inhalation abuse | 1382531000168100 | | Main analysis | Sedatives and others |
| Petrol sniffing abuse | 1382541000168100 | | Main analysis | Sedatives and others |
| Paint sniffing abuse | 1382551000168100 | | Main analysis | Sedatives and others |

| SNOMED CT-Codes | | | | |
|---|---|---|---|---|
| **Definition** | **SNOMED code** | **ICD-10 Equivalent[2]** | **Analysis** | **Substance type** |
| Butane inhalation abuse | 1382561000168100 | | Main analysis | Sedatives and others |
| Nitrite inhalation abuse | 1382571000168100 | | Main analysis | Sedatives and others |
| Nitrous oxide inhalation abuse | 1382581000168100 | | Main analysis | Sedatives and others |
| Glue sniffing abuse | 145841000119107 | | Main analysis | Sedatives and others |
| Mental and behavioural disorders due to multiple drug use and use of other psychoactive substances | 88320008 | F18 | Main analysis | Sedatives and others |
| Mental and behavioural disorders due to use of volatile solvents | 191853003 | F18 | Main analysis | Sedatives and others |
| Mental and behavioural disorders due to use of sedatives | 231473004 | F13 | Main analysis | Sedatives and others |
| Continuous phencyclidine abuse | 425885002 | | Main analysis | Sedatives and others |
| **Stimulants** | | | | |
| Amphetamine dependence | 21647008 | | Main analysis | Stimulants |
| Cocaine intoxication | 27956007 | | Main analysis | Stimulants |
| Cocaine dependence | 31956009 | | Main analysis | Stimulants |
| Cocaine use disorder | 78267003 | | Main analysis | Stimulants |
| Cocaine dependence, continuous | 191831000 | | Main analysis | Stimulants |
| Nondependent cocaine abuse | 191916008 | | Main analysis | Stimulants |
| Nondependent cocaine abuse, continuous | 191918009 | | Main analysis | Stimulants |
| Nondependent cocaine abuse, episodic | 191919001 | | Main analysis | Stimulants |
| Overdose of cocaine | 296321004 | | Main analysis | Stimulants |
| Crack cocaine misuse | 428493006 | | Main analysis | Stimulants |
| Amphetamine misuse | 428659002 | | Main analysis | Stimulants |
| Cocaine misuse | 429782000 | | Main analysis | Stimulants |
| Stimulant abuse | 441527004 | | Main analysis | Stimulants |
| Stimulant dependence | 442406005 | | Main analysis | Stimulants |
| Methamphetamine abuse | 699449003 | | Main analysis | Stimulants |
| Abuse of synthetic cathinone | 762504005 | | Main analysis | Stimulants |
| Synthetic cathinone dependence | 762505006 | | Main analysis | Stimulants |
| Intoxication due to synthetic cathinone | 762671008 | | Main analysis | Stimulants |
| Amphetamine and/or amphetamine derivative use disorder | 785277001 | | Main analysis | Stimulants |
| Intoxication caused by central stimulant | 1155772001 | | Main analysis | Stimulants |
| Amfetamine and/or amfetamine derivative abuse, episodic | 1230071005 | | Main analysis | Stimulants |
| Amfetamine and/or amfetamine derivative abuse, continuous | 1230072003 | | Main analysis | Stimulants |
| Nondependent psychostimulant abuse, continuous | 1230084001 | | Main analysis | Stimulants |
| Nondependent amfetamine and/or amfetamine derivative abuse, continuous | 1230085000 | | Main analysis | Stimulants |
| Nondependent psychostimulant abuse, episodic | 1230086004 | | Main analysis | Stimulants |
| Nondependent amfetamine and/or amfetamine derivative abuse, episodic | 1230087008 | | Main analysis | Stimulants |

### SNOMED CT-Codes

| Definition | SNOMED code | ICD-10 Equivalent[2] | Analysis | Substance type |
|---|---|---|---|---|
| Nondependent amfetamine and/or amfetamine derivative abuse | 1230088003 | | Main analysis | Stimulants |
| Nondependent psychostimulant abuse | 1230089006 | | Main analysis | Stimulants |
| Methylenedioxymethamphetamine dependence | 1231319004 | | Main analysis | Stimulants |
| Intravenous cocaine abuse | 145101000119102 | | Main analysis | Stimulants |
| Catha edulis abuse | 429001000124103 | | Main analysis | Stimulants |
| Methamphetamine intoxication | 12398571000119100 | | Main analysis | Stimulants |
| Mental and behavioural disorders due to use of other stimulants, including caffeine | 8837000 | F15 | Main analysis | Stimulants |
| Mental and behavioural disorders due to use of other stimulants, including caffeine | 45421006 | F15 | Main analysis | Stimulants |
| Mental and behavioural disorders due to use of cocaine | 46975003 | F14 | Main analysis | Stimulants |
| Mental and behavioural disorders due to use of cocaine | 51493001 | F14 | Main analysis | Stimulants |
| Mental and behavioural disorders due to use of cocaine | 80868005 | F14 | Main analysis | Stimulants |
| Mental and behavioural disorders due to use of other stimulants, including caffeine | 84758004 | F15 | Main analysis | Stimulants |
| Poisoning by cocaine | 290544006 | T40.5 | Main analysis | Stimulants |
| Poisoning by cocaine | 290545007 | T40.5 | Main analysis | Stimulants |
| Poisoning by cocaine | 296322006 | T40.5 | Main analysis | Stimulants |
| Poisoning by cocaine | 296323001 | T40.5 | Main analysis | Stimulants |
| Mental and behavioural disorders due to use of other stimulants, including caffeine | 428370001 | F15 | Main analysis | Stimulants |
| Amfetamine and/or amfetamine derivative dependence continuous | 1231328003 | | Main analysis | Stimulants |

Footnotes: 1. SNOMED Clinical Terms® (SNOMED CT®) which is used by permission of the SNOMED International. All rights reserved. SNOMED CT® was originally created by the College of American Pathologists. "SNOMED", "SNOMED CT" and "SNOMED Clinical Terms" are registered trademarks of the SNOMED International (www.snomed.org); 2. ICD10AM equivalent code based on mapping by Yuen 2022
